# Preventive interventions for post Covid-19 condition: systematic review update

**DOI:** 10.1101/2024.09.18.24313918

**Authors:** Jennifer Pillay, Sholeh Rahman, Nicole Gehring, Samantha Guitard, Ashiqur Rahman Ashiq, Lisa Hartling

## Abstract

**Background:** Post COVID-19 condition (PCC) can affect individuals regardless of the severity of their initial illness, and its impact on daily life can be significant. There are uncertainties about whether treatments in the acute or post-acute phase of infection can prevent PCC. We report an update to a previous systematic review on the effects of interventions to prevent PCC.

**Methods:** We updated our previous peer-reviewed searches on February 9, 2024. We searched bibliographic databases and grey literature resources to identify trials and comparative observational studies reporting on any intervention provided during the acute (symptom onset to 4 weeks) or post-acute phase (4-8 weeks) of COVID-19 and our primary outcome of incidence of PCC, ascertained at 3 months or longer following infection and capturing, at a minimum, symptoms of fatigue, dyspnea and one or more aspects of cognitive function. Non-recovery from COVID-19 was included if necessary. Secondary outcomes included fatigue, breathlessness/dyspnea, post-exertional malaise, health-related quality of life, psychopathology, cognitive impairment, hospitalization, return to work/education, and adverse effects of the intervention. For screening we employed artificial intelligence to prioritize records and modified our methods to rely on single-reviewer screening after 50% of citations were screened in duplicate. Study selection and risk of bias assessments were conducted independently by two reviewers and data extraction relied on verification of another reviewer’s work. We grouped studies by intervention type and timing, and by acute-care setting, and performed meta-analysis where appropriate. Sensitivity analyses were conducted for the primary outcome, excluding studies with high risk of bias, using non-recovery as a proxy outcome, and evaluating the outcome at more than 12 months of follow-up. We assessed the certainty of evidence using GRADE.

**Results:** Twenty-four studies (5 randomized and 19 non-randomized), all among adults, were included. The acute care setting in nine studies was outpatient and in 15 studies was in-patient; all but one intervention was administered during the acute-phase of illness. The use of convalescent plasma in outpatient acute COVID-19 care probably does not reduce the risk of PCC (relative risk [RR]: 0.93, 95% CI: 0.77-1.12; 1 RCT; moderate certainty). There was low-certainty evidence suggesting that probiotics (RR [95% CI]: 0.32 [0.13-0.78]; 1 RCT) and metformin (0.50 [0.25-0.99]; 1 RCT among individuals with a BMI ≥25 kg/m^2^) reduce PCC to a small-to-moderate extent in outpatients, while ivermectin (outpatients), antivirals (outpatients), steroids (in-patients), and therapeutic-dose heparin (vs. prophylactic dose; in-patients) may not be effective. Evidence was very low certainty for several other acute-phase pharmacologic intervention and post-acute outpatient assessment and referrals. For outpatient antiviral treatment, while overall PCC risk may not decrease, there might be a slight reduction in psychopathology. Similarly, inpatient antiviral use may not prevent PCC but may offer a small reduction in prolonged general malaise after light exertion. Therapeutic-dose heparin may slightly reduce the risk of cognitive impairment compared to prophylactic-dose heparin among in-patients. The findings remained consistent across all these sensitivity analyses.

**Conclusions:** Evidence suggests that PCC can be prevented to some extent among outpatients with the use of probiotics and metformin during the acute phase of COVID-19. Effects from interventions used among in-patients and within the post-acute phase are uncertain at this time. Evidence on commonly recommended interventions including rehabilitation or multidisciplinary care was lacking.

**Protocol registration:** CRD42024513247

## BACKGROUND

The global pandemic caused by coronavirus disease 2019 (COVID-19) affected over 775 million people and resulted in more than 5 million deaths worldwide [1]. Though most often people infected with SARS-CoV-2 recover within a few weeks [2], some may experience persistent symptoms lasting for several weeks or even months after the initial infection [3,4]. Some people inflicted with symptoms after infection develop post-COVID-condition, with most commonly reported symptoms being fatigue, weakness, and breathlessness among others including “brain fog”, anxiety and depression [5–9]. The term post COVID-19 condition (PCC) was established by the WHO on October 6, 2021, to refer to new or ongoing symptoms in individuals with a history of probable or confirmed SARS-CoV-2 infection, occurring usually 3 months from the onset of the infection and lasting for at least 2 months following initial recovery that cannot be explained by an alternative diagnosis [10]. Reported rates of prevalence of PCC are varied, partly due to differences in source populations (e.g., differing severities of acute COVID-19, health-care seeking) and how definitions are operationalized in studies [11]. Estimates from general populations in Canada and the US suggest that 19% [12] and 18-23% [13,14], respectively, of adults who experience COVID-19 will develop PCC. Interventions to prevent PCC could offer great impact. We previously undertook a systematic review on a wide range of potential preventive interventions and the evidence was at that time (to July 2021) was very uncertain [15].

### Purpose

We report here an update to our previous systematic review on preventive interventions for post COVID-19 condition [15]. For this review, the following key question was addressed: Among people in the acute (symptom onset to 4 weeks) or early post-acute phase (4-8 weeks) of COVID-19, what are the effects of interventions to prevent post COVID-19 condition (PCC)?

## METHODS

The review followed a pre-defined, registered (PROSPERO (CRD42024513247) protocol and our reporting follows the Preferred Reporting Items for Systematic Reviews and Meta-analyses (PRISMA) framework [16]. The scope of the review was very similar to our original review with the main exception that all studies had to report on our primary outcome of prevalence of PCC. Wide awareness of the condition and the availability of a universal definition enabled us to rely for this update on this outcome rather than having to consider specific aspects/symptoms of the condition. Other outcomes (e.g., dyspnea, fatigue) were considered secondary, only extracted from studies also reporting on the primary outcome, and used mainly to help explain the findings on PCC prevalence. We also refined which secondary outcomes were of interest, to focus on the most common PCC symptoms as well as three key patient-important outcomes (i.e., health-related quality of life [HRQoL], hospital admission, return to work/education).

### Eligibility criteria

The population of interest included people of any age in the acute (0-4 weeks) and/or post-acute (5-8 weeks) phase of COVID-19. We did not limit inclusion based on method for confirming SARS-CoV-2 infection or whether re-infection during follow-up was ruled out. We included studies where ≤20% of participants were at 9-12 weeks in post-acute phase and studies reporting data for those at a combined timeframe of 0-8 weeks following infection. We excluded studies where most (≥80%) participants were in ICU during their acute infection. Specific populations of interest for examining variations of effects included: age (e.g., 0-17 vs. 18-65 vs. 65+ years), sex, socioeconomic status, race/ethnicity, population with poor access to interventions (e.g., rural dwelling), re-infection status, vaccination status (0 vs. full dosing vs. ≥1 boosters), SARS-Cov-2 variants, severity of acute infection (e.g., setting of acute care), pre and/or co-existing conditions (e.g., 0 vs. ≥2 conditions, pregnant, and immunocompromised).

The interventions of interest included: single and combinations of medications (including but not limited to steroids, antivirals, anti-inflammatory, immunomodulators, anticoagulation, antibiotics, metformin) and/or supplements (e.g., vitamins, minerals, probiotics); self-management advice; self-management programs, with or without tailoring or support groups; multidisciplinary care models/pathways with comprehensive centralized screening/triage, and, if indicated, assessment and direct treatment and/or referrals to community/primary care or specialist clinics; primary care treatment or referrals with/without screening/assessment; rehabilitation; single discipline interventions (e.g., psychology, psychiatry, cardiology, occupational therapy, physical activity, nutritional counselling); and other interventions (e.g., Indigenous health practices, Ayurvedic medicine, Chinese medicine, combinations of above). We excluded studies focused on vaccination (pre- or post-COVID infection) and post-ICU syndrome, and interventions used as primary prevention (e.g., masking), that is to prevent or reduce severity of infection. The interventions had to be provided to COVID-19 infected individuals at or before 8 weeks following diagnostic test or symptom onset (in ≥90% of participants) as a therapeutic and/or prophylactic measure. A minimum follow-up of 21 days after starting the intervention was required. Any country and/or setting were eligible for inclusion.

For the comparator, we considered usual medical care including supportive care for COVID-19 during acute phase (but not all patients receiving one treatment/medication), as well as no comparator (if no other study was found for the same intervention). We also considered including studies with an active control arm(s).

The primary outcome was PCC. Studies could use WHO criteria with specification about symptom persistence and attempt to rule out other diagnoses, but we also included studies where one or more PCC symptoms were present at 3 months or longer (in ≥75% of the sample) after an initial infection (or 2 months after hospital discharge). The symptoms used (often via checklists) for defining PCC could vary across studies but needed to capture at least fatigue, dyspnea and one or more aspects of cognitive function. We did not include studies that captured diagnoses of chronic condition or major acute sequelae (e.g., stroke, renal failure) as aspects of PCC (e.g., [17,18]). We considered studies defining PCC by documentation of PCC-related ICD/clinical codes or by at least two visits to a PCC clinic (to avoid counting as cases those that were referred/assessed and seen once but not having PCC) unless it was clear that a substantial proportion (e.g., >25%) of the documentation occurred before 3 months post-COVID. Author-defined recovery rates were also considered as indirect markers of PCC. These may have used binary outcomes (e.g., at least some limitations) from composite scores of validated scales covering multiple domains of generic HRQoL or functional capacity related to common PCC symptoms [e.g., < grade 2 on Post-COVID-19 Functional Status scale], as well as single questions about recovery to pre-infection health. We excluded studies only reporting on individual PCC symptoms and where it was clear that outcome ascertainment was performed under 3 months after the infection onset in ≥25% participants.

Secondary outcomes of interest included: fatigue; breathlessness/dyspnea; post-exertional malaise; HRQoL; psychopathology (i.e., anxiety, depression, post-traumatic stress disorder); cognitive impairment; hospital admission; return to work/education; any and serious adverse effects of the intervention (or number of withdrawals due to the adverse events).

We included randomized trials, quasi-randomized or experimental studies (e.g., controlled before-after, interrupted time-series, before-after studies), prospective and retrospective cohorts with a control group, and (if no comparative study was available for an intervention) case-series/uncontrolled cohorts if baseline and follow-up scores were reported. Studies had to have at least 30 participants. We included all nonrandomized studies regardless of whether they had a study design feature (e.g., matching) or used analysis to adjust for potential confounders, but this was considered during our risk of bias assessments. We included peer-reviewed articles, results in trial registration systems (if no report published), or pre-prints/reports (not peer-reviewed journal publications) from original quantitative research.

### Literature search and study selection

A research librarian updated our previous peer-reviewed searches on February 9, 2024. The search included a combination of controlled vocabulary and key words combining concepts of PCC, broad intervention/prevention terms as well as a comprehensive list of possible pharmacologic and nonpharmacologic interventions relevant for use during acute or post-acute phases of COVID-19, and a broad study design filter for trials and observational studies (excluding case reports and [from Embase] conference abstracts]). Minor revisions were made to our previous search, to add some intervention terms and precision for the PCC concept (**Supplementary file**). The search was limited to studies published in English and French. Searches were conducted via the Ovid platform in MEDLINE ALL and Embase with a date limit of May 2021 onwards (to allow for overlap with our previous search in July 2021). These databases now index preprint sites such as MedRxiv. We searched ClinicalTrials.gov for studies reporting results data over the past year. New to this update, we also performed forward citation searches in Scopus for studies citing those included from our database searches. Additionally, one reviewer scanned the reference lists of relevant reviews and the included studies for studies not found in our database searches. We uploaded the results of these searches to an EndNote (v. X9, Clarivate Analytics, Philadelphia, PA) library, removed duplicates, and imported them into DistillerSR (DistillerSR Inc., Ottawa, ON).

We performed study selection in DistillerSR using a two-step process, first by title and abstract (level 1) and then by full text (level 2). During level 1, we used DistillerAI which learns from human screening decisions and continually re-prioritizes records according to a predicted probability of inclusion. After screening 50% of citations (shown when using DistillerAI to allow for at least 95% recall of included studies based on dual review [19,20]) using a liberal accelerated method, whereby each title and abstract requires one reviewer to be included and two to be excluded, we modified our approach to rely on single reviewer screening for the remaining records. Any potentially relevant records were automatically moved to level 2. At level 2, two reviewers reviewed all full-text records independently and came to consensus, using input from the review lead or other reviewer for decisions when necessary. We contacted study authors by email (with 1-week response duration and no reminders) if additional information was needed to come to a final decision. Using standardized forms, all reviewers involved in each level piloted a random sample of records (using 100 records for level 1 and 20 records for level 2 screening) prior to beginning each stage.

### Data extraction and management

We developed standardized data extraction forms in Microsoft Office Excel (v. 2016, Microsoft Corporation, Redmond, WA) to collect relevant information from the included studies. All reviewers independently piloted the form on a sample of three studies to ensure accuracy and adequacy of the forms in capturing data and to make modifications if necessary. Following the pilot phase, one reviewer independently extracted data from the included studies, with verification for accuracy and completeness by another reviewer. Disagreements were resolved by discussion or by consulting a third reviewer.

From each study we extracted information related to the study characteristics (author, year, country, funding source, registration/protocol, design [considered by us as prospective or retrospective based on the method to ascertain the exposure/intervention], data collection dates), population characteristics (eligibility criteria, sample size, demographics, COVID-19 ascertainment method and timing, re-infections, vaccinations), setting and type of care for acute phase (out-patient, hospitalization [with % in ICU], mixed with out- and in-patients), intervention (providers, timing, components, dose, frequency, duration, delivery mode), comparator(s), follow-up length, analysis details (e.g., variables accounted for in analysis of nonrandomized studies), and outcomes (methods of ascertainment, definitions and scale range and directions, sample analyzed, number of events or mean change scores and variance in each group). We also extracted any author-reported subgroup analyses related to our specific populations of interest. We used figures to extract data if necessary, using PlotDigitizer. We contacted study authors by email to clarify missing or unclear information, or to obtain results matching our outcome definitions.

For randomized studies, we extracted and relied on crude events rates or (for continuous data) values by arm, when these data were reported; otherwise we used author-reported between-group statistics from unadjusted analyses. For observational studies, if multiple analyses were reported we extracted results from the most adjusted analysis. If data for outcomes were reported at multiple timepoints, we selected one data point closest to 3 months after infection. This was revised from our original protocol where we specified two timepoints of interest. If multiple outcomes were used by the authors that fit within our eligible PCC outcome, we chose the one closest to the WHO criteria (e.g., single-item questions about non-recovery rates were not used when a checklist of common PCC symptoms was provided). For secondary outcomes of interest, if multiple measures were reported for the same outcome, we prioritized data from measurement tools that were considered more valid or to best represent the outcome domain (e.g., validated measures over single item author-developed questions, risk for at least minimal anxiety versus a measure of mean anxiety scores) [21]. If more than two measurements were considered equal/interchangeable, we extracted all data and calculated an average of the values for analysis (e.g., anxiety and depression for our outcome of psychopathology) [22].

When there were multiple reports using data from the same source population (e.g., same database but differing dates or focusing on different interventions/outcomes), we avoided using data from more than one report in a single analysis and instead chose the data best fitting our timing (e.g., closest to 3-6 months to avoid long-term follow-up when some cases of PCC may have been resolved) and outcome (e.g., PCC via WHO vs. non-recovery rates), or, if all else was similar, the largest sample. We considered these separate studies but report on their association and which data was used from each.

### Risk of bias assessment

We assessed risk of bias of the included studies using the Cochrane ROB 2.0 tool for randomized and nonrandomized trials [23] and the JBI critical appraisal checklist for cohort studies [24]. Before performing the assessments, we piloted each tool on a sample of five studies. After piloting, two reviewers independently assessed the risk of bias for each study. Disagreements were resolved by discussion, or the involvement of a third reviewer if needed.

We assessed the risk of bias for objective (i.e., measured) and subjective (i.e., self-reported) outcomes separately. We considered the overall risk of bias for an outcome in a trial to be low if all domains/questions were at low risk of bias, having some concerns if no domains were at high risk of bias or we did not feel the study conclusions were impacted by the results (e.g., findings of little-to-no effect for objective outcomes), and high if one or more domains were assessed as being at high risk of bias and we believed the conclusions could have been impacted (e.g., findings of small benefit for subjective outcomes). For outcomes reported in observational studies, we considered inadequate adjustment for potential confounders (i.e., minimum adjustment for age, sex, severity of acute COVID-19 infection, and for receipt of other medications) and high attrition rates as the major variables of interest and rated studies with these concerns as high risk for bias; studies with concerns in other areas were rated as having some concerns. We used information from all publications related to any study, and from published or registered protocols, to inform our assessments. Final risk of bias assessments were incorporated into our assessments of the certainty of evidence using GRADE.

### Data manipulation and synthesis

For PCC prevalence and each secondary outcome, we grouped the studies by intervention type and timing (i.e., acute vs. post-acute), and acute-care setting (i.e., inpatient, outpatient, or mixed). For binary data, we used our calculated RR for randomized trials and the adjusted odds ratio (OR) from nonrandomized studies, if reported. When adjusted ORs were not reported we used crude data to calculate effects using a RR (if not pooled with other ORs) or an OR depending on the analysis. We assumed ORs are similar to RR for rare events (≤5%).

Within each timepoint and setting, we considered pooling data if there was enough similarity in interventions and outcome definitions regardless of the timing of the outcome assessment, keeping randomized studies separate from non-randomized and observational studies. For medications, our focus was on classes of drugs rather than specific drugs, though we reported findings as specific to a drug if that was used for all studies in the analysis. We used a pairwise random effects meta-analysis using the DerSimonian and Laird between-study variance estimator in Review Manager (RevMan v.5.3, Copenhagen: The Nordic Cochrane Centre, the Cochrane Collaboration, 2014). Data that were not appropriate for meta-analysis were synthesized descriptively. We also used data from studies not included in a meta-analysis but reporting on similar outcomes (e.g., only reporting direction of effect and/or p values), to help interpret findings from the meta-analysis. For all syntheses, a RR of 0.75-1.25 was considered a finding of little-to-no effect; 0.51-0.74 and 1.26 to 1.99 were small-to-moderate effects (for benefit or harm, respectively), and ≤0.50 or ≥2.00 were large effects. When the analysis used an OR and where the outcome probability was between 20% to 80% in the population studied, we converted the thresholds into ORs by taking the square root of each RR (i.e., ORs 0.56-1.56 for little-to-no effect, 0.26-0.55 and 1.57-3.96 for small-to-moderate effect, and ≤0.25 or ≥4.00 for large effect) [25]. We did not test for small study bias as there were no pooled data with 10 or more studies included in the synthesis.

For PCC prevalence, we performed sensitivity analyses by removing studies from any synthesis assessed as having high risk of bias, using the proxy outcome of non-recovery, and assessing the outcome at >6 months follow-up. We had planned to undertake between-study subgroup analyses for the specific populations of interest (see section on eligibility) but ended up having to rely on reports of within-study analyses undertaken by study authors.

### Certainty of evidence and interpreting findings

We assessed the certainty of evidence for the primary and secondary outcomes using GRADE [26,27]. At least two reviewers met and reached consensus through discussion for each outcome. Because we were assessing causation, certainty started at low for nonrandomized studies whereas randomized studies started at high certainty. We rated down for each domain (risk of bias, indirectness, inconsistency, imprecision, reporting biases) by 0, 1 or 2 levels depending on how serious the concerns were, i.e., how much overall conclusions appeared to be impacted by the domain. For indirectness, major considerations were given to whether outcome timing (i.e., outcome ascertainment outside of 3 to 12-month follow-up) and measurement (i.e., used proxy outcome of non-recovery) matched our target outcome domains. When there was a single small study (e.g., <100), we also had serious concerns about the directness/applicability of findings to a larger general population. For imprecision, we compared the 95% confidence intervals (CI) to our thresholds and rated down once or twice if one or both, respectively, of the CI limits crossed a threshold (unless the width of the CI was thought to be impacted mainly due to inconsistency). Statistical significance was not considered. For inconsistency, we rated down only if there were point estimates on either side of our thresholds; we did not rely on statistical measures of heterogeneity though report I^2^ values. When we included randomized and nonrandomized studies for a similar outcome, these were rated separately and we chose the findings with higher certainty for our conclusions. Our narrative descriptions of the findings use the terms “probably/likely”, “may/suggests” and “uncertain” when the evidence was of moderate, low and very low uncertainty, respectively [28]. Because our targets of certainty (and thus conclusions) were based on our thresholds and not the actual point estimates, findings of benefits should be interpreted as such, that is, showing at least a 25% reduction. Likewise, findings on no benefit should be interpreted as not meeting our thresholds for benefit or harm.

## RESULTS

We identified 3,298 unique records from all sources, and examined full texts of 412 papers (**Figure 1**). Twenty-four studies [29–52] (and one associated paper [53]) were included, including five randomized [32,41,42,44,48] and 19 non-randomized studies [29–31,33–40,43,45–47,49–52]. Two studies were carried over from the previous version of this review [40,46]; most of the previous reviewed studies did not report on PCC prevalence as per our revised methods. In two instances, studies using overlapping databases were included. In one case, the larger [49] study was used for the PCC outcome and one secondary outcome, and the smaller study [34] was used for subgroup analysis for PCC prevalence and other secondary outcomes. In the second instance, two studies were used for different interventions [39,45]; another relevant study [54] on this population was excluded based on our decision rules (see methods). The **Supplementary file** has a list of excluded studies, with reasons.

**Figure 1.**
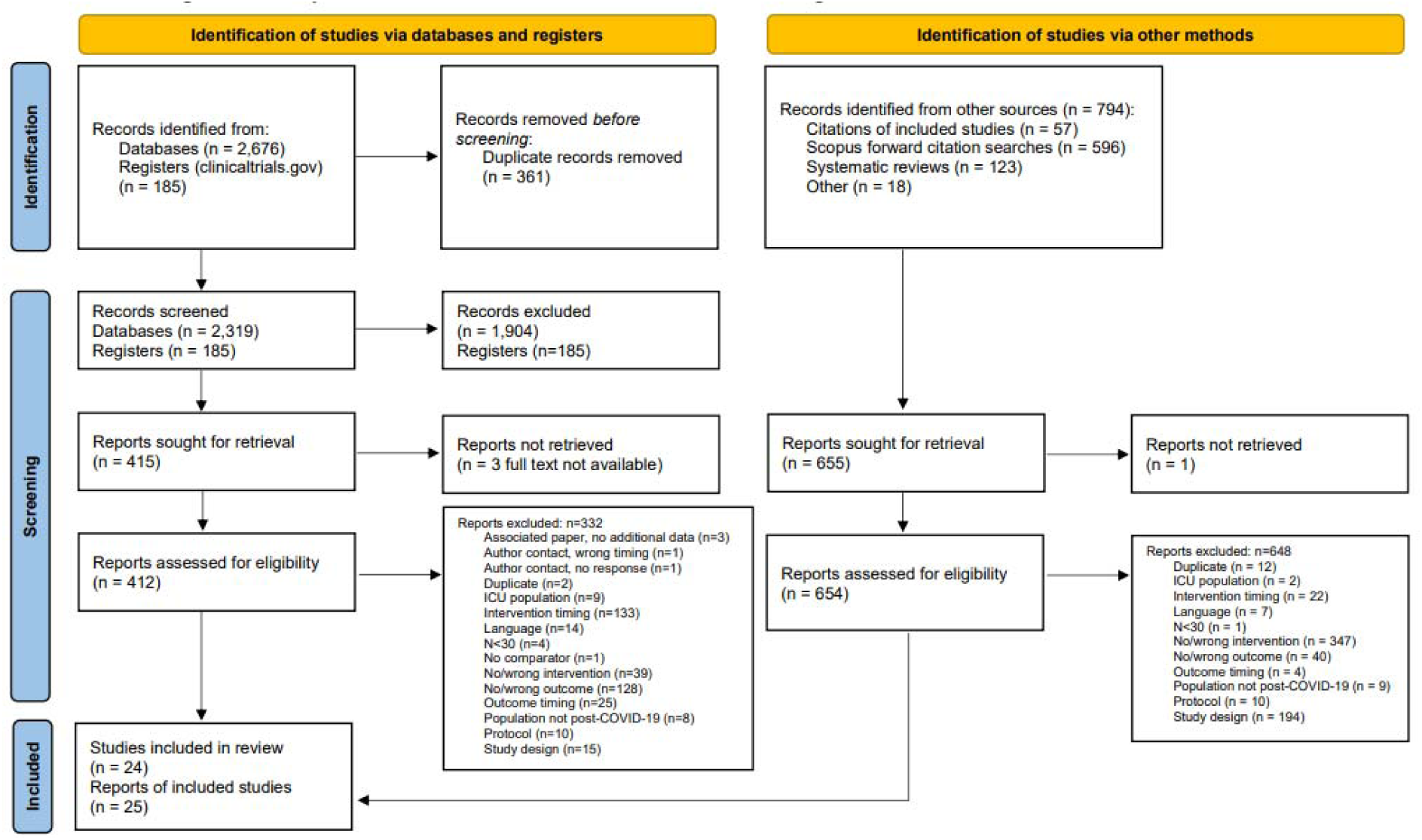
Literature flow

Across studies, all limited their sample to adults and the median age was 60 years (range: 29 to 74) and median sample size was 468 (range: 69 to 53,186). The acute-care setting was outpatient for 9 studies (one having 1% hospitalized) and inpatient in 15 studies (0% to 40% of inpatients receiving ICU care). Five studies were from the United States, three from the United Kingdom, three across multiple countries, two studies each were from China, Italy, the Netherlands, and Spain, and there were single studies from Ukraine, Finland, Brazil, Belgium, and France.

Nine studies were included for each of antivirals (5 in inpatients and 4 outpatients) and steroids (inpatients), five examined antibiotics (inpatients), four anticoagulants (in-patients), two hydroxychloroquine (inpatients)and monoclonal antibodies (in- and outpatients), and single studies examined anti-inflammatory agents (inpatients), probiotics (outpatients), platelet anti-aggregants (inpatients), zinc (inpatients), and (all outpatients) convalescent plasma, metformin, ivermectin, fluvoxamine, activity levels (≥150 minutes per week), assessment and referrals. All interventions except one (assessment and referrals) were initiated in the acute phase of COVID-19 (0-4 weeks after infection symptom onset). Two studies examined combinations of treatments. Secondary outcomes were reported in all five trials and in 12 nonrandomized studies [29,33–35,40,43,45–47,49,50,52].

Risk of bias assessments for PCC prevalence were low risk in three studies, some concerns in nine studies, and high risk in 11 studies; in one study risk varied by intervention (some concerns in the first intervention, and high risk in the second and third intervention). Tables with details of the study characteristics, risk of bias assessments, within-study subgroup analyses, and summary of findings for the secondary outcomes are reported in the **Supplementary file**.

### Post-COVID condition

Moderate certainty evidence was found from one randomized trial on the use of convalescent plasma versus control plasma for acute COVID-19 in the outpatient setting. Among individuals who received this intervention, it probably does not reduce the risk of PCC (**Table 1**). The timing of receiving convalescent plasma (≤ 5 versus >5 days) after acute COVID-19 infection did not impact findings (**Supplementary file)**.

**Table 1.**
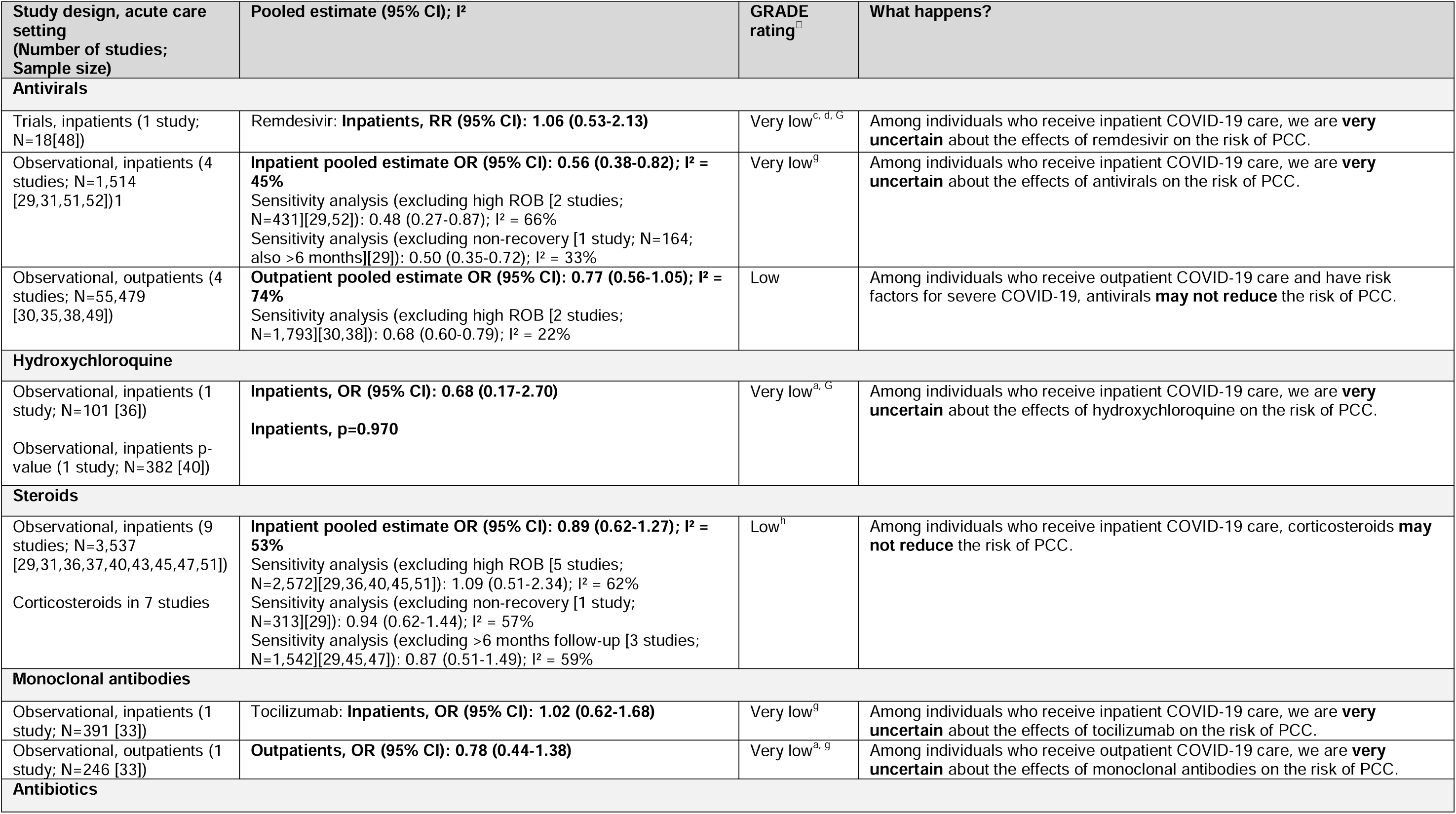
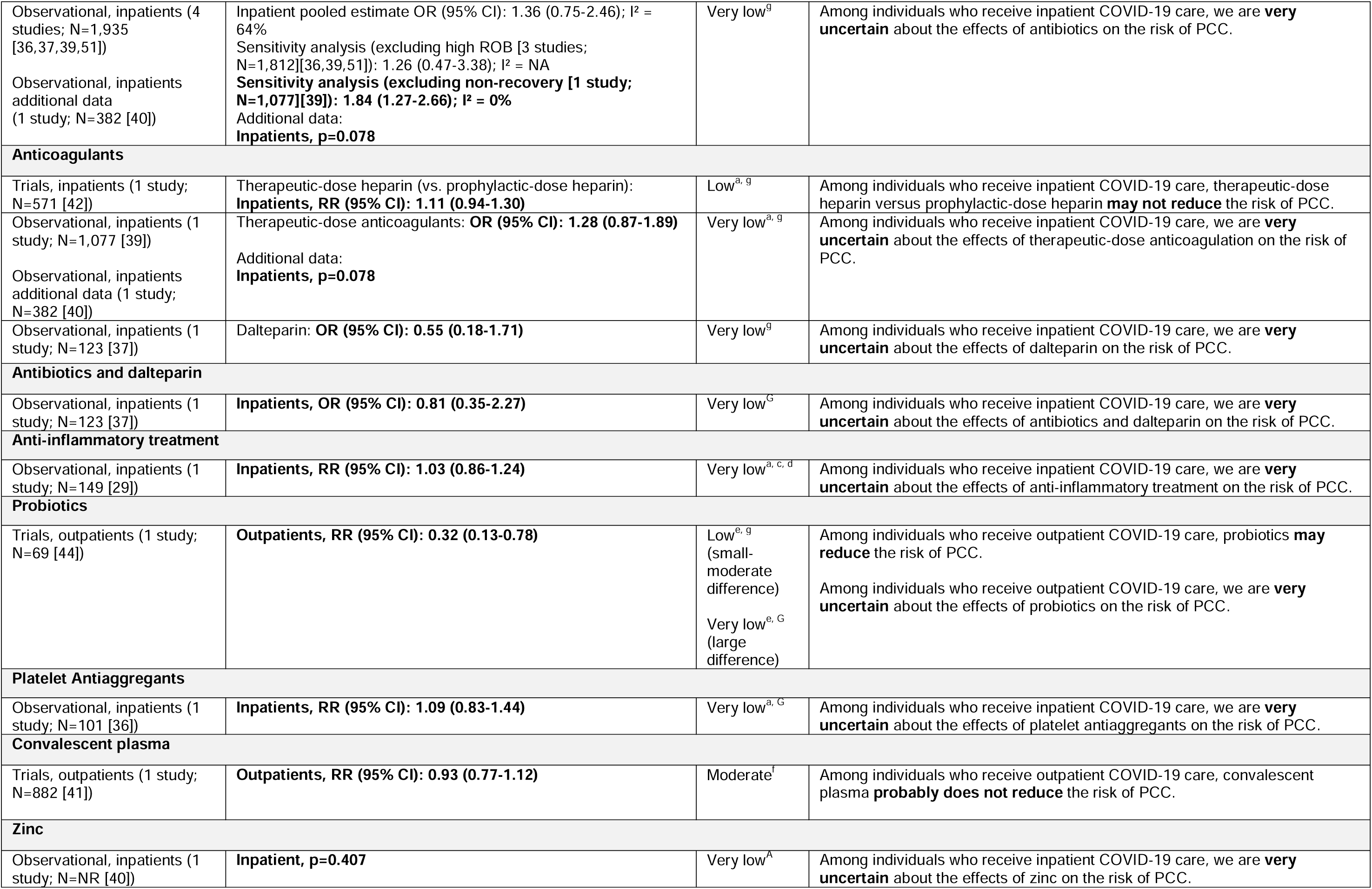
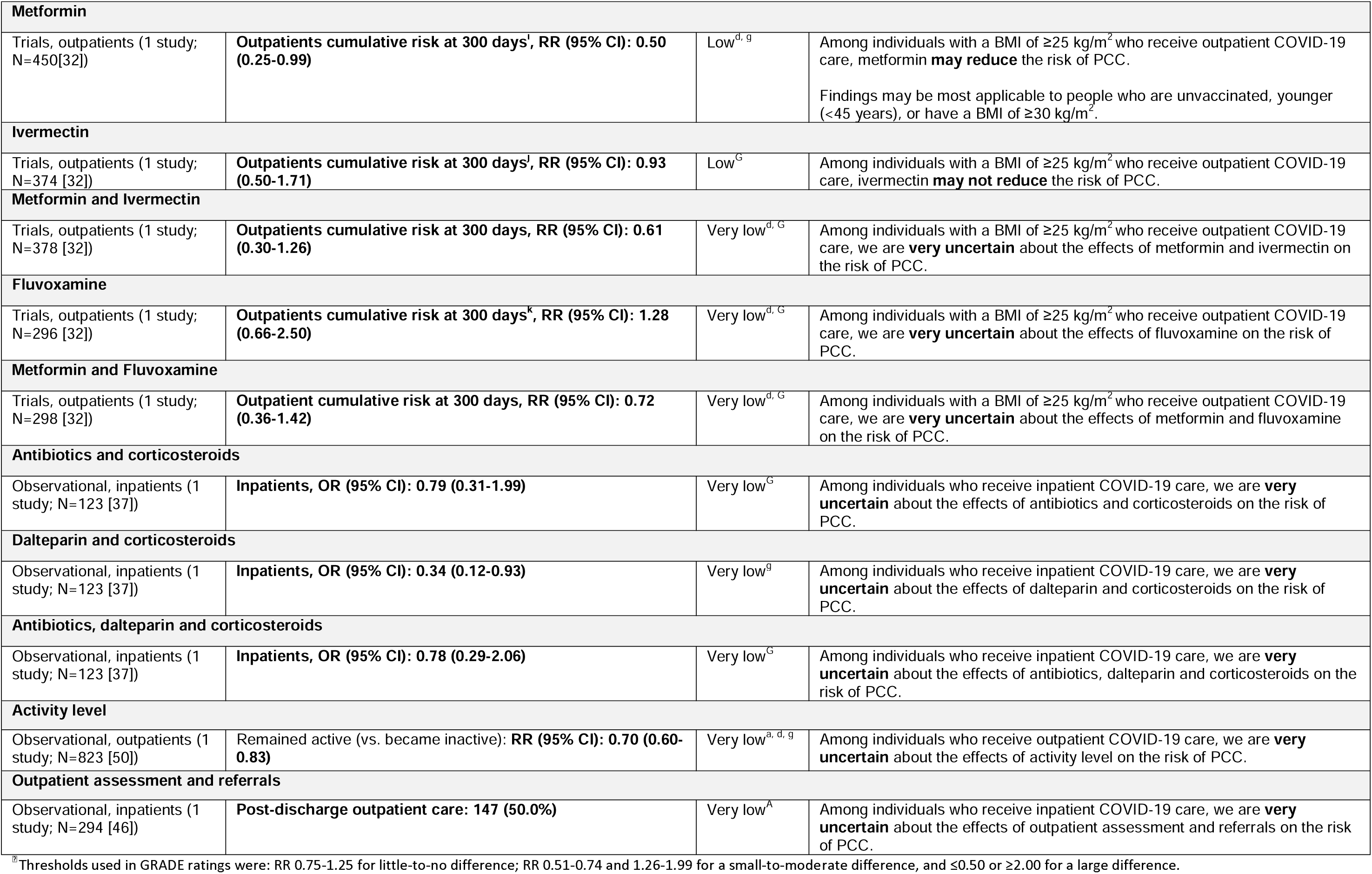
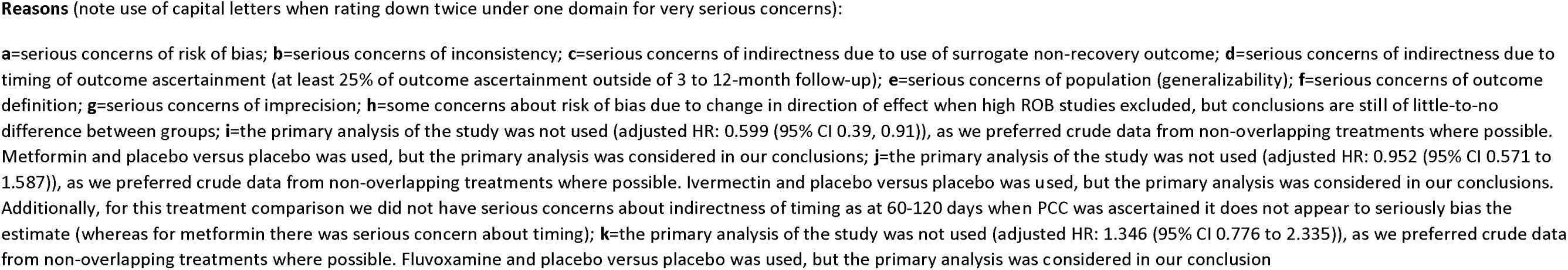
Summary of findings for prevention of Post-COVID condition, by intervention.

We found low certainty evidence for six interventions (**Table 1**). Four observational studies reported on the use of antivirals (in three specified as nirmatrelvir/ritonavir) in the outpatient setting for individuals that had risk factors for severe COVID-19. Among this population, antivirals may not reduce the risk of PCC (**Table 1**, **Figure 2**). Sensitivity analysis removing studies at high risk of bias found similar results. Findings for antiviral use in the inpatient setting were very uncertain mainly due to imprecision around our thresholds. If one thinks a 15% risk reduction is important (i.e., use of RR 0.85 instead of 0.75 as the threshold for a benefit) the effects would be considered low certainty for a reduction because there would no longer be imprecision. Nine observational studies found that receiving steroids (“steroids” in 1, corticosteroids in 7, and glucocorticoids in 1 study) during inpatient care may not reduce the risk of PCC (**Table 1**, **Figure 3**). Several sensitivity analyses were done to remove high risk of bias studies, surrogate non-recovery outcomes, and outcome ascertainment over 6 months post-acute infection. Findings were similar across all sensitivity analyses. From one trial assessing probiotic use among outpatients, probiotics may reduce the risk of PCC to a small extent. If we were to consider the point estimate (RR 0.32) as a large effect based on our thresholds, we would be very uncertain about a large effect of probiotics, due to imprecision. A trial assessing multiple interventions among individuals with a BMI of ≥25 kg/m^2^ who received outpatient COVID-19 care, found that metformin may reduce the risk of PCC. Based on subgroup analysis (**Supplementary file**), findings may be most applicable to people who are unvaccinated, younger (<45 years), or have a BMI of ≥30 kg/m^2^. Conclusions did not appear to vary based on COVID variant during acute infection (i.e., omicron versus pre-omicron). This trial also evaluated the use of ivermectin, with low certainty that ivermectin may not reduce the risk of PCC. Lastly, a trial of inpatients provided low certainty evidence that therapeutic-dose heparin did not reduce PCC compared with prophylactic-dose heparin.

**Figure 2.**
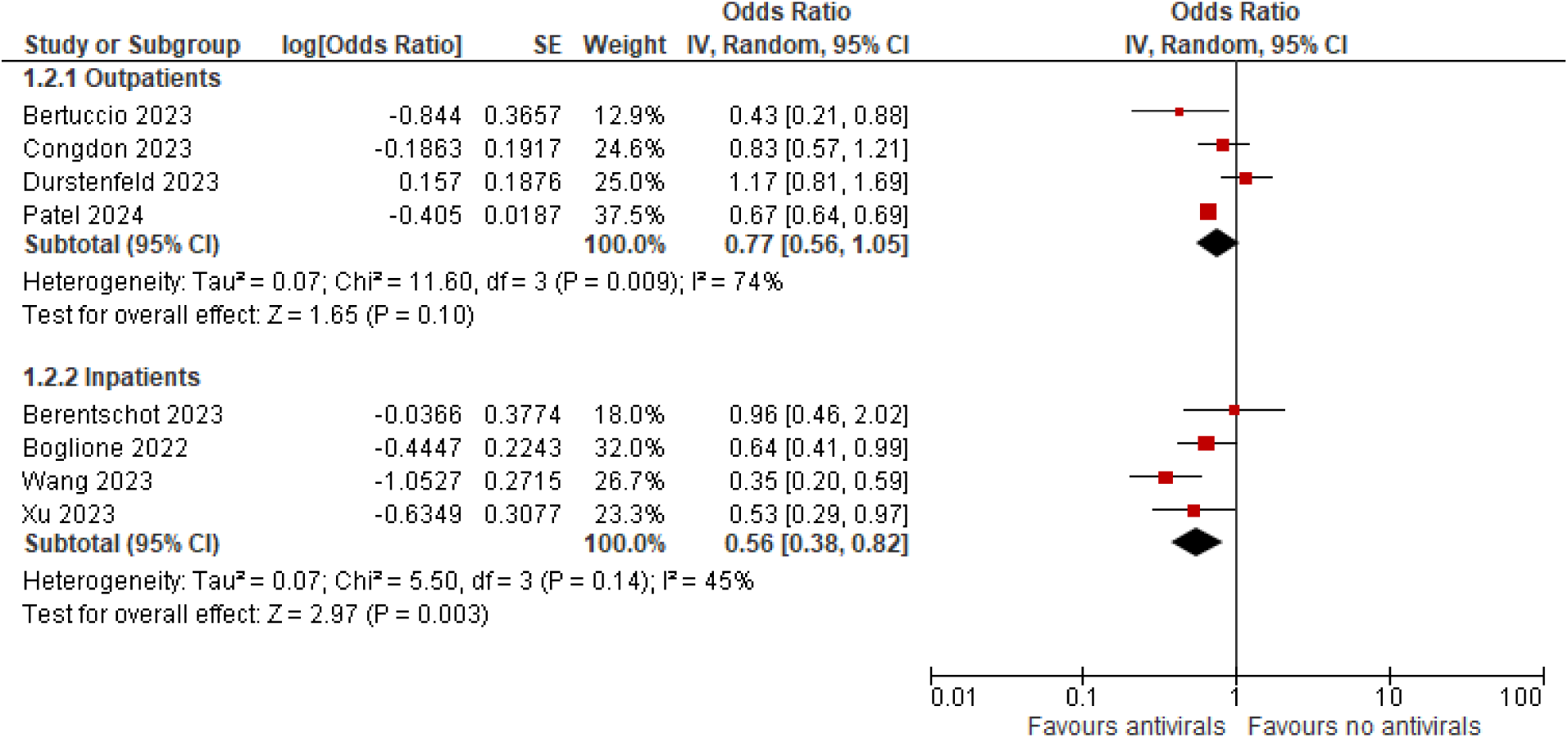
Meta-analyses for PCC prevalence from use of antivirals during acute-phase COVID-19 among outpatients and inpatients

**Figure 3.**
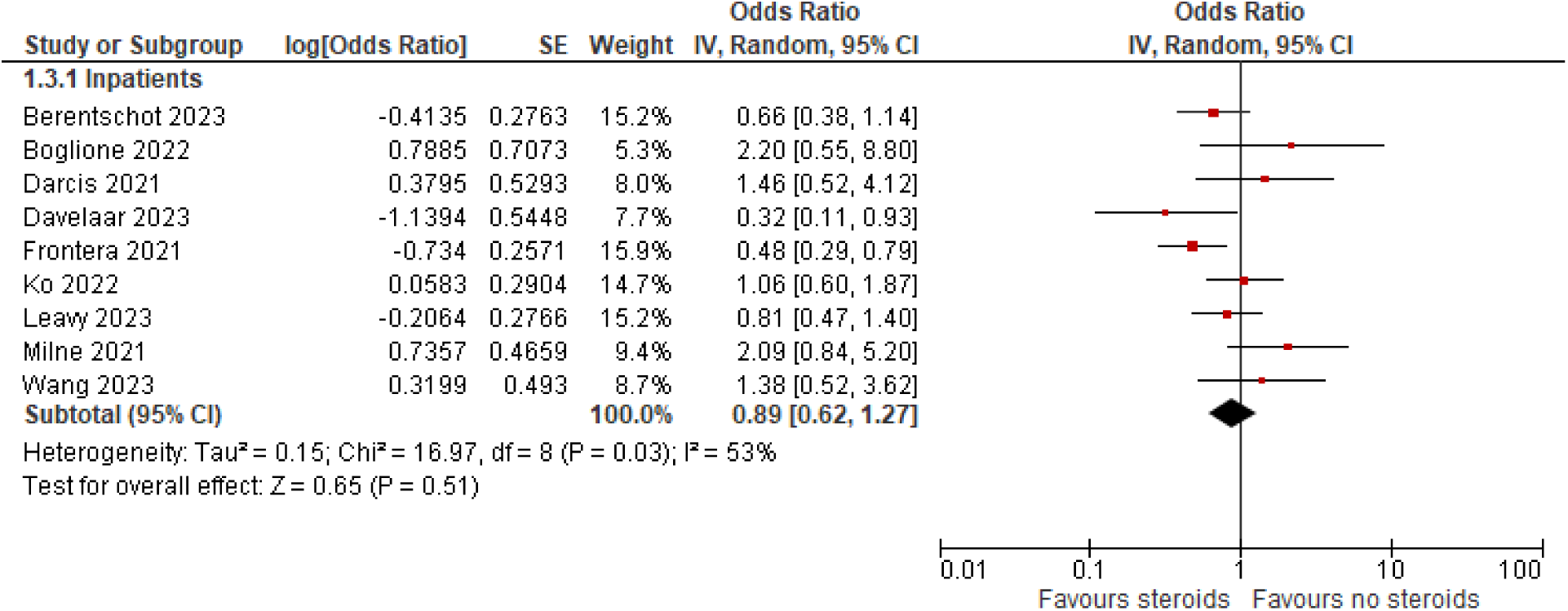
Meta-analyses for PCC prevalence from use of steroid during acute-phase COVID-19 among inpatients

### Secondary outcomes

The only moderate certainty evidence was found from a trial of individuals who received inpatient acute COVID-19 care (n=410) [42]. Therapeutic-versus prophylactic-dose heparin probably does not improve HRQoL, measured with the EQ-5D-5L index tool (MD (95% CI): 0.12 (-3.22 to 3.46) [range 0-100]).

Across the remaining studies, 13 outcomes had low certainty evidence.

### Antivirals

A single observational study (n=53,186) of individuals with risk factors for severe COVID-19 evaluated nirmatrelvir/ritonavir use for outpatient care [49]. Results indicated that this antiviral may reduce the risk of psychopathology (RR (95% CI): 0.71 (0.69 to 0.74)). Findings from an observational study (n=24,490) investigating outpatient care with nirmatrelvir/ritonavir found that it may reduce the risk of all-cause hospitalizations (RR (95% CI): 0.54 (0.45 to 0.65)) [34]. Two observational studies (N=24,990) of outpatients evaluating nirmatrelvir/ritonavir found that it may not reduce the risk of fatigue (RR (95% CI): 0.91 (0.79 to 1.05)) [34,35].

One trial (n=181) of inpatients assessed the use of remdesivir on HRQoL with the EQ-VAS tool [48]. Remdesivir may not reduce the risk of worsened (not defined) HRQoL (OR (95% CI): 0.83 (0.49 to 1.40)). The same trial (n=181) also found that remdesivir may reduce to a small extent the risk of moderate-to-severe prolonged general malaise following even light exertion (RR (95% CI): 0.41 (0.15 to 1.12). If considering these results as a large effect based on our thresholds, we would be very uncertain about the effects due to added imprecision.

### Other interventions

A trial (n=571) of inpatients receiving acute COVID-19 treatment found that therapeutic-dose versus prophylactic-dose heparin may reduce to a small extent the risk of cognitive impairment (RR (95% CI): 0.49 (0.22 to 1.07)) [42]. If we were to consider this a large effect based on our thresholds, we would be very uncertain about the effects due to added imprecision. This trial also found that therapeutic-dose versus prophylactic-dose heparin may not reduce the risk of fatigue (RR (95% CI): 1.05 (0.85 to 1.30)). One trial assessed several interventions among individuals with a BMI of ≥25 kg/m^2^ who received outpatient COVID-19 care [32]. For the primary intervention of metformin (n=569), the study found that it may not reduce the risk of serious adverse events (no events per group). Similar findings that study interventions may not reduce the risk of serious adverse events were also found for fluvoxamine (n=319), ivermectin (n=406), metformin and fluvoxamine (n=336) and metformin and ivermectin (n=401) (no events in any comparison).

There were no findings with moderate or low certainty for the secondary outcomes of breathlessness/dyspnea, return to work, and any adverse events, as well as other interventions not listed above. The **Supplementary file** contains summary of findings for all secondary outcomes.

## DISCUSSION

From this update to our previous systematic review on interventions to prevent PCC, there is no longer very low certainty across all interventions. For preventing PCC, convalescent plasma used for patients receiving outpatient acute-COVID-19 care probably does not reduce the risk of PCC. Low certainty evidence was found that probiotics and metformin may reduce, and ivermectin and antivirals may not reduce PCC among outpatients, and steroids and therapeutic heparin may not reduce PCC among inpatients. All certainty ratings are based on conclusions of small effects (≤25% risk reduction). For outpatient antiviral treatment, though overall PCC may not be reduced findings suggested a small reduction in psychopathology. Likewise, for inpatient use of antivirals where there may be at most a very small benefit for PCC there may be a small reduction in moderate-to-severe prolonged general malaise following even light exertion. Therapeutic-dose versus prophylactic-dose heparin may reduce to a small extent the risk of cognitive impairment. All examined medications were initiated during the acute phase of infection, and there was still very low certainty for several treatments such as monoclonal antibodies, anti-inflammatory agents, zinc, and outpatient assessment and referral.

### Comparison with other reviews

Based our group’s regular surveillance on research related to PCC, we are not aware of many reviews on prevention. One systematic review with a search in July 2023 [55] focused on pharmacological treatments received during acute SARS-CoV-2 infection. Five studies on antivirals were reviewed, with the authors reporting conflicting results among studies among in- and out-patients. One large study [18] did not meet our eligibility criteria since PCC was not defined as a symptomatic syndrome in the study. The authors included two studies on dexamethasone and concluded findings of some benefit, though the studies would not have met our criteria due to their use of specific symptoms or a data collection period before to 3 months post-infection. They examined the same trial we did on metformin/ivermectin/fluoxetine, and report benefit from using metformin. The authors’ search included several terms for antivirals, but other interventions would have required to be found by the term “acute therapy”. Further, the review authors did not use thresholds for effect or rate their certainty in the evidence. As previously mentioned, our previous review found very low certainty across all interventions, and several studies were not eligible for this update [15]. The national guidelines underway for Canada (https://can-pcc.recmap.org/recommendations) have used the same trial as ours on metformin for their topic about PCC treatment (hence finding the evidence indirect), and have at the time of this report not yet included recommendations about this treatment for prevention. All published recommendations related to prevention have so far focused on primary prevention (i.e., masking) which was not of interest for this review.

### Limitations of evidence

Very little evidence had moderate or high certainty evidence, often due to use of observational study designs, high risk of bias, and small samples leading to imprecision. Very few studies reported on nonpharmacological interventions or on interventions initiated in the post-acute period when persisting symptoms may become apparent. None evaluated the impact of some highly relevant interventions such as rehabilitation or multidisciplinary care models.

### Strengths and limitations of review

We employed rigorous methods to locate, appraise, analyze and interpret findings across a wide variety of potential interventions to prevent PCC. We used a structured approach to assess effects based on differences in populations and intervention timing, and evaluated whether findings were robust when considering risk of bias and factors potentially leading to indirectness in terms of the PCC outcome. We rated the certainty of evidence and interpreted findings in light of whether or not they were thought to provide important effects. There are some potential limitations. We employed AI software to prioritize records during title/abstract screening and did not use dual reviewer screening for our entire search

[19]; we may have missed a few studies from using this approach though our hand-searching of reference lists and reviews as well as the Scopus searches would help circumvent this. Studies that assessed long-term follow-up after acute-care treatments for PCC (e.g., as a secondary/tertiary study endpoint) but did not report this or similar concepts in their titles or abstract may have been missed by our search. We included only English and French language articles and may have not included some studies published in other languages. While for many topics our search date would be quite recent, research on COVID-19 emerges quickly and other studies will have likely been published but not reviewed. We are aware of reports on one matched retrospective cohort study on remdesivir in in-patients (n=432; showing a protective effect at 18 months follow-up) [56], an analysis of two trials on fluvoxamine (n=521 after about 30% attrition; showing a protective effect at 3-months on poor recovery based on ≤60% recovered) [57], and a trial on a 2-week multi-nutrient supplement use (n=246; showing no benefit at 180-days) [58]. Apart from this study on multinutrient supplement, the others would be considered to offer indirect evidence for our primary outcome of PCC. Because all eligible studies had to report our primary outcome, other reports about our secondary outcomes will exist; our purpose was not to systematically assess findings for all secondary outcomes (e.g., individual symptoms) but rather the use these findings together with those on overall PCC incidence to help explain the findings, such as helping to describe the mechanisms of effect (or lack thereof) on overall PCC.

### Conclusions

To a small degree, PCC may be prevented by using probiotics and metformin during the acute phase of COVID-19 among outpatients. It is probably not prevented among outpatients from use of convalescent plasma during acute care. Findings could be strengthened by more studies for these and several other interventions where there was low or very low certainty evidence. Effects from interventions used among in-patients and within the post-acute phase are uncertain at this time. Evidence on commonly recommended interventions including rehabilitation or multidisciplinary care is lacking.

## DECLARATIONS

### Ethics approval and consent to participate

Not applicable.

### Consent for publication

Not applicable.

### Availability of data and materials

The data generated during this study are available within the manuscript or its supplementary files.

## Data Availability

The data for the primary outcome generated during this study are available within the manuscript or its supplementary files. Any other data can be obtained by request of the corresponding author.

## Acknowledgments.

The authors thank Kate Merucci from Health Canada for developing the original search and Taline Ekmekjian from the Public Health Agency of Canada for revising and running the searches for this update.

