## Supplementary file for "Preventive interventions for post Covid-19 condition: systematic review update"

Table S1. Characteristics table of included studies

| First Author, Year; Country; Study Design[ǂ]; Timing of Infection<br><br>Recruitment setting and participant eligibility<br><br>Risk of bias for primary outcome | Type of Acute Care<br>Average age (years); % Female; % Vaccinated<br><br>COVID-19 Illness Severity<br><br>Eligible Population All Having Post-acute Symptoms (Y/N) | Intervention/Exposure Group (IG): Description & Timing<br><br>Control Group (CG)<br><br>N analyzed/enrolled | PCC Outcome Description & Timing<br>Main Results<br><br>Available Sub-group Data | Reported findings related to secondary outcomes (bold indicates data used for synthesis where >1) |
|---|---|---|---|---|
| **Outpatients/mixed population with treatment initiated in acute phase** | | | | |
| Kolesnyk, 2024<br><br>Ukraine<br><br>RCT<br><br>November 2021 to June 2022<br><br>General population in primary care and outpatient practices; 18–65-year-old men or non-pregnant women with symptomatic COVID-19 lasting no longer than 5 days and confirmed by antigen or PCR testing<br><br>ROB: Low | Outpatients (0% hospitalized)<br><br>Median 45.0; 53%; NR<br><br>Mild-to-moderate<br><br>N | IG: Dietary supplement with probiotics (1 pill / day x 28 days); ≤5 days after positive test (n=34/36)<br><br>CG: Placebo (n=35/37)<br><br>* Other medication received during acute phase in both groups included: NSAIDs, Antitussives, Nasal decongestants, Throat antiseptics, Antispasmodics (only in the placebo group), Antidiarrheal, Antiplatelet, herbs<br><br>Total: 69 analyzed/73 randomized | Post-COVID Functional<br><br>Scale (PCFS) range 0-4; 13 weeks after COVID-19 positivity<br><br>Score 0 (absence of any functional limitation):<br><br>IG: 29/34 (85.3%)<br><br>CG: 19/35 (54.3%)<br><br>P=0.008<br><br>None | Fatigue (single item)<br><br>Cognitive impairment (attention deficit, single item)<br><br>Psychopathology (average of anxiety and depression,<br><br>single items)<br><br>Any adverse event (withdrawals due to AEs) |
| Rocha, 2023<br><br>Brazil<br><br>Retrospective cohort | Outpatients (0% hospitalized)<br><br>NR; 59%; NR | IG: Remained physically active (≥150 mins/week) after COVID-19 (n=388/NR) | Symptom checklist (≥1 symptom of 19 [not necessarily absent before COVID-19] focused on physical symptoms); 6-10 months after positive test | Fatigue (single item)<br><br>Breathlessness/Dyspnea (dyspnea, single item)<br><br>Cognitive impairment (average of memory loss |

| First Author, Year; Country; Study Design[i]; Timing of Infection<br><br>Recruitment setting and participant eligibility<br><br>Risk of bias for primary outcome | Type of Acute Care<br>Average age (years); % Female; % Vaccinated<br><br>COVID-19 Illness Severity<br><br>Eligible Population All Having Post-acute Symptoms (Y/N) | Intervention/Exposure Group (IG): Description & Timing<br><br>Control Group (CG)<br><br>N analyzed/enrolled | PCC Outcome Description & Timing<br>Main Results<br><br>Available Sub-group Data | Reported findings related to secondary outcomes (bold indicates data used for synthesis where >1) |
|---|---|---|---|---|
| December 2020 to March 2021<br><br>≥18-year old Brazilian participants who received a COVID-19 diagnosis through RT-PCR testing, experienced COVID-19 symptoms, and received medical care in Rio Grande<br><br>ROB: High | Mild-to-moderate<br><br>N | CG1: Became physically inactive after COVID-19 (n=435/NR)<br><br>CG2: Remained physically inactive (<150 mins/week) after COVID-19 (n=2,096/NR)<br><br>Total: 823 analyzed in comparison of interest (2,919 analyzed overall)/3,550 eligible | IG: 142/388 (36.5%)<br><br>CG1: 226/435 (51.9%)<br><br>CG2: 1,041/2,096 (49.7%)<br><br>IG vs. CG1: aPR NR<br><br>Additional analyses (not of interest):<br><br>IG vs. CG2: aPR 0.80 (0.63, 0.99)<br><br>CG1 vs. CG2: aPR 0.97 (0.80, 1.17)<br><br>None | and attention loss, single items) |
| Gebo, 2023<br><br>US<br><br>Secondary analysis of a double-blind RCT (75% of original sample)<br><br>June 2020 to October 2021<br><br>Symptomatic adult outpatients (≥18 years old) with acute SARS-CoV-2 infection from 23 sites. Analysis restricted to those with blood drawn at screening, day 14, and day 90, and complete symptom data on day 90 | Outpatients (0% hospitalized)<br><br>Median 43.0; 57.4%; 15.8% and 6.2% fully and partially vaccinated<br><br>Mild-to-moderate<br><br>N | IG: COVID-19 convalescent plasma (CCP); ≤9 days of symptom onset (n=445/592)<br><br>CG: Control plasma; ≤9 days of symptom onset (n=437/589)<br><br>Total: 882 analyzed/1,225 randomized | Symptom checklist (≥1 symptom of 17) at 90 days post-transfusion<br><br>Among those with 90 days symptom data and blood samples:<br><br>IG: 142/445 (31.9%)<br><br>CG: 150/437 (34.3%)<br><br>Subgroups: by timing of receiving convalescent plasma | None |

| First Author, Year; Country; Study Design[ǂ]; Timing of Infection<br><br>Recruitment setting and participant eligibility<br><br>Risk of bias for primary outcome | Type of Acute Care<br>Average age (years); % Female; % Vaccinated<br><br>COVID-19 Illness Severity<br><br>Eligible Population All Having Post-acute Symptoms (Y/N) | Intervention/Exposure Group (IG): Description & Timing<br><br>Control Group (CG)<br><br>N analyzed/enrolled | PCC Outcome Description & Timing<br>Main Results<br><br>Available Sub-group Data | Reported findings related to secondary outcomes (bold indicates data used for synthesis where >1) |
|---|---|---|---|---|
| ROB: Low | | | | |
| Durstenfeld, 2023<br><br>US<br><br>Prospective matched cohort<br><br>March to August<br><br>2022<br><br>Vaccinated, non-hospitalized, nonpregnant adults (≥18 years old) who were registered in an online cohort study (COVID-19 Citizen Science). Outcomes were ascertained only for those who responded to surveys about Long COVID symptoms in November and December 2022<br><br>ROB: High | Outpatients (0% hospitalized)<br><br>Median 55.0; 66%; 100%<br><br>Mild-to-moderate, but eligible for antivirals<br><br>N | IG: Oral nirmatrelvir/ritonavir; ≤30 days of positive test (n=353/988)<br><br>CG: No exposure, not all eligible/high-risk (n=1,258/3,696)<br><br>Total: 1,611 analyzed/4,684 enrolled | Symptom checklist (≥1 symptom of 19 plus "other"); 5.4 ± 1.3 months after infection<br><br>IG: 57/353 (16.1%)<br><br>CG: 176/1,258 (14.0%)<br><br>Propensity-score adjusted (and adjusted for additional factors) aOR: 1.17 (0.81, 1.69)<br><br>Propensity-score adjusted aOR: 1.15 (0.80, 1.64); P=0.45<br><br>Analysis restricted to those at high-risk for severe infection i.e. eligible for treatment, by<br><br>sensitive definition<br><br>(n=968 treated and n=3326 untreated):<br><br>OR: 1.11 (0.77, 1.59)<br><br>by specific definition (n=890 treated and n=2,840):<br><br>OR: 1.12 (0.78, 1.60)<br><br>Analysis among those at high risk for severe COVID-19 (propensity-matching):<br><br>0.032 (-0.017, 0.081); P=0.20 | None |

| First Author, Year; Country; Study Design[ǂ]; Timing of Infection<br><br>Recruitment setting and participant eligibility<br><br>Risk of bias for primary outcome | Type of Acute Care<br>Average age (years); % Female; % Vaccinated<br><br>COVID-19 Illness Severity<br><br>Eligible Population All Having Post-acute Symptoms (Y/N) | Intervention/Exposure Group (IG): Description & Timing<br><br>Control Group (CG)<br><br>N analyzed/enrolled | PCC Outcome Description & Timing<br>Main Results<br><br>Available Sub-group Data | Reported findings related to secondary outcomes (bold indicates data used for synthesis where >1) |
|---|---|---|---|---|
| | | | 10.5% vs. 10.2% reported 1+ severe PCC symptom | |
| Congdon, 2023<br><br>US<br><br>Retrospective cohort<br><br>January to July 2022<br><br>Adult COVID-19 patients ≥18 years old who met the criteria to be prescribed nirmatrelvir/ritonavir under the Food and Drug Administration's EUA<br><br>ROB: Some concerns | Outpatients (1% hospitalized)<br><br>Mean 50.6 (SD: 15.2); 70%; 67.6% primary series+booster, 24.6% primary series alone<br><br>Mild-to-severe, but eligible for antivirals<br><br>N | IG: Nirmatrelvir/ritonavir full/partial dose (out of 10 total doses) received during acute infection (n=250)<br><br>CG: No exposure, but eligible (n=250)<br><br>Total: 500 analyzed/500 enrolled | Symptom checklist (≥1 new or worsened symptoms of 11); 4 months (120-150 days) after positive test<br><br>IG: 110/250 (44%)<br><br>CG: 125/250 (50%)<br><br>aOR: 0.83 (0.57, 1.2), P=0.31<br><br>None | Fatigue (generalized fatigue, single item)<br><br>Breathlessness/Dyspnea (dyspnea, single item)<br><br>Cognitive impairment (brain fog, single item) |
| Bertuccio, 2023<br><br>Italy<br><br>Retrospective cohort<br><br>April 2021 to March 2022 (94.3% during Omicron)<br><br>Outpatient adults (>18 years old) with a positive SARS-CoV-2 nasopharyngeal swab and mild-to moderate COVID-19 within the prior 5 (for oral antivirals) or 7 (for intravenous short-course remdesivir or mAbs) days; and | Outpatients (0% hospitalized)<br><br>Median 67.0 (IQR 54–76); 48.4%; 81.3% with ≥ 2 doses<br><br>Mild-to-moderate, but all having ≥ 1 risk factor for severe illness<br><br>N | IG1: Anti-SARS-CoV-<br><br>2 monoclonal antibodies (bamlanivimab, etesevimab, casirivimab, imdevimab, sotrovimab), ≤7 days of symptom onset (n=141)<br><br>IG2: Oral antivirals (nirmatrelvir/ritonavir, molnupiravir, or short-course remdesivir); ≤5-7 days of symptom onset (n=77)<br><br>CG: patients eligible for IG1 or IG2 who refused treatment (n=105) | CDC symptom list (≥1 symptom, unclear if present before COVID-19), 3 months post symptom-onset<br><br>IG1: 42/141 (29.8%)<br><br>IG2: 16/77 (20.8%)<br><br>CG: 36/105 (34.3%)<br><br>IG1 vs. CG: aOR 0.78 (0.44, 1.36)<br><br>IG2: vs. CG: aOR 0.43 (0.21, 0.87) | None |

| First Author, Year; Country; Study Design[i]; Timing of Infection<br><br>Recruitment setting and participant eligibility<br><br>Risk of bias for primary outcome | Type of Acute Care<br>Average age (years); % Female; % Vaccinated<br><br>COVID-19 Illness Severity<br><br>Eligible Population All Having Post-acute Symptoms (Y/N) | Intervention/Exposure Group (IG): Description & Timing<br><br>Control Group (CG)<br><br>N analyzed/enrolled | PCC Outcome Description & Timing<br>Main Results<br><br>Available Sub-group Data | Reported findings related to secondary outcomes (bold indicates data used for synthesis where >1) |
|---|---|---|---|---|
| reported presence of at least one risk factors for COVID-19 progression<br><br>ROB: High | | Total: 323 analyzed/649 enrolled | Sensitivity analysis did not show any differences between the three periods, where prevalent variants were respectively Alpha, Delta and Omicron (data not shown). | |
| Patel, 2024<br><br>US (TriNetX)<br><br>Retrospective matched cohort<br><br>December 2021-<br><br>December 2022<br><br>Non-hospitalized, vaccinated adult patients (≥18 years old) with preexisting CVD (i.e., presence of ≥1 comorbidity including hypertension, hyperlipidemia, type 2 diabetes mellitus, ischemic heart disease, cerebrovascular disease, cardiomyopathy/heart failure, arrhythmia) who were vaccinated and subsequently developed COVID-19<br><br>ROB: Some concerns | Outpatients (0% hospitalized)<br><br>Mean 62.4 (SD: 14.5); 61%; 100%<br><br>Mild-to-moderate<br><br>N | IG: Nirmatrelvir-ritonavir, ≤5 days of diagnosis (n=26,593)<br><br>CG: No exposure (n=26,593), not all eligible<br><br>Total: 53,186 analyzed/53,186 enrolled | ≥1 symptom based on the CDC and WHO definition (included a list of the constitutional, cardio-respiratory, gastrointestinal, musculoskeletal, and nervous system, and/or mood/cognitive disorder symptoms), 90-180 days post index infection<br><br>IG: 6,692/26,593 (25.1%)<br><br>CG: 8,910/26,593 (33.5%)<br><br>Propensity score matching OR: 0.67 (0.64, 0.69); P < 0.001 | Psychopathology (Anxiety/mood disorders, ICD codes) |

| First Author, Year; Country; Study Design[ǂ]; Timing of Infection<br><br>Recruitment setting and participant eligibility<br><br>Risk of bias for primary outcome | Type of Acute Care<br>Average age (years); % Female; % Vaccinated<br><br>COVID-19 Illness Severity<br><br>Eligible Population All Having Post-acute Symptoms (Y/N) | Intervention/Exposure Group (IG): Description & Timing<br><br>Control Group (CG)<br><br>N analyzed/enrolled | PCC Outcome Description & Timing<br>Main Results<br><br>Available Sub-group Data | Reported findings related to secondary outcomes (bold indicates data used for synthesis where >1) |
|---|---|---|---|---|
| Chuang, 2023<br><br>Taiwan (TriNetX)<br><br>Retrospective matched cohort<br><br>January 1 and July 31, 2022<br><br>Adult population (>18 years old) with more than two visits to Health Care Organizations (HCOs) that tested SARS-CoV-2<br><br>ROB: Low | Outpatients (0% hospitalized)<br><br>Mean 56.0; 57.3%; NR<br><br>NR<br><br>N | IG: Nirmatrelvir plus ritonavir, ≤5 days of diagnosis (n=12,245)<br><br>CG: No exposure (n=12,245), not all eligible<br><br>Total: 24,490 analyzed/24,490 enrolled | Overlapping population with Patel 2024, thus used for the subgroup data for the primary outcome and secondary outcomes<br><br>Subgroups: age, sex, and vaccination<br><br>status (i.e., none vs. ≥1 dose >2 weeks before the diagnosis of COVID-19) | Fatigue (ICD codes)<br><br>Cognitive impairment (ICD codes)<br><br>Psychopathology (anxiety/depression, single item, used Patel 2024 data)<br><br>Hospitalization (**all-cause hospitalizations, ICD codes**; ER visits) |
| Bramante, 2023<br><br>US<br><br>RCT (factorial design 2x3)<br><br>December 2020 to January 2022<br><br>Adults 30-85 years old with overweight or obesity (BMI ≥25 kg/m² or ≥23 kg/m² for people identifying as Asian or Latino) who had COVID-19 symptoms for fewer than 7 days and confirmed COVID-19 infection within 3 days before enrolment, with no known previous SARS-CoV-2 infection | Outpatients (0% hospitalized)<br><br>Median 45.0 (IQR 37–54); 56.1%; 55% with primary series<br><br>Symptomatic<br><br>N | IG1: Metformin+placebo (n=221)<br><br>IG2: Ivermectin+placebo (n=189)<br><br>IG3: Fluvoxamine+placebo (n=144)<br><br>IG1+1G2 (n=189)<br><br>IG1+IG3 (n=154)<br><br>CG: Matched placebo for each drug (n=229 for IG1, 185 for IG2, 152 for IG3, 189 for IG1+IG2, 144 for IG1+IG3)<br><br>all ≤7 days after symptom onset | Self-reported diagnosis of PCC by medical provider (with EHR confirmation), diagnoses 10 months (300 days) from randomization<br><br>Crude data for single drugs + placebo chosen over main analysis in paper:<br><br>IG1 vs. CG1: 11/221 (5.0%) vs. 23/229 (10.0%)<br><br>IG2 vs. CG2: 18/189 (9.5%) vs. 19/185 (10.3%)<br><br>IG3 vs. CG3: 17/144 (11.8%) vs. 14/152 (9.2%) | Serious AEs (self-report in diaries) |

| First Author, Year; Country; Study Design[I]; Timing of Infection<br><br>Recruitment setting and participant eligibility<br><br>Risk of bias for primary outcome | Type of Acute Care<br>Average age (years); % Female; % Vaccinated<br><br>COVID-19 Illness Severity<br><br>Eligible Population All Having Post-acute Symptoms (Y/N) | Intervention/Exposure Group (IG): Description & Timing<br><br>Control Group (CG)<br><br>N analyzed/enrolled | PCC Outcome Description & Timing<br>Main Results<br><br>Available Sub-group Data | Reported findings related to secondary outcomes (bold indicates data used for synthesis where >1) |
|---|---|---|---|---|
| ROB: Some concerns | | *Every participant received a pill that looked like metformin (either active metformin or exact-matching metformin placebo). For combination of interventions, the second pill was either ivermectin or exact-matching ivermectin placebo; or fluvoxamine or exact-matching fluvoxamine placebo.<br><br>Total: 1,126 analyzed/1,323 enrolled in mITT analysis (1,431 originally randomized) | IG1+IG2 vs. CG: 11/189 (5·8%) vs. 18/189 (9.5%)<br><br>IG1+IG3 vs. CG: 13/154 (8.4%) vs. 17/144 (11.8%)<br><br>Main analysis from paper:<br><br>Metformin vs. placebo: aHR: 0.599 (0.39, 0.91)<br><br>Ivermectin vs. placebo: aHR: 0.952 (0.571, 1.587)<br><br>Fluvoxamine vs. placebo: aHR: 1.346 (0.776, 2.335)<br><br>Subgroups (available for the first three main analyses from the paper): sex, age group (<45 vs. ≥45y), timing of the first dose of IG (≤3 vs ≥4 days from symptom onset), SARS variants, vaccination status | |
| **Hospitalized population with treatment initiated in acute phase** | | | | |
| Darcis, 2021<br><br>Belgium<br><br>Retrospective cohort<br><br>March to October 2020 (discharge date) | Hospitalized (26% ICU admission)<br><br>Mean 60.5 (SD: 13.9); 36.7%; NR<br><br>Moderate-to-severe | IG1: Hydroxychloroquine (n=88)<br><br>IG2: Corticosteroids (n=25)<br><br>IG3: Antibiotics (n=98)<br><br>IG4: Platelet antiaggregants (n=24)<br><br>Timing NR | ≥1 persistent symptom (from a list of 20 symptoms), 3 months post-discharge<br><br>IG1 vs. CG1: 61/88 (69.3%) vs.10/13 (76.9%)<br><br>IG2 vs. CG2: 19/25 (76.0%) vs. 52/76 (68.4%)<br><br>IG3 vs. CG3: 70/98 (71.4%) vs. 1/3 (33.3%) | None |

| First Author, Year; Country; Study Design[ł]; Timing of Infection<br><br>**Recruitment setting and participant eligibility**<br><br>**Risk of bias for primary outcome** | Type of Acute Care<br>**Average age (years); % Female; % Vaccinated**<br><br>**COVID-19 Illness Severity**<br><br>**Eligible Population All Having Post-acute Symptoms (Y/N)** | Intervention/Exposure Group (IG): Description & Timing<br><br>**Control Group (CG)**<br><br>**N analyzed/enrolled** | PCC Outcome Description & Timing<br>**Main Results**<br><br>**Available Sub-group Data** | Reported findings related to secondary outcomes (bold indicates data used for synthesis where >1) |
|---|---|---|---|---|
| Patients ≥16 years of age admitted to the University Hospital of Liège with moderate-to-severe confirmed COVID-19<br><br>ROB: High | N | CG: did not receive the IG in each comparison (n=13 for IG1, 76 for IG2, 3for IG3, 77 for IG4)<br><br>Total: 101 analyzed/199 enrolled | IG4 vs. CG4: 18/24 (75%) vs. 53/77 (68.8%)<br><br>None | |
| Milne, 2021<br><br>UK<br><br>Prospective cohort<br><br>April to August 2020<br><br>Patients ≥18 years of age admitted to a single UK hospital with confirmed COVID-19 pneumonia, with an oxygen requirement during their admission<br><br>ROB: Some concerns | Hospitalized (11% ICU admission)<br><br>Median 60.0 (IQR: 53-72 in IG, 51-68 in CG); 31%; NR<br><br>Moderate-to-severe<br><br>N | IG: Dexamethasone (orally 6mg/day for up to 10 days or duration of hospitalization, received following the pre-print of the RECOVERY trial [June 2020], or as part of randomisation to that trial), during hospitalization (n=39)<br><br>CG: No dexamethasone (n=48)<br><br>Total: 87 analyzed/198 recruited | Prevalence of ongoing symptoms (total # and as a rate from a list 17 symptoms), 8 months post-admission<br><br>IG: 16/39 (41.0%)<br><br>CG: 12/48 (25.0%)<br><br>IG: n=73 symptoms, 1.9 symptoms/patient (n per group not reported)<br><br>CG: n=152 symptoms, 3.2 symptoms/patient (n per group not reported)<br><br>P=0.01<br><br>None | Fatigue (single item)<br><br>Psychopathology (MCS from SF-36; range 0-100, higher score=less mental disability) |
| Berentschot, 2024<br><br>Netherlands<br><br>Retrospective cohort | Hospitalized (40% ICU admission)<br><br>Median 60.0 (IQR: 53-68); 30.3%; 1.2% | IG1: Steroids only (n=211)<br><br>IG2: Anti-inflammatory treatment (n=47)<br><br>IG3: Antivirals (n=62) | Non-recovery status based on self-reported recovery assessed with the COVID-19 Core Outcome Measure for recovery (dichotomized into completely recovered and not completely recovered [comprising mostly recovered, somewhat recovered, half recovered, and not recovered at all]), median 730.0 (IQR: 726.0-739.0) days post-discharge | Fatigue (FAS score, range: 10-50, higher score=more fatigue) |

| First Author, Year; Country; Study Design[i]; Timing of Infection<br><br>Recruitment setting and participant eligibility<br><br>Risk of bias for primary outcome | Type of Acute Care<br>Average age (years); % Female; % Vaccinated<br><br>COVID-19 Illness Severity<br><br>Eligible Population All Having Post-acute Symptoms (Y/N) | Intervention/Exposure Group (IG): Description & Timing<br><br>Control Group (CG)<br><br>N analyzed/enrolled | PCC Outcome Description & Timing<br>Main Results<br><br>Available Sub-group Data | Reported findings related to secondary outcomes (bold indicates data used for synthesis where >1) |
|---|---|---|---|---|
| July 2020 to October 2021<br><br>Patients ≥18 years of age hospitalized for COVID-19 and within 6 months post-discharge. Included patients who had completed at least one of our outcomes<br><br>of interest at the 2-year follow-up<br><br>ROB: High | Moderate-to-severe<br><br>N | All during hospitalization<br><br>CG: No steroids, anti-inflammatory treatment, or antivirals, may have received (hydroxy)chloroquine (n=102)<br><br>Total: 502 analyzed/502 enrolled (N eligible NR) | IG1: 144/211 (68.2%)<br><br>IG2: 37/47 (78.7%)<br><br>IG3: 47/62 (75.8%)<br><br>CG: 78/102 (76.5%)<br><br>Between-groups adjusted P=0.23 | Breathlessness/Dyspnea (**dyspnea, mMRC score ≥1**; 6-MWT)<br><br>Cognitive failure (CFQ score; range 0-100, higher score=more cognitive failure)<br><br>HRQoL (EQ-5D-5L index; range 0-1, higher score=higher quality of life) |
| Greenstein, 2024<br><br>US and Spain<br><br>Open-label RCT<br><br>NR<br><br>Patients admitted to one of 34 hospitals in the US and Spain for COVID-19 (noncritically ill patients who were not receiving ICU-level care at enrollment) and were expected to be hospitalized for ≥72 hours | Hospitalized (0.0% ICU)<br><br>Mean 61.0; NR; NR<br><br>Noncritically ill (i.e., absence of critical care–level organ support at enrollment)<br><br>N | IG: Therapeutic-dose heparin for up to 14 days or until recovery/discharge (n=289) vs. CG: usual care with prophylactic-dose heparin (n=282)<br><br>IG2: Antiplatelet (aspirin or others) agents (n=163)<br><br>IG3: Anticoagulant therapies (n=447)<br><br>IG4: Steroids (N=462)<br><br>IG5: Remdesivir (N=381)<br><br>All during hospitalization | Self-reported presence of any symptom (from a list of 6 symptoms), at 3 months post-enrollment<br><br>IG1: 153/ 289 (53%)<br><br>CG: 135/282 (47.9%)<br><br>aOR: 1.19 (95% CI: 0.85, 1.66), P=0.30<br><br>IG2: 80/163 (49%)<br><br>IG3: 221/447 (49.4%) | Fatigue (single item)<br><br>Breathlessness/Dyspnea (average of shortness of breath at rest and shortness of breath with exertion, single items)<br><br>Cognitive impairment (single item)<br><br>Psychopathology (EQ-5D-5L subscore, moderate to extreme anxiety/depression)<br><br>HRQoL (**EQ-5DL index; range 0-100, higher** |

| First Author, Year; Country; Study Design[ǂ]; Timing of Infection<br><br>Recruitment setting and participant eligibility<br><br>Risk of bias for primary outcome | Type of Acute Care<br>Average age (years); % Female; % Vaccinated<br><br>COVID-19 Illness Severity<br><br>Eligible Population All Having Post-acute Symptoms (Y/N) | Intervention/Exposure Group (IG): Description & Timing<br><br>Control Group (CG)<br><br>N analyzed/enrolled | PCC Outcome Description & Timing<br>Main Results<br><br>Available Sub-group Data | Reported findings related to secondary outcomes (bold indicates data used for synthesis where >1) |
|---|---|---|---|---|
| ROB: High | | Total: 571 analyzed/727 randomized | IG4: 235/462 (50.9%)<br><br>IG5: 191/381 (50.1%)<br><br>None | **score=higher quality of life**; EQ-VAS) |
| Boglione 2022<br><br>Italy<br><br>Retrospective cohort<br><br>March 2020 to January 2021<br><br>Patients affected by<br><br>COVID-19 and hospitalized at Saint Andrea Hospital, and followed in a 'post-COVID ambulatory' care for at least 6 months after discharge<br><br>ROB: Some concerns | Hospitalized (13.8% ICU admitted)<br><br>Median 65.0 (IQR: 56-75.5); 22%; NR<br><br>N | IG1: remdesivir (n=163), < 10 days of symptom onset<br><br>IG2: corticosteroids (n=390), timing NR<br><br>CG: no remdesivir (n=286) for IG1, no corticosteroids (n=59) for IG2 (could have received other medications)<br><br>Total: 435 analyzed/449 enrolled | The level of functional status and the PCC severity reported using the PCFS which assigns the presence of significant long COVID for values<br><br>>2. Grades 3–4 were referred to significant limitations in everyday life with important functional limitations<br><br>Crude proportion NR<br><br>aOR, IG1 vs. CG: 0.641 (0.413, 0.782); P<0.001<br><br>aOR, IG2 vs. CG: 2.2 (0.55, 4.9); P-value NR<br><br>None | None |
| Cardona-Pascual, 2023<br><br>Spain<br><br>Retrospective matched cohort | Hospitalized (17% ICU admission)<br><br>Mean 58.9 (SD: 13.2); 36.5%; NR | IG: Tocilizumab, either with 600mg (400mg for weight<75kg) followed by a second dose of 400mg 12h apart, or a unique dose of 600mg (400mg for weight<75kg); during acute infection (n=129) | Patient/clinic reported persistent COVID-19 symptoms (≥1 symptom, not present prior to infection), 6 months after discharge | Breathlessness/Dyspnea (breathless/dyspnea, single item) |

| First Author, Year; Country; Study Design[ǂ]; Timing of Infection<br><br>Recruitment setting and participant eligibility<br><br>Risk of bias for primary outcome | Type of Acute Care<br>Average age (years); % Female; % Vaccinated<br><br>COVID-19 Illness Severity<br><br>Eligible Population All Having Post-acute Symptoms (Y/N) | Intervention/Exposure Group (IG): Description & Timing<br><br>Control Group (CG)<br><br>N analyzed/enrolled | PCC Outcome Description & Timing<br>Main Results<br><br>Available Sub-group Data | Reported findings related to secondary outcomes (bold indicates data used for synthesis where >1) |
|---|---|---|---|---|
| March to April 2020<br><br>Discharged SARS-CoV-2 patients from Valid' Hebron Barcelona Campus Hospital after surviving the first wave in 2020. All patients had to meet the criteria for tocilizumab therapy in accordance with the Spanish Agency of Medicines and Medical Devices<br><br>ROB: Some concerns | N | CG: No tocilizumab, but eligible (n=262)<br><br>All treated with standard pharmacological protocol, including antiviral drugs (lopinavir plus ritonavir twice a day), hydroxychloroquine 400mg/day and antibiotic prophylaxis (azithromycin and ceftriaxone).<br><br>Total: 392 analyzed/405 enrolled | IG: 47/129 (36.5%)<br><br>CG: 82/269 (31.3%)<br><br>Propensity score matching aOR: 1.02 (0.62, 1.68)<br><br>None | Psychopathology (anxiety/depression, single item)<br><br>Cognitive impairment (single item) |
| Davelaar, 2023<br><br>The Netherlands<br><br>Retrospective cohort<br><br>March 2020 to September 2021<br><br>Adult patients (≥18 years old) who were hospitalized due to COVID-19 symptoms confirmed using a PCR test | Hospitalized (% ICU NR)<br><br>Mean 62.1 (SD: 9.5); 38.2%; NR<br><br>31% "High" severity admission<br><br>N | IG1: Antibiotics, as prophylaxis or on suspicion of a bacterial infection, ceftriaxone 2000 mg was primarily used (n=89)<br><br>IG2: Anticoagulants (dalteparin), standard for most in acute phase (n=100)<br><br>IG3: Corticosteroids (mostly dexamethasone), if indicated (n=73)<br><br>IG1+IG2 (n=71)<br><br>IG1+IG3 (n=53)<br><br>IG2+IG3 (n=64) | Self-report using multiple questions requiring limitations in health, worse health than previous year, interference with daily life, and at least one of mental health, fatigue, or dyspnea symptoms, 6 months after discharge<br><br>IG1 vs. CG1: 24/89 (27.0%) vs. 9/34 (26.5%)<br><br>a OR: 1.26 (0.47, 3.39)<br><br>IG2 vs. CG2: 25/100 (25.0%) vs. 8/23 (34.8%) aOR: 0.55 (0.18, 1.71)<br><br>IG3 vs. CG3: 16/73 (21.9%) vs. 17/50 (34.0%) aOR: 0.32 (0.11, 0.90) | None |

| First Author, Year; Country; Study Design[ł]; Timing of Infection<br><br>Recruitment setting and participant eligibility<br><br>Risk of bias for primary outcome | Type of Acute Care<br>Average age (years); % Female; % Vaccinated<br><br>COVID-19 Illness Severity<br><br>Eligible Population All Having Post-acute Symptoms (Y/N) | Intervention/Exposure Group (IG): Description & Timing<br><br>Control Group (CG)<br><br>N analyzed/enrolled | PCC Outcome Description & Timing<br>Main Results<br><br>Available Sub-group Data | Reported findings related to secondary outcomes (bold indicates data used for synthesis where >1) |
|---|---|---|---|---|
| ROB: Some concerns | | IG1+IG2+IG3 (n=46)<br><br>Variable doses and durations, per protocol; during acute phase<br><br>CG: did not receive the IG in each comparison (n= 34 for IG1, 23 for IG2, 50 for IG3, 52 for IG1+IG2, 70 for IG1+IG3, 59 for IG2+IG3, 77 for IG1+IG2+IG3)<br><br>Total: 123 analyzed/123 enrolled | IG1+IG2 vs CG: 18/71 (25.4%) vs.15/52 (28.8%) aOR: 0.81 (0.35, 2.27)<br><br>IG1+IG3 vs. CG: 13/53 (24.5%) vs. 20/70 (28.6%)<br><br>aOR: 0.79 (0.31, 1.99)<br><br>IG2+IG3 vs. CG: 13/64 (20.3%) vs. 20/59 (33.9%)<br><br>aOR: 0.34 (0.12, 0.93)<br><br>IG1+IG2+IG3 vs. CG: 11/46 (23.9%) vs. 22/77(28.6%)<br><br>aOR: 0.78 (0.29, 2.06)<br><br>None | |
| Leavy, 2023 (PHOSP study)<br><br>UK<br><br>Retrospective matched cohort<br><br>February 2020 to March 2021 (discharge dates)<br><br>Adults admitted to hospital for COVID-19 and meeting current guideline recommendations for dexamethasone treatment; not on immunosuppressants | Hospitalized (% ICU NR)<br><br>Mean 58.6 (SD: 12.2); 35.4%; NR<br><br>Moderate-to-critical (all COVID-19 requiring at least oxygen supplementation; 18% severe)<br><br>N | IG: Corticosteroids (any systemic, oral or intravenous), during their hospital admission (n=738)<br><br>CG: No corticosteroid (n=489)<br><br>Total: 1,227 analyzed/1,888 enrolled | Presence of any symptoms at 1 year after discharge (long list of symptoms)<br><br>IG: 657/738 (89.1%)<br><br>CG: 424/489 (86.6%)<br><br>P=0.508<br><br>None | Fatigue (**FACIT fatigue score; range 0-52, higher score=less fatigue**; Fatigue VAS score)<br><br>Breathlessness/Dyspnea (**airway obstruction, FEV1/FVC <0.7;** FEV1 % predicted <80%; FVC % predicted <80%; breathlessness VAS score, dyspnea-12 score)<br><br>Cognitive impairments (mild cognitive |

| First Author, Year; Country; Study Design[ǀ]; Timing of Infection<br><br>**Recruitment setting and participant eligibility**<br><br>**Risk of bias for primary outcome** | **Type of Acute Care**<br>**Average age (years); % Female; % Vaccinated**<br><br>**COVID-19 Illness Severity**<br><br>**Eligible Population All Having Post-acute Symptoms (Y/N)** | **Intervention/Exposure Group (IG): Description & Timing**<br><br>**Control Group (CG)**<br><br>**N analyzed/enrolled** | **PCC Outcome Description & Timing**<br>**Main Results**<br><br>**Available Sub-group Data** | **Reported findings related to secondary outcomes (bold indicates data used for synthesis where >1)** |
|---|---|---|---|---|
| ROB: High | | | | impairment or worse, MoCA score <12)<br><br>Psychopathology (**probably general anxiety disorder, GAD score >8**; PCL-5 score, PHQ-8 score)<br><br>HRQoL (EQ-5D-5L; range 0-1, higher score=higher quality of life) |
| Evans, 2021 (PHOSP study)<br><br>UK<br><br>Retrospective cohort<br><br>March 5 to November 30, 2020 (discharge dates)<br><br>Patients ≥18 years old discharged from one of 53 National Health Service hospitals after admission for confirmed or clinician diagnosed COVID-19 (patients required to attend a research visit 2-7 months post discharge) | Hospitalized (% ICU NR)<br><br>Mean 58.0 (SD: 13.0); 36%; NR<br><br>17% very severe, 21% severe, 17% moderate, 46% mild<br><br>N | IG1: Systemic steroids, oral or intravenous, during hospitalization (n=301)<br><br>IG2: Antibiotics, during hospitalization (n=841)<br><br>IG3: Therapeutic dose anticoagulants, during hospitalization (n=321)<br><br>CG: did not receive the IG in each comparison (n=776 for IG1, 236 for IG2, 756 for IG3)<br><br>Total: 830 analyzed/1,170 enrolled | Patient-perceived recovery (i.e., fully recovered from COVID-19), median of 5·9 months (IQR 4·9–6·5) after discharge<br><br>Crude proportions NR<br><br>IG1 vs. CG, aOR: 1.02 (0.70,1.49), P=0.909<br><br>IG2 vs. CG, aOR: 1.21 (0.77, 1.90), P=0.410<br><br>IG3 vs. CG, aOR: 0.78 (0.53, 1.15), P=0.202<br><br>None | None |

| First Author, Year; Country; Study Design[i]; Timing of Infection<br><br>**Recruitment setting and participant eligibility**<br><br>**Risk of bias for primary outcome** | Type of Acute Care<br>**Average age (years); % Female; % Vaccinated**<br><br>**COVID-19 Illness Severity**<br><br>**Eligible Population All Having Post-acute Symptoms (Y/N)** | Intervention/Exposure Group (IG): Description & Timing<br><br>**Control Group (CG)**<br><br>**N analyzed/enrolled** | PCC Outcome Description & Timing<br>**Main Results**<br><br>**Available Sub-group Data** | Reported findings related to secondary outcomes (bold indicates data used for synthesis where >1) |
|---|---|---|---|---|
| ROB: High | | | | |
| Ko, 2022<br><br>France<br><br>Retrospective cohort<br><br>February to December 2020 (discharge dates)<br><br>Hospitalized adult patients with COVID-19, who were discharged from the Amiens Picardie University Hospital, France and had face-to-face assessment 4 months after admission<br><br>ROB: Some concerns | Hospitalized (37% ICU admission)<br><br>Mean 65.0 (SD: 14.0); 42%; NR<br><br>N | IG1: Corticosteroids, 6mg (n=104)<br><br>IG2: Antivirals (lopinavir/ritonavir/remdisivir (n=61)<br><br>IG3: Hydroxychloroquine (n=45)<br><br>All administered based on clinical decisions and during hospitalization<br><br>CG: did not receive the IG in each comparison 9n=202 for IG1, 245 for IG2, 261 for IG3)<br><br>Total: 306 analyzed/454 eligible for follow-up | Self-reported at least one current symptom, 4 months after admission<br><br>IG1 vs. CG: 66/104 (63.5%) vs. 127/202 (62.8%)<br><br>aOR: 1.06 (0.60, 1.88), P=0.84 (accounting for other treatments received)<br><br>IG2 and IG3: did not have any effect on prolonged symptoms (data NR)<br><br>Subgroups: ICU subpopulation, patients with non-invasive ventilation, and those receiving mechanical ventilation | Breathlessness/Dyspnea (dyspnea, single item) |
| Nevalainen, 2022<br><br>Finland<br><br>Open-label RCT<br><br>July 2020 to January 2021<br><br>Adults ≥18 years old with a PCR-confirmed | Hospitalized (11% ICU admission)<br><br>Mean 58.3 (SD: 13.4); 36%; NR<br><br>73% received dexamethasone | IG: Intravenous remdesivir once daily until discharge or a maximum of 10 days, 89% ≤2 days after hospitalization (n=98 analyzed/114 randomized)<br><br>CG: No remdesivir (n=83 analyzed/94 randomized) | Recovery from COVID-19 (fully or largely), 1 year after discharge<br><br>IG: 83/98 (85%)<br><br>CG: 71/83 (86%)<br><br>Unadjusted RR: 0.94 (0.47, 1.90) | Fatigue (moderate/severe, single item)<br><br>Breathlessness/Dyspnea (exertional dyspnea, mmRC score 2-4)<br><br>Cognitive impairments (average of moderate/severe problems with memory |

| First Author, Year; Country; Study Design[ǂ]; Timing of Infection<br><br>**Recruitment setting and participant eligibility**<br><br>**Risk of bias for primary outcome** | Type of Acute Care<br>**Average age (years); % Female; % Vaccinated**<br><br>**COVID-19 Illness Severity**<br><br>**Eligible Population All Having Post-acute Symptoms (Y/N)** | Intervention/Exposure Group (IG): Description & Timing<br><br>**Control Group (CG)**<br><br>**N analyzed/enrolled** | PCC Outcome Description & Timing<br>**Main Results**<br><br>**Available Sub-group Data** | Reported findings related to secondary outcomes (bold indicates data used for synthesis where >1) |
|---|---|---|---|---|
| diagnosis of COVID-19 requiring hospitalization for more than 24 hours (from 11 hospitals in<br><br>Finland), restricted to those who completed the 1-year follow-up<br><br>ROB: Some concerns | N | Total: 181 analyzed/208 randomized | Subgroup: need of oxygen therapy during hospital admission | and attention/ concentration, single items)<br><br>Psychopathology (**EQ-5D-5L subscore, moderate to extreme anxiety/depression**; depression; mental anxiety)<br><br>HRQoL (EQ-VAS; range 0-100, higher score=higher quality of life)<br><br>Return to work (unable to return to work or study, single item) |
| Xu, 2023<br><br>China<br><br>Non-randomized clinical controlled trial<br><br>November 5 to 28, 2022<br><br>Adults aged between 18-50 years old, infected with the Omicron variant who | Hospitalized (% ICU NR)<br><br>Mean 29.3 (SD: 7.34); 40%; NR<br><br>Mild-to-moderate<br><br>N | IG: Paxlovid (oral nirmatrelvir plus ritonavir) twice daily for 5 days, ≤5 days of symptom onset (n=172 analyzed/195 completed the IG) (most not at high-risk)<br><br>CG: No exposure, but eligible. Received standard treatment (n=95 analyzed/120 enrolled)<br><br>Total: 267 analyzed/320 enrolled | Self-report PCC symptoms (WHO criteria), at/within 3 months post-discharge<br><br>IG: 32/172 (18.6%)<br><br>CG: 30/95 (31.6%)<br><br>P=0.016<br><br>aOR: 0.531 (0.290, 0.974), P=0.041 | Fatigue (single item)<br><br>Any adverse event (self-reported)<br><br>Serious adverse events (self-reported) |

| First Author, Year; Country; Study Design[ᵻ]; Timing of Infection<br><br>**Recruitment setting and participant eligibility**<br><br>**Risk of bias for primary outcome** | Type of Acute Care<br>Average age (years); % Female; % Vaccinated<br><br>**COVID-19 Illness Severity**<br><br>**Eligible Population All Having Post-acute Symptoms (Y/N)** | Intervention/Exposure Group (IG): Description & Timing<br><br>**Control Group (CG)**<br><br>**N analyzed/enrolled** | PCC Outcome Description & Timing<br>Main Results<br><br>**Available Sub-group Data** | Reported findings related to secondary outcomes (bold indicates data used for synthesis where >1) |
|---|---|---|---|---|
| were admitted to the designated hospital for treating COVID-<br><br>19 in Shijiazhuang<br><br>ROB: High | | | | |
| Wang 2023<br><br>China<br><br>Prospective comparative cohort using retrospective exposure data<br><br>April to June 2022 (discharge dates)<br><br>High-risk patients who were discharged from Shidong Hospital and who completed the follow-up<br><br>ROB:<br><br>IG1: Some concerns; IG2+IG3: High | Hospitalized (% ICU NR)<br><br>Mean 74.1 (SD: 11.4); 54.4%; NR<br><br>Moderate-to-critical<br><br>N | IG1: Paxlovid (nirmatrelvir plus ritonavir), timing NR (n=178)<br><br>IG2: Glucocorticoids (n=21)<br><br>IG3: Antibiotics (n=157)<br><br>CG: did not receive the IG in each comparison (n=456 for IG1, 613 for IG2, 477 for IG3)<br><br>Total: 634 analyzed/747 eligible for follow-up | Self-reported symptom checklist (13 items), 6 months after discharge<br><br>Crude proportions NR<br><br>IG1 vs. CG: aOR (age and COPD): 0.349 (0.205, 0.595), P<0.001<br><br>IG2 vs. CG: unadjusted OR: 1.377 (0.524,3.616), P=0.516<br><br>IG3 vs CG: unadjusted OR: 1.904 (1.271, 2.853), P=0.002<br><br>None | None |
| Frontera, 2021<br><br>US | Hospitalized (32.2% ICU admission)<br><br>Median 68.0 (IQR: 55-77) in neurological cases and | IG1: Corticosteroid (n=101)<br><br>IG2: Hydroxychloroquine (n=272) | Functional incapacity (i.e., Barthel index <100), median of 6.7 (IQR: 6.5-6.8) months post-neurological symptom onset | Fatigue (worse than average, neuro-QoL subscore, t-score >50) |

| First Author, Year; Country; Study Design[ǂ]; Timing of Infection<br><br>Recruitment setting and participant eligibility<br><br>Risk of bias for primary outcome | Type of Acute Care<br>Average age (years); % Female; % Vaccinated<br><br>COVID-19 Illness Severity<br><br>Eligible Population All Having Post-acute Symptoms (Y/N) | Intervention/Exposure Group (IG): Description & Timing<br><br>Control Group (CG)<br><br>N analyzed/enrolled | PCC Outcome Description & Timing<br>Main Results<br><br>Available Sub-group Data | Reported findings related to secondary outcomes (bold indicates data used for synthesis where >1) |
|---|---|---|---|---|
| Retrospective matched cohort<br><br>March 10 to May 20, 2020<br><br>Patients ≥18 years of age hospitalized at four New York hospitals and discharged alive; propensity-matched those with and without new neurological complications during index hospitalization<br><br>ROB: High | 69.0 (IQR: 57-78) in controls; 35%; NR<br><br>N | IG3: Azithromycin (n=246)<br><br>IG4: Therapeutic anticoagulation (n=134)<br><br>IG5: Zing (n=NR)<br><br>All during hospitalization<br><br>CG: did not receive the IG in each comparison (n=281 for IG1, 110 for IG2, 136 for IG3, 248 for IG4, NR for IG5)<br><br>Total: 382 (n=196 and 186 with and without neurological complications, respectively) analyzed/700 eligible for follow-up | Crude proportions: NR<br><br>IG1 vs. CG, unadjusted OR 0.48 (0.29, 0.80), P=0.005<br><br>Unadjusted P-values:<br><br>IG2 vs. CG=0.970<br><br>IG3 vs. CG=0.078<br><br>IG4 vs. CG=0.078<br><br>IG5 vs. CG=0.407<br><br>None | Cognitive impairment (moderate or worse, telephone MOCA score <18)<br><br>Psychopathology (worse than average anxiety and depression, neuro-Qol subscore, t-score >50)<br><br>Return to work (not returned to work of those working pre-morbidly, single item) |

| First Author, Year; Country; Study Design[ꟾ]; Timing of Infection<br><br>Recruitment setting and participant eligibility<br><br>Risk of bias for primary outcome | Type of Acute Care<br>Average age (years); % Female; % Vaccinated<br><br>COVID-19 Illness Severity<br><br>Eligible Population All Having Post-acute Symptoms (Y/N) | Intervention/Exposure Group (IG): Description & Timing<br><br>Control Group (CG)<br><br>N analyzed/enrolled | PCC Outcome Description & Timing<br>Main Results<br><br>Available Sub-group Data | Reported findings related to secondary outcomes (bold indicates data used for synthesis where >1) |
|---|---|---|---|---|
| Meije, 2021<br><br>Spain<br><br>Prospective cohort<br><br>March to May 2020 (discharge date)<br><br>Patients >15 years of age who were discharged after hospitalization for COVID-19<br><br>ROB: High | Hospitalized (8.9% ICU admission)<br><br>Mean 68.8 (SD: 12.7); 43% female<br><br>Variable severity (oxygen supplementation 74.8%; 1% IV) | IG: Outpatient assessment (standard follow-up protocol checklist of symptoms and adverse events, medical history, physical examination, laboratory testing including chest x-ray) with referrals as required (n=294)<br><br>CG: None<br><br>Total: 302/461 | Proportion with any persistent symptoms, median 7 (range 6-7.4) months after hospital discharge<br><br>At post discharge assessment vs. 7-month follow-up: 228/294 (77.6%) vs. 147/294 (50.0%) | Breathlessness/Dyspnea (dyspnea, single item)<br><br>Psychopathology (need for psychological medication, single item) |

Abbreviations: aOR: adjusted odds ratio; aHR: adjusted hazard ratio; CFQ: Cognitive Failures Questionnaire; CG: control group; EQ-5D: EuroQol-5 Dimensions-5 Levels; HRQoL: health-related quality of life; IG: intervention group; IQR: inter-quartile range; MCS: mental component score; mMRC: Modified Medical Research Council; NR: not reported; PCR- Polymerase Chain Reaction; SD: standard deviation; SF-36: 36-Item Short Form Health Survey questionnaire; VS: versus

[ꟾ]Study design: focus was on whether the exposure and baseline demographic information was prospectively or retrospectively collected.

Table S2. Quality assessment of the included cohort studies

| **Author, year**<br><br>**Outcome** | **1. Were the two groups similar and recruited from the same population?** | **2. Were the exposures measured similarly to assign people to both exposed and unexposed groups?** | **3. Was the exposure measured in a valid and reliable way?** | **4. Were confounding factors identified?** | **5. Were strategies to deal with confounding factors stated?** | **6. Were the groups/ participants free of the outcome at the start of the study (or at the moment of exposure)?** | **7. Were the outcomes measured in a valid and reliable way?** | **8. Was the follow up time reported and sufficient to be long enough for outcomes to occur?** | **9. Was follow up complete, and if not, were the reasons to loss to follow up described and explored?** | **10. Were strategies to address incomplete follow up utilized?** | **11. Was appropriate statistical analysis used ?** | **Covid test valid?** | **Evidence of selective reporting of results and/or analyses?** | **Evidence of missing outcome data?** | **Overall rating** |
|---|---|---|---|---|---|---|---|---|---|---|---|---|---|---|---|
| Rocha, 2023<br>PCC: Long COVID | + | + | - | + | - | + | + | + | ? | - | - | + | + | - | High |
| Rocha, 2023<br><br>Fatigue | + | + | ? | + | - | + | ? | + | ? | - | - | + | + | - | High |
| Rocha, 2023<br><br>Dyspnea | + | + | ? | + | - | + | ? | + | ? | - | - | + | + | - | High |
| Rocha, 2023<br><br>Memory loss | + | + | ? | + | - | + | ? | + | ? | - | - | + | + | - | High |
| Rocha, 2023<br>Attention loss | + | + | ? | + | - | + | ? | + | ? | - | - | + | + | - | High |
| Patel, 2024<br>PCC: PASC | ? | + | + | + | + | + | + | + | ? | - | ? | + | + | - | Some concerns |
| Patel, 2024<br>Anxiety/mood disorder | ? | + | + | + | + | + | ? | + | ? | - | ? | + | + | - | Some concerns |
| Durstenfeld, 2023 | + | + | - | + | + | + | + | + | - | ? | ? | + | - | - | High |

| | | | | | | | | | | | | | | | |
|---|---|---|---|---|---|---|---|---|---|---|---|---|---|---|---|
| PCC: Long COVID symptoms | | | | | | | | | | | | | | | |
| Cardona-Pascual, 2023<br><br>PCC: Persisting symptoms | + | + | + | + | + | + | ? | + | + | NA | + | + | ? | - | Some concerns |
| Cardona-Pascual, 2023<br><br>Breathlessness/<br><br>dyspnea | + | + | + | + | - | + | ? | + | + | NA | ? | + | ? | - | High |
| Cardona-Pascual, 2023<br><br>Depression/<br><br>anxiety | + | + | + | + | - | + | ? | + | + | NA | ? | + | ? | - | High |
| Cardona-Pascual, 2023<br><br>Cognitive impairment | + | + | + | + | - | + | ? | + | + | NA | ? | + | ? | - | High |
| Davelaar, 2023<br><br>PCC: Post-COVID syndrome | + | + | + | + | + | ? | ? | + | + | NA | + | + | - | - | Some concerns |
| Bertuccio, 2023<br><br>PCC: Long COVID symptoms | + | + | + | + | + | + | ? | + | - | - | ? | + | - | - | High |
| Congdon, 2023 | + | + | ? | + | + | + | + | + | ? | - | ? | + | - | - | Some concerns |

| | | | | | | | | | | | | | | | |
|---|---|---|---|---|---|---|---|---|---|---|---|---|---|---|---|
| PCC: Long COVID symptoms | | | | | | | | | | | | | | | |
| Congdon, 2023<br><br>Dyspnea | + | + | ? | + | - | + | ? | + | ? | - | - | + | - | - | High |
| Congdon, 2023<br><br>Generalized fatigue | + | + | ? | + | - | + | ? | + | ? | - | - | + | - | - | High |
| Congdon, 2023<br><br>Brain fog | + | + | ? | + | - | + | ? | + | ? | - | - | + | - | - | High |
| Chuang, 2023<br><br>PCC: Post-acute COVID-19 symptoms | + | + | + | + | + | + | + | + | + | NA | + | + | - | - | Low |
| Chuang, 2023<br><br>Hospitalization | + | + | + | + | + | + | + | + | + | NA | + | + | - | - | Low |
| Chuang, 2023<br><br>Fatigue | + | + | + | + | + | + | + | + | + | NA | + | + | - | - | Low |
| Chuang, 2023<br><br>Cognitive impairments | + | + | + | + | + | + | + | + | + | NA | + | + | - | - | Low |
| Leavy, 2023<br><br>PCC: Any symptoms | + | + | - | + | + | + | + | + | ? | - | ? | ? | - | - | High |
| Leavy, 2023<br><br>Recovery: Fully recovered from COVID-19 | + | + | - | + | + | + | ? | + | ? | - | ? | ? | - | - | High |

| | | | | | | | | | | | | | | |
|---|---|---|---|---|---|---|---|---|---|---|---|---|---|---|
| Leavy, 2023<br><br>HRQoL | + | + | - | + | + | + | ? | + | ? | - | ? | ? | - | - | High |
| Leavy, 2023<br><br>Fatigue | + | + | - | + | + | ? | + | + | ? | - | ? | ? | - | - | High |
| Leavy, 2023<br><br>Anxiety | + | + | - | + | + | ? | + | + | ? | - | ? | ? | - | - | High |
| Leavy, 2023<br><br>Cognitive impairment | + | + | - | + | + | ? | + | + | ? | - | ? | ? | - | - | High |
| Leavy, 2023<br><br>Lung function | + | + | - | + | + | ? | + | + | ? | - | ? | ? | - | - | High |
| Evans, 2021<br><br>Recovery: Patient-perceived recovery | + | + | + | + | + | + | ? | + | - | - | ? | + | - | - | High |
| Darcis, 2021<br><br>PCC: At least one symptom | ? | + | ? | + | - | + | ? | + | - | - | - | ? | - | - | High |
| Milne, 2021<br><br>PCC: Ongoing symptoms | + | + | + | + | ? | + | ? | + | + | NA | ? | + | - | - | Some concerns |
| Milne, 2021<br><br>Fatigue | + | + | + | + | ? | + | ? | + | + | NA | ? | + | - | - | Some concerns |
| Milne, 2021 | + | + | + | + | ? | ? | + | + | + | NA | ? | + | - | - | Some concerns |

| | | | | | | | | | | | | | | |
|---|---|---|---|---|---|---|---|---|---|---|---|---|---|---|---|
| Mental component score | | | | | | | | | | | | | | | |
| Berentschot, 2024<br><br>Recovery: Completely recovered | ? | + | + | + | - | + | + | + | ? | - | - | + | + | - | High |
| Berentschot, 2024<br><br>Dyspnea | ? | + | + | + | - | ? | + | + | ? | - | - | + | + | - | High |
| Berentschot, 2024<br><br>Fatigue | ? | + | + | + | - | ? | + | + | ? | - | - | + | + | - | High |
| Berentschot, 2024<br><br>Cognitive failures | ? | + | + | + | - | ? | + | + | ? | - | - | + | + | - | High |
| Berentschot, 2024<br><br>HRQoL | ? | + | + | + | - | ? | + | + | ? | - | - | + | + | - | High |
| Boglione, 2024<br><br>PCC: Long post-COVID syndrome | ? | + | ? | + | + | + | + | + | + | NA | + | ? | - | - | Some concerns |
| Ko, 2022<br><br>PCC: Persistent symptoms | + | + | + | + | + | + | ? | + | ? | - | ? | + | ? | - | Some concerns |
| Ko, 2022<br><br>Dyspnea | + | + | + | + | + | + | ? | + | ? | - | ? | + | ? | - | Some concerns |

| | | | | | | | | | | | | | | | |
|---|---|---|---|---|---|---|---|---|---|---|---|---|---|---|---|
| Wang, 2023<br><br>PCC: Long COVID (comparison 1) | ? | + | + | + | ? | + | + | + | + | NA | ? | ? | - | - | Some concerns |
| Wang, 2023<br><br>PCC: Long COVID (comparison 2) | ? | + | + | + | - | + | + | + | + | NA | ? | ? | - | - | High |
| Wang, 2023<br><br>PCC: Long COVID (comparison 3) | ? | + | + | + | - | + | + | + | + | NA | ? | ? | - | - | High |
| Frontera, 2021<br><br>PCC: Functional incapacity | ? | + | + | + | - | ? | + | + | ? | - | - | + | - | + | High |
| Frontera, 2021<br><br>Cognitive impairment | ? | + | + | + | - | ? | + | + | ? | - | - | + | - | + | High |
| Frontera, 2021<br><br>Anxiety | ? | + | + | + | - | ? | + | + | ? | - | - | + | - | + | High |
| Frontera, 2021<br><br>Depression | ? | + | + | + | - | ? | + | + | ? | - | - | + | - | + | High |
| Frontera, 2021<br><br>Fatigue | ? | + | + | + | - | ? | + | + | ? | - | - | + | - | + | High |
| Frontera, 2021<br><br>Return to work | ? | + | + | + | - | + | ? | + | ? | - | - | + | - | + | High |
| Meije, 2021 | + | + | + | + | - | + | ? | + | - | + | - | + | - | - | High |

| | | | | | | | | | | | | | | | |
|---|---|---|---|---|---|---|---|---|---|---|---|---|---|---|---|
| PCC: Persistent symptoms | | | | | | | | | | | | | | | |
| Meije, 2021<br><br>Need for psychological medication | + | + | + | + | - | + | - | + | - | + | - | + | - | - | High |
| Meije, 2021<br><br>Dyspnea | + | + | + | + | - | + | - | + | - | + | - | + | - | - | High |

+: Yes/low risk of bias; -: No/high risk of bias; ?: Unclear/Some concerns of bias; NA: Not applicable

Table S3: Quality assessment of the included randomized trials

| **Author, year** | **Domain 1. Randomization process** | **Domain 2. Deviations from the intended interventions (effect of assignment to intervention)** | **Domain 3. Missing outcome data** | **Domain 4. Measurement of the outcome** | **Domain 5. Selection of reported results** | **COVID-19 test valid?** | **Overall rating** |
|---|---|---|---|---|---|---|---|
| Greenstein, 2024<br><br>PCC: Presence of any symptoms | + | ? | - | ? | + | + | High |
| Greenstein, 2024<br><br>HRQoL | + | ? | - | ? | + | + | High |
| Greenstein, 2024<br><br>Anxiety/depression | + | ? | - | ? | + | + | High |
| Greenstein, 2024<br><br>Shortness of breath with extertion | + | ? | - | ? | + | + | High |

| | | | | | | | |
|---|---|---|---|---|---|---|---|
| Greenstein, 2024<br><br>Shortness of breath at rest | + | ? | - | ? | + | + | High |
| Greenstein, 2024<br><br>Cognitive impairment | + | ? | - | ? | + | + | High |
| Greenstein, 2024<br><br>Fatigue | + | ? | - | ? | + | + | High |
| Kolesnyk, 2024<br><br>PCC: Post-COVID functional scale | + | + | + | + | + | + | Low |
| Kolesnyk, 2024<br><br>Attention deficit | + | + | + | + | + | + | Low |
| Kolesnyk, 2024<br><br>Fatigue | + | + | + | + | + | + | Low |
| Kolesnyk, 2024<br><br>Depression | + | + | + | + | + | + | Low |
| Kolesnyk, 2024<br><br>Anxiety | + | + | + | + | + | + | Low |
| Kolesnyk, 2024<br><br>Withdrawals due to AEs | + | + | + | + | + | + | Low |
| Gebo, 2023<br><br>PCC: Any new symptom | + | + | + | + | + | ? | Low |
| Gebo, 2023<br><br>Subgroup by timing of drug administration, PCC: Any new symptom | ? | + | + | + | + | ? | Some concerns |

| | | | | | | | |
|---|---|---|---|---|---|---|---|
| Xu, 2023<br><br>PCC: Post COVID condition | - | ? | ? | ? | + | + | High |
| Xu, 2023<br><br>Fatigue | - | ? | ? | ? | + | + | High |
| Xu, 2023<br><br>Adverse events | - | ? | + | ? | + | + | High |
| Bramante, 2023<br><br>PCC: Long COVID | + | + | ? | + | + | + | Some concerns |
| Bramante, 2023<br><br>Serious AEs | + | + | + | ? | + | + | Low |
| Nevalainen, 2022<br><br>Recovery | + | ? | + | ? | + | + | Some concerns |
| Nevalainen, 2022<br><br>HRQoL | + | ? | + | ? | + | + | Some concerns |
| Nevalainen, 2022<br><br>Fatigue | + | ? | + | ? | + | + | Some concerns |
| Nevalainen, 2022<br><br>Anxiety/depression | + | ? | + | ? | + | + | Some concerns |
| Nevalainen, 2022<br><br>Dyspnea | + | ? | + | ? | + | + | Some concerns |
| Nevalainen, 2022<br><br>Problems with attention and concentration | + | ? | + | ? | + | + | Some concerns |

| | | | | | | | |
|---|---|---|---|---|---|---|---|
| Nevalainen, 2022<br><br>Memory difficulties and forgetting things | + | ? | + | ? | + | + | Some concerns |
| Nevalainen, 2022<br><br>Post-exertional malaise | + | ? | + | ? | + | + | Some concerns |
| Nevalainen, 2022<br><br>Inability to return to work | + | ? | + | ? | + | + | Some concerns |

+: Low risk of bias; -: High risk of bias; ?: Unclear/Some concerns of bias; NA: Not applicable

Table S4. Summary of findings for secondary outcomes, by intervention

| **Study design, acute care setting**<br><br>**(Number of studies;**<br><br>**Sample size)** | **Pooled estimate (95% CI); $I^2$** | **GRADE rating** [ǂ] | **What happens?** |
|---|---|---|---|
| **Fatigue: Antivirals** | | | |
| Trials, inpatients (1 study; N=181) | Remdesivir: **Inpatients, RR (95% CI): 0.88 (95% CI: 0.54, 1.44)** | Very low[d, G] | Among individuals who received inpatient COVID-19 care, we are **very uncertain** about the effects of remdesivir on the risk of moderate to severe fatigue. |

| | | | |
|---|---|---|---|
| Observational, inpatients (1 study; N=267)<br><br>Observational, inpatients, continuous (1 study [Berentschot 2024], n=197) | **Inpatients, OR (95% CI): 0.57 (0.25-1.29)**<br><br>**Inpatients, MD (95% CI): 1.50 (-1.08-4.08)** | Very low[G] | Among individuals who received inpatient COVID-19 care, we are **very uncertain** about the effects of antivirals on the risk of fatigue. |
| Observational, outpatients (2 studies; N=24,990) | Paxlovid (nirmatrelvir and ritonavir): **Outpatient pooled estimate RR (95% CI): 0.91 (0.79-1.05); $I^2$ = 0%** | Low | Among individuals who received outpatient COVID-19 care, paxlovid (nirmatrelvir and ritonavir) **may not reduce** the risk of fatigue. |
| **Fatigue: Hydroxychloroquine** | | | |
| Observational, inpatients (1 study; N=382) | **Inpatients, p=0.221** | Very low[a] | Among individuals who received inpatient COVID-19 care, we are **very uncertain** about the effects of hydroxychloroquine on the risk of worse than average fatigue. |
| **Fatigue: Steroids** | | | |
| Observational, inpatients (2 studies; N=469)<br><br>Observational, inpatients continuous (2 studies [Berentschot 2024; Leavy 2023]; N=1,594) | **Inpatient pooled estimate OR (95% CI): 1.05 (0.23-4.85); $I^2$ = 89%**<br><br>**Inpatient pooled estimate SMD (95% CI): 0.02 (-0.09-0.12); $I^2$ = 0%** | Very low[a, G] | Among individuals who received inpatient COVID-19 care, we are **very uncertain** about the effects of steroids on the risk of fatigue. |
| **Fatigue: Anti-inflammatory treatment** | | | |
| Observational, inpatients (1 study [Berentschot 2024]; N=176) | **Inpatients, MD (95% CI): 0.10 (-2.77-2.97)** | Very low[a, d] | Among individuals who received inpatient COVID-19 care, we are **very uncertain** about the effects of anti-inflammatory treatment on the risk of fatigue. |
| **Fatigue: Azithromycin** | | | |
| Observational, inpatients (1 study; N=382) | **Inpatients, p=0.633** | Very low[a] | Among individuals who received inpatient COVID-19 care, we are **very uncertain** about the effects of azithromycin on the risk of worse than average fatigue. |
| **Fatigue: Therapeutic anticoagulants** | | | |

| | | | |
|---|---|---|---|
| Trials, inpatients (1 study; N=571) | Therapeutic-dose heparin (vs. prophylactic-dose heparin): **Inpatients, RR (95% CI): 1.05 (0.85-1.30)** | Low[a, g] | Among individuals who received inpatient COVID-19 care, therapeutic-heparin versus prophylactic-dose heparin **may not reduce** the risk of fatigue. |
| Observational, inpatients (1 study; N=382) | **Inpatients, OR (95% CI): 1.80 (1.10-3.10); p=0.022** | Very low[a, g] | Among individuals who received inpatient COVID-19 care, we are **very uncertain** about the effects of therapeutic anticoagulants on the risk of worse than average fatigue. |
| **Fatigue: Probiotics** | | | |
| Trials, outpatients (1 study; N=69) | **Inpatients, RR (95% CI): 1.34 (0.68-2.63)** | Very low[e, G] | Among individuals who received outpatient COVID-19 care, we are **very uncertain** about the effects of probiotics on the risk of fatigue. |
| **Fatigue: Zinc** | | | |
| Observational, inpatients (1 study; N=NR) | **Inpatients, p=0.272** | Very low[A] | Among individuals who received inpatient COVID-19 care, we are **very uncertain** about the effects of zinc on the risk of worse than average fatigue. |
| **Fatigue: Activity** | | | |
| Observational, outpatients (1 study; N=823) | Remained active (vs. became inactive): **Outpatients, RR (95% CI): 0.36 (0.25-0.52)** | Very low[a] | Among individuals who received outpatient COVID-19 care, we are **very uncertain** about the effects of activity level on the risk of fatigue. |
| **Breathlessness/Dyspnea: Antivirals** | | | |
| Trials, inpatients (1 study; N=181) | Remdesivir: **Inpatients, RR (95% CI): 0.61 (0.20, 1.85)** | Very low[d, G] | Among individuals who received inpatient COVID-19 care, we are **very uncertain** about the effects of remdesivir on the risk of exertional dyspnea. |
| Observational, inpatients (1 study; N=186) | **Inpatients, RR (95% CI): 1.16 (0.84-1.60)** | Very low[a, d, g] | Among individuals who received inpatient COVID-19 care, we are **very uncertain** about the effects of antivirals on the risk of breathlessness/dyspnea. |
| Observational, outpatients (1 study; N=500) | Paxlovid (nirmatrelvir and ritonavir): **Outpatients, RR (95% CI): 0.68 (0.42-1.09)** | Very low[a, g] | Among individuals who received outpatient COVID-19 care, we are **very uncertain** about the effects of paxlovid (nirmatrelvir and ritonavir) on the risk of breathlessness/dyspnea. |
| **Breathlessness/Dyspnea: Steroids** | | | |
| Observational, inpatients (3 studies; N=1,865) | **Inpatient pooled estimate OR (95% CI): 0.79 (0.59-1.05); $I^2$ = 0%** | Very low[a, d] | Among individuals who received inpatient COVID-19 care, we are **very uncertain** about the effects of steroids on the risk of breathlessness/dyspnea. |

| | | | |
|---|---|---|---|
| | Sensitivity analysis (excluding high ROB studies [2 studies; N=1,559], both also >6 months): 0.82 (0.48-1.40); I² = NA | | |
| **Breathlessness/Dyspnea: Anti-inflammatory treatment** | | | |
| Observational, inpatients (1 study; N=159) | **Inpatients, RR (95% CI): 0.80 (0.50-1.27)** | Very low[a, d, g] | Among individuals who received inpatient COVID-19 care, we are **very uncertain** about the effects of anti-inflammatory treatment on the risk of breathlessness/dyspnea. |
| **Breathlessness/Dyspnea: Therapeutic-dose heparin** | | | |
| Trials, inpatients (1 study; N=571) | Therapeutic-dose heparin (vs. prophylactic-dose heparin): **Inpatients, average RR (95% CI): 0.96 (0.56-1.36)** | Very low[a, G] | Among individuals who received inpatient COVID-19 care, we are **very uncertain** about the effects of therapeutic-dose heparin versus prophylactic-dose heparin on the risk of breathlessness/dyspnea. |
| **Breathlessness/Dyspnea: Tocilizumab** | | | |
| Observational, inpatients (1 study; N=391) | **Inpatients, RR (95% CI): 2.03 (1.37-3.00)** | Very low[a] | Among individuals who received inpatient COVID-19 care, we are **very uncertain** about the effects of tocilizumab on the risk of breathlessness/dyspnea. |
| **Breathlessness/Dyspnea: Activity** | | | |
| Observational, outpatients (1 study; N=823) | Remained active (vs. became inactive): **Outpatients, RR (95% CI): 0.34 (0.18-0.65)** | Very low[a] | Among individuals who received outpatient COVID-19 care, we are **very uncertain** about the effects of activity level on the risk of breathlessness/dyspnea. |
| **Breathlessness/Dyspnea: Outpatient assessment and referrals** | | | |
| Observational, inpatients (1 study; N=294) | **Post-discharge outpatient care: 28 (9.5%)** | Very low[A] | Among individuals who received inpatient COVID-19 care, we are **very uncertain** about the effects of outpatient assessment and referrals on the risk of breathlessness/dyspnea. |
| **Cognitive impairment: Antivirals** | | | |
| Trials, inpatients (1 study; N=181) | Remdesivir: **Inpatients, average RR (95% CI): 1.15 (0.40-1.90)** | Very low[d, G] | Among individuals who received inpatient COVID-19 care, we are **very uncertain** about the effects of remdesivir on the risk of moderate to severe cognitive impairment. |
| Observational, inpatients (1 study [Berentschot 2024]; n=197) | **Inpatients, MD (95% CI): -0.40 (-5.56-4.76)** | Very low[a, d, g] | Among individuals who received inpatient COVID-19 care, we are **very uncertain** about the effects of antivirals on the risk of cognitive impairment. |

| | | | |
|---|---|---|---|
| Observational, outpatients (2 studies; N=24,990) | Paxlovid (nirmatrelvir and ritonavir): **Outpatients, pooled estimate RR (95% CI): 0.72 (0.59-0.88); $I^2 = 0\%$** | Very low[g] | Among individuals who received outpatient COVID-19 care, we are **very uncertain** about the effects of paxlovid (nirmatrelvir and ritonavir) on the risk of cognitive impairment. |
| **Cognitive impairment: Hydroxychloroquine** | | | |
| Observational, inpatients (1 study; N=382) | **Inpatients, p=0.128** | Very low[a] | Among individuals who received inpatient COVID-19 care, we are **very uncertain** about the effects of hydroxychloroquine on the risk of cognitive impairment. |
| **Cognitive impairment: Steroids** | | | |
| Observational, inpatients (1 study; N=965)<br><br>Observational, inpatients p-value (1 study, N=382)<br><br>Observational, inpatients continuous (1 study [Berentschot 2024]; N=382); continuous | **Inpatients, RR (95% CI): 0.84 (0.57-1.23)**<br><br>**Inpatients, p=0.982**<br><br>**Inpatients, MD (95% CI): -1.60 (-5.68-2.48)** | Very low[a, d, g] | Among individuals who received inpatient COVID-19 care, we are **very uncertain** about the effects of steroids on the risk of cognitive impairment. |
| **Cognitive impairment: Anti-inflammatory treatment** | | | |
| Observational, inpatients (1 study [Berentschot 2024]; N=176) | **Inpatients, MD (95% CI): -2.50 (-6.28-1.28)** | Very low[a, d, g] | Among individuals who received inpatient COVID-19 care, we are **very uncertain** about the effects of anti-inflammatory treatment on the risk of cognitive impairment. |
| **Cognitive impairment: Tocilizumab** | | | |
| Observational, inpatients (1 study; N=391) | **IG: NR vs. CG: 1 (0.4%)** | Very low[a] | Among individuals who received inpatient COVID-19 care, we are **very uncertain** about the effects of tocilizumab on the risk of cognitive impairment. |
| **Cognitive impairment: Azithromycin** | | | |
| Observational, inpatients (1 study; N=382) | **Inpatients, p=0.188** | Very low[a] | Among individuals who received inpatient COVID-19 care, we are **very uncertain** about the effects of azithromycin on the risk of cognitive impairment. |
| **Cognitive impairment: Therapeutic anticoagulants** | | | |

| | | | |
|---|---|---|---|
| Trials, inpatients (1 study; N=571) | Therapeutic-dose heparin (vs. prophylactic-dose heparin): **Inpatients, RR (95% CI): 0.49 (0.22-1.07)** | Low[a, g]<br><br>(small-moderate difference)<br><br>Very low[a, G]<br><br>(large difference) | Among individuals who received outpatient COVID-19 care, therapeutic-dose heparin versus prophylactic-dose heparin **may reduce** the risk of cognitive impairment.<br><br>Among individuals who received inpatient COVID-19 care, we are **very uncertain** about the effects of therapeutic-dose heparin versus prophylactic-dose heparin on the risk of cognitive impairment. |
| Observational, inpatient (1 study; N=382) | **Inpatients, p=0.995** | Very low[a] | Among individuals who received inpatient COVID-19 care, we are **very uncertain** about the effects of therapeutic anticoagulants on the risk of cognitive impairment. |
| **Cognitive impairment: Zinc** | | | |
| Observational, inpatients (1 study; N=NR) | **Inpatients, p=0.523** | Very low[A] | Among individuals who received inpatient COVID-19 care, we are **very uncertain** about the effects of zinc on the risk of cognitive impairment. |
| **Cognitive impairment: Probiotics** | | | |
| Trials, inpatients (1 study; N=69) | **Inpatients, RR (95% CI): 1.44 (0.51-4.10)** | Very low[e, G] | Among individuals who received inpatient COVID-19 care, we are **very uncertain** about the effects of probiotics on the risk of cognitive impairment. |
| **Cognitive impairment: Activity** | | | |
| Observational, outpatients (1 study; N=823) | Remained active (vs. became inactive): **Outpatients, average RR (95% CI): 0.63 (0.40-0.86)** | Very low[a, g] | Among individuals who received inpatient COVID-19 care, we are **very uncertain** about the effects of activity level on the risk of cognitive impairment. |
| **Psychopathology: Antivirals** | | | |
| Trials, inpatient (1 study; N=181) | Remdesivir: **Inpatients, RR (95% CI): 1.27 (0.47-3.42)** | Very low[d, G] | Among individuals who received inpatient COVID-19 care, we are **very uncertain** about the effects of remdesivir on the risk of moderate to severe anxiety. |
| Observational, outpatient (1 study; N=53,186) | Paxlovid (nirmatrelvir and ritonavir): **Outpatients, RR (95% CI): 0.71 (0.69-0.74)** | Low | Among individuals with risk factors for severe COVID-19 who received outpatient COVID-19 care, paxlovid (nirmatrelvir and ritonavir) **may reduce** the risk of psychopathology. |
| **Psychopathology: Hydroxychloroquine** | | | |
| Observational, inpatients (1 study; N=382) | Anxiety: **Inpatients: p=0.945**<br><br>Depression: **Inpatients: p=0.500** | Very low[a] | Among individuals who received inpatient COVID-19 care, we are **very uncertain** about the effects of hydroxychloroquine on the risk of psychopathology. |

| | | | |
|---|---|---|---|
| **Psychopathology: Corticosteroids** | | | |
| Observational, inpatients (1 study; N=1,227)<br><br>Observational, inpatients p-values (1 study; N=382)<br><br>Observational, inpatients, continuous (1 study; N=87) | Anxiety: **Inpatients, RR (95% CI): 0.96 (0.77-1.18)**<br><br>Anxiety: **Inpatients: p=0.822**<br><br>Depression: **Inpatients: p=0.367**<br><br>Mental disability:<br><br>**MD (95% CI): -4.00 (-8.74-0.74)** | Very low[a] | Among individuals who received inpatient COVID-19 care, we are **very uncertain** about the effects of corticosteroids on the risk of psychopathology. |
| **Psychopathology: Tocilizumab** | | | |
| Observational, inpatients (1 study; N=391) | **Inpatients, RR (95% CI): 0.90 (0.28-2.88)** | Very low[a, G] | Among individuals who received inpatient COVID-19 care, we are **very uncertain** about the effects of tocilizumab on the risk of psychopathology. |
| **Psychopathology: Azithromycin** | | | |
| Observational, inpatients (1 study; N=382) | Anxiety: **Inpatients: p=0.606**<br><br>Depression: **Inpatients: p=0.104** | Very low[a] | Among individuals who received inpatient COVID-19 care, we are **very uncertain** about the effects of azithromycin on the risk of worse than average psychopathology. |
| **Psychopathology: Therapeutic anticoagulants** | | | |
| Trials, inpatients (1 study; N=410) | Therapeutic-dose heparin (vs. prophylactic-dose heparin): **Inpatients, RR (95% CI): 1.06 (0.67-1.69)** | Very low[a, G] | Among individuals who received inpatient COVID-19 care, we are **very uncertain** about the effects of therapeutic-dose heparin versus prophylactic-dose heparin on the risk of moderate to extreme psychopathology. |
| Observational, inpatients (1 study; N=382) | Anxiety: **Inpatients: p=0.914**<br><br>Depression: **Inpatients: p=0.982** | Very low[a] | Among individuals who received inpatient COVID-19 care, we are **very uncertain** about the effects of therapeutic anticoagulants on the risk of worse than average psychopathology. |
| **Psychopathology: Zinc** | | | |
| Observational, inpatients (1 study; N=NR) | Anxiety: **Inpatients: p=0.906**<br><br>Depression: **Inpatients: p=0.621** | Very low[A] | Among individuals who received inpatient COVID-19 care, we are **very uncertain** about the effects of zinc on the risk of worse than average psychopathology. |
| **Psychopathology: Probiotics** | | | |

| | | | |
|---|---|---|---|
| Trials, outpatients (1 study; N=69) | **Inpatients, average RR (95% CI): 0.93 (0.00-2.98)** | Very low[e, G] | Among individuals who received outpatient COVID-19 care, we are **very uncertain** about the effects of probiotics on the risk of psychopathology. |
| **Psychopathology: Outpatient assessment and referrals** | | | |
| Observational, inpatients (1 study; N=294) | **Post-discharge outpatient care: 30 (10.2%)** | Very low[A] | Among individuals who received inpatient COVID-19 care, we are **very uncertain** about the effects of outpatient assessment and referrals on the risk of psychopathology (need for psychological medication). |
| **Hospitalization: Paxlovid (Nirmatrelvir and ritonavir)** | | | |
| Observational, outpatients (1 study; N=24,490) | **Outpatients, RR (95% CI): 0.54 (0.45-0.65)** | Low | Among individuals who received outpatient COVID-19 care, paxlovid (nirmatrelvir and ritonavir) **may reduce** the risk of psychopathology. |
| **Health-related quality of life: Antivirals** | | | |
| Trials, inpatients (1 study; N=181) | Remdesivir: **Inpatients, OR (95% CI): 0.83 (0.49-1.40)** | Low[d, g] | Among individuals who received inpatient COVID-19 care, remdesivir **may not reduce** the risk of worse health-related quality of life. |
| Observational, inpatients (1 study [Berentschot 2024]; N=197) | **Inpatients, MD (95% CI): 0.01 (-0.06-0.08)** | Very low[a, d] | Among individuals who received inpatient COVID-19 care, we are **very uncertain** about the effects of antivirals on the risk of worse health-related quality of life. |
| **Health-related quality of life: Steroids** | | | |
| Observational, inpatients (2 studies [Berentschot 2024; Leavy 2023]; N=1,594) | **Inpatient pooled estimate MD (95% CI): -0.00 (-0.03-0.02); I² = 13%** | Very low[a, d] | Among individuals who received inpatient COVID-19 care, we are **very uncertain** about the effects of steroids on the risk of worse health-related quality of life. |
| **Health-related quality of life: Anti-inflammatory treatments** | | | |
| Observational, inpatients (1 study; N=176) | **Inpatients, MD (95% CI): 0.02 (-0.04-0.08)** | Very low[a, d] | Among individuals who received inpatient COVID-19 care, we are **very uncertain** about the effects of anti-inflammatory treatments on the risk of worse health-related quality of life. |
| **Health-related quality of life: Therapeutic-dose heparin** | | | |
| Trials, inpatients (1 study [Greenstein 2024]; N=410) | Therapeutic-dose heparin (vs. prophylactic-dose heparin): **Inpatients, MD (95% CI): 0.12 (-3.22-3.46)** | Moderate[a] | Among individuals who received inpatient COVID-19 care, therapeutic-dose heparin versus prophylactic-dose heparin **probably does not reduce** the risk of worse health-related quality of life. |
| **Post-exertional malaise: Remdesivir** | | | |

| | | | |
|---|---|---|---|
| Trials, inpatients (1 study; N=181) | **Inpatients, RR (95% CI): 0.41 (0.15-1.12)** | Low[d, g]<br><br>(small-moderate difference)<br><br>Very low[d, G]<br><br>(large difference) | Among individuals who received inpatient COVID-19 care, remdesivir **may reduce** the risk of moderate to severe prolonged general malaise following even light exertion.<br><br>Among individuals who received inpatient COVID-19 care, we are **very uncertain** about the effects of remdesivir on the risk of the risk of moderate to severe prolonged general malaise following even light exertion. |
| **Not returned to work or study: Remdesivir** | | | |
| Trials, inpatients (1 study; N=111) | **Inpatients, RR (95% CI): 0.26 (0.01-6.13)** | Very low[d, G] | Among individuals who received inpatient COVID-19 care, we are **very uncertain** about the effects of remdesivir on the risk of not returning to work or study. |
| **Not returned to work: Hydroxychloroquine** | | | |
| Observational, inpatients (1 study; N=NR) | Return to work: **Inpatients: p=0.192** | Very low[A] | Among individuals who received inpatient COVID-19 care, we are **very uncertain** about the effects of hydroxychloroquine on the risk of not returning to work. |
| **Not returned to work: Corticosteroids** | | | |
| Observational, inpatients (1 study; N=NR) | **Inpatients: OR (95% CI): 2.50 (1.25-5.00)** | Very low[a, G] | Among individuals who received inpatient COVID-19 care, we are **very uncertain** about the effects of corticosteroids on the risk of not returning to work. |
| **Not returned to work: Azithromycin** | | | |
| Observational, inpatients (1 study; N=NR) | Return to work: **Inpatients: p=0.458** | Very low[A] | Among individuals who received inpatient COVID-19 care, we are **very uncertain** about the effects of azithromycin on the risk of not returning to work. |
| **Not returned to work: Therapeutic anticoagulants** | | | |
| Observational, inpatients (1 study; N=NR) | **Inpatients, OR (95% CI): 3.23 (1.67-6.25)** | Very low[a] | Among individuals who received inpatient COVID-19 care, we are **very uncertain** about the effects of therapeutic anticoagulants on the risk of not returning to work. |
| **Not returned to work: Zinc** | | | |
| Observational, inpatients (1 study; N=NR) | **Inpatients, OR (95% CI): 0.43 (0.23-0.83)** | Very low[a, g] | Among individuals who received inpatient COVID-19 care, we are **very uncertain** about the effects of zinc on the risk of not returning to work. |
| **Any adverse events: Paxlovid (Nirmatrelvir and Ritonavir)** | | | |

| | | | |
|---|---|---|---|
| Observational, inpatients (1 study; N=267) | **Inpatients, RR (95% CI): 1.33 (0.48-3.65)** | Very low[a, G] | Among individuals who received inpatient COVID-19 care, we are **very uncertain** about the effects of paxlovid (nirmatrelvir plus Ritonavir) on the risk of any adverse events. |
| **Any adverse events: Probiotics** | | | |
| Trials, outpatients (1 study; N=71) | **Outpatients, RR (95% CI): 3.08 (0.13-73.23)** | Very low[e, G] | Among individuals who received outpatient COVID-19 care, we are **very uncertain** about the effects of probiotics on the risk of any adverse events. |
| **Serious adverse events: Paxlovid (Nirmatrelvir and Ritonavir)** | | | |
| Observational, inpatients (1 study; N=267) | **Inpatients: zero events** | Very low[a, G, h] | Among individuals who received inpatient COVID-19 care, we are **very uncertain** about the effects of paxlovid (nirmatrelvir plus ritonavir) on the risk of serious adverse events. |
| **Serious adverse events: Metformin** | | | |
| Trials, outpatients (1 study; N=569) | **Inpatients: zero events** | Low[G, h] | Among individuals with a BMI of ≥25 kg/m$^2$ who received outpatient COVID-19 care, metformin **may not reduce** the risk of serious adverse events. |
| **Serious adverse events: Fluvoxamine** | | | |
| Trials, outpatients (1 study; N=319) | **Inpatients: zero events** | Low[G, h] | Among individuals with a BMI of ≥25 kg/m$^2$ who received outpatient COVID-19 care, fluvoxamine **may not reduce** the risk of serious adverse events. |
| **Serious adverse events: Ivermectin** | | | |
| Trials, outpatients (1 study; N=406) | **Inpatients: zero events** | Low[G, h] | Among individuals with a BMI of ≥25 kg/m$^2$ who received outpatient COVID-19 care, ivermectin **may not reduce** the risk of serious adverse events. |
| **Serious adverse events: Metformin and Ivermectin** | | | |
| Trials, outpatients (1 study; N=401) | **Inpatients: zero events** | Low[G, h] | Among individuals with a BMI of ≥25 kg/m$^2$ who received outpatient COVID-19 care, metformin and ivermectin **may not reduce** the risk of serious adverse events. |
| **Serious adverse events: Metformin and Fluvoxamine** | | | |
| Trials, outpatients (1 study; N=336) | **Inpatients: zero events** | Low[G, h] | Among individuals with a BMI of ≥25 kg/m$^2$ who received outpatient COVID-19 care, metformin and fluvoxamine **may not reduce** the risk of serious adverse events. |

[i] Thresholds used in GRADE ratings were: RR 0.75-1.25 for little-to-no difference; RR 0.51-0.74 and 1.26-1.99 for a small-to-moderate difference, and ≤0.50 or ≥2.00 for a large difference.

Reasons (note use of capital letters when rating down twice under one domain for very serious concerns):

**a**=serious concerns of risk of bias; **b**=serious concerns of inconsistency; **c**=serious concerns of indirectness due to use of surrogate outcome; **d**=serious concerns of indirectness due to timing of outcome ascertainment (outcome ascertainment outside of 3 to 12-month follow-up); **e**=serious concerns of population (generalizability); **f**=serious concerns of outcome definition; **g**=serious concerns of imprecision. **h**=Some concerns about indirectness as serious adverse events were not defined (unclear whether requiring hospitalization), and serious concerns of imprecision due to zero events and sample size <2000.

Table S5. Subgroup data for post-COVID-19 condition

| Author, Year | Intervention (s) | Subgroups/sensitivity analyses |
|---|---|---|
| Bertuccio, 2023 | 1. Mononclonal antibodies<br><br>2. Antivirals | Sensitivity analysis by time period (most prevalent COVID variants):<br><br>Sensitivity analysis did not show any differences between the three periods, where prevalent variants were respectively Alpha, Delta and Omicron. |
| Chuang, 2023 (Patel 2024 used for primary outcome from TriNetX database) | Antivirals | Subgroup, Age groups, OR (95% CI):<br><br>≥65 years: 1.073 (0.967, 1.190)<br><br>45–64 years: 0.999 (0.898, 1.111)<br><br>18–44 years: 1.033 (0.904, 1.181)<br><br>p-value: >0.05<br><br>Subgroup, Sex, OR (95% CI):<br><br>Male: 1.042 (0.939, 1.155)<br><br>Female: 1.004 (0.924, 1.092)<br><br>p-value: >0.05<br><br>Subgroup, Vaccination status, OR (95% CI):<br><br>Unvaccinated: 0.945 (0.869, 1.031)<br><br>Vaccinated: 1.053 (0.951, 1.164)<br><br>p-value: >0.05 |
| Gebo, 2023 | Convalescent plasma | Subgroup, timing of intervention, RR (95% CI):<br><br>Convalescent plasma ≤5 days (vs. control plasma ≤5 days): Outpatients, RR: 0.84 (0.62-1.13)<br><br>Convalescent plasma >5 days (vs. control plasma >5 days): Outpatients, RR: 1.00 (0.79-1.27) |

| | | |
|---|---|---|
| Bramante, 2023 | 1. Metformin<br><br>2. Fluvoxamine<br><br>3. Ivermectin | Subgroup, sex, HR (95% CI):<br><br>Female: Metformin vs. placebo: 0.56 (0.34-0.91)<br><br>Male: Metformin vs. placebo: 0.76 (0.34-1.69)<br><br>p-value: 0.53<br><br>Female: Fluvoxamine vs. placebo: 1.40 (0.74-2.65)<br><br>Male: Fluvoxamine vs. placebo: 1.55 (0.52-4.62)<br><br>p-value: 0.88<br><br>Female: Ivermectin vs. placebo: 1.04 (0.58-1.88)<br><br>Male: Ivermectin vs. placebo: 0.86 (0.30-2.44)<br><br>p-value: 0.75<br><br>Subgroup, BMI, HR (95% CI):<br><br><30 kg/m$^2$: Metformin vs. placebo: 0.81 (0.44-1.47)<br><br>≥30 kg/m$^2$: Metformin vs. placebo: 0.44 (0.24-0.80)<br><br>p-value: 0.15<br><br><30 kg/m$^2$: Fluvoxamine vs. placebo: 1.51 (0.65-3.48)<br><br>≥30 kg/m$^2$: Fluvoxamine vs. placebo: 1.26 (0.61-2.62)<br><br>p-value: 0.75<br><br><30 kg/m$^2$: Ivermectin vs. placebo: 1.08 (0.51-2.26)<br><br>≥30 kg/m$^2$: Ivermectin vs. placebo: 0.85 (0.41-1.73)<br><br>p-value: 0.64<br><br>Subgroup, Time from symptom onset to first dose of study drug (days), HR (95% CI):<br><br>≤3 days: Metformin vs. placebo: 0.37 (0.15-0.95)<br><br>≥4 days: Metformin vs. placebo: 0.66 (0.41-1.06)<br><br>p-value: 0.27<br><br>≤3 days: Fluvoxamine vs. placebo: 1.47 (0.42-5.23) |

| | | |
|---|---|---|
| | | ≥4 days: Fluvoxamine vs. placebo: 1.32 (0.72-2.43)<br>p-value: 0.88<br><br>≤3 days: Ivermectin vs. placebo: 0.80 (0.30-2.12)<br>≥4 days: Ivermectin vs. placebo: 1.00 (0.54-1.82)<br>p-value: 0.70<br><br>Subgroup, Age groups:<br><45 years: Metformin vs. placebo: 0.39 (0.20-0.73)<br>≥45 years: Metformin vs. placebo: 0.85 (0.48-1.51)<br>p-value: 0.07<br><br><45 years: Fluvoxamine vs. placebo: 0.87 (0.37-2.07)<br>≥45 years: Fluvoxamine vs. placebo: 1.78 (0.84-3.77)<br>p-value: 0.22<br><br><45 years: Ivermectin vs. placebo: 1.00 (0.51-1.98)<br>≥45 years: Ivermectin vs. placebo: 0.93 (0.42-2.04)<br>p-value: 0.89<br><br>Subgroup, Dominant SARS-CoV-2 variant at time of randomization, HR (95% CI):<br>Alpha: Metformin vs. placebo: 0.21 (0.02-1.87)<br>Delta: Metformin vs. placebo: 0.68 (0.42-1.12)<br>Omicron: Metformin vs. placebo: 0.45 (0.18-1.11)<br>p-value: 0.41<br><br>Alpha: Fluvoxamine vs. placebo: 0.42 (0.04-4.67)<br>Delta: Fluvoxamine vs. placebo: 1.35 (0.73-2.48)<br>Omicron: Fluvoxamine vs. placebo: 2.32 (0.45-11.94)<br>p-value: 0.48<br><br>Alpha: Ivermectin vs. placebo: 0.15 (0.00-6.17) |

| | | |
|---|---|---|
| | | Delta: Ivermectin vs. placebo: 1.25 (0.68-2.32)<br><br>Omicron: Ivermectin vs. placebo: 0.60 (0.21-1.69)<br><br>p-value:0.08<br><br>Subgroup, SARS-CoV-2 vaccination status, HR (95% CI):<br><br>Not vaccinated: Metformin vs. placebo: 0.44 (0.24-0.80)<br><br>Vaccinated: Metformin vs. placebo: 0.85 (0.46-1.57)<br><br>p-value: 0.12<br><br>Not vaccinated: Fluvoxamine vs. placebo: 1.19 (0.52-2.71)<br><br>Vaccinated: Fluvoxamine vs. placebo: 1.50 (0.72-3.14)<br><br>p-value: 0.68<br><br>Not vaccinated: Ivermectin vs. placebo: 1.18 (0.60-2.30)<br><br>Vaccinated: Ivermectin vs. placebo: 0.65 (0.28-1.49)<br><br>p-value: 0.27 |
| Ko, 2022 | 1. Steroids<br><br>2. Antivirals<br><br>3. Hydroxychloroquine | Sensitivity analysis by ICU population, OR (95% CI):<br><br>ICU patients: 1.26 (0.51–3.11)<br><br>p-value: 0.62<br><br>Sensitivity analysis by non-invasive mechanical ventilation population, OR (95% CI):<br><br>Non-invasive mechanical ventilation patients: 0.49 (0.13-1.88)<br><br>p-value: 0.30<br><br>Sensitivity analysis by mechanical ventilation population, OR (95% CI):<br><br>Mechanical ventilation patients: 2.25 (0.21-23.87)<br><br>p-value: 0.50 |
| Nevalainen, 2022 | Antivirals | Subgroup, need for oxygen therapy, RR (95% CI):<br><br>Oxygen during admission: 1.03 (0.90-1.17)<br><br>No oxygen during admission: 0.88 (0.65-1.18) |

Table S6. Subgroup data, secondary outcomes

| Author, Year | Secondary outcome | | Subgroups/sensitivity analyses |
|---|---|---|---|
| Chuang, 2023 | Hospitalizations | Antivirals | Subgroup, Age groups, OR (95% CI):<br>≥65 years: 0.476 (0.365-0.620)<br>45–64 years: 0.333 (0.234-0.473)<br>18–44 years: 0.532 (0.373-0.759)<br>p-value: <0.001<br><br>Subgroup, Sex, OR (95% CI):<br>Male: 0.588 (0.444-0.779)<br>Female: 0.398 (0.314-0.505)<br>p-value: <0.001<br><br>Subgroup, Vaccination status, OR (95% CI):<br>Unvaccinated: 0.375 (0.294-0.488)<br>Vaccinated: 0.759 (0.569-1.011)<br>p-value: <0.001 |
| Chuang, 2023 | Fatigue | Antivirals | Subgroup, Age groups, OR (95% CI):<br>≥65 years: 0.952 (0.741-1.224)<br>45–64 years: 0.735 (0.558-0.966); p-value: <0.05<br>18–44 years: 0.845 (0.578-1.236)<br><br>Subgroup, Sex, OR (95% CI):<br>Male: 0.902 (0.697-1.167)<br>Female: 0.797 (0.644-0.988)<br>p-value: <0.05 |

| | | | |
|---|---|---|---|
| | | | <u>Subgroup, Vaccination status, OR (95% CI):</u><br><br>Unvaccinated: 0.913 (0.731-1.149)<br><br>Vaccinated: 0.833 (0.646-1.073)<br><br>p-value: >0.05 |
| Chuang, 2023 | Cognitive impairments | Antivirals | <u>Subgroup, Age groups, OR (95% CI):</u><br><br>≥65 years: 0.931 (0.669-1.297)<br><br>45–64 years: 0.540 (0.275-1.063)<br><br>18–44 years: 0.769 (0.336-1.754)<br><br>p-value: >0.05<br><br><u>Subgroup, Sex, OR (95% CI):</u><br><br>Male: 0.657 (0.417-1.036)<br><br>Female: 0.862 (0.614-1.209)<br><br>p-value: >0.05<br><br><u>Subgroup, Vaccination status, OR (95% CI):</u><br><br>Unvaccinated: 0.742 (0.527-1.053)<br><br>Vaccinated: 0.600 (0.394-0.915)<br><br>p-value: <0.05 |
| Chuang, 2023 | Anxiety/depression (Patel 2024 used for this outcome from TriNetX database) | Antivirals | <u>Subgroup, Age groups, OR (95% CI):</u><br><br>≥65 years: 1.211 (0.986-1.488)<br><br>45–64 years: 1.037 (0.860-1.253)<br><br>18–44 years: 1.085 (0.890-1.323)<br><br>p-value: >0.05<br><br><u>Subgroup, Sex, OR (95% CI):</u><br><br>Male: 0.941 (0.765-1.157)<br><br>Female: 1.140 (0.997-1.304)<br><br>p-value: >0.05 |

| | | | |
|---|---|---|---|
| | | | <u>Subgroup, Vaccination status, OR (95% CI):</u><br>Unvaccinated: 0.982 (0.846-1.136)<br>Vaccinated: 1.088 (0.920-1.287)<br>p-value: >0.05 |

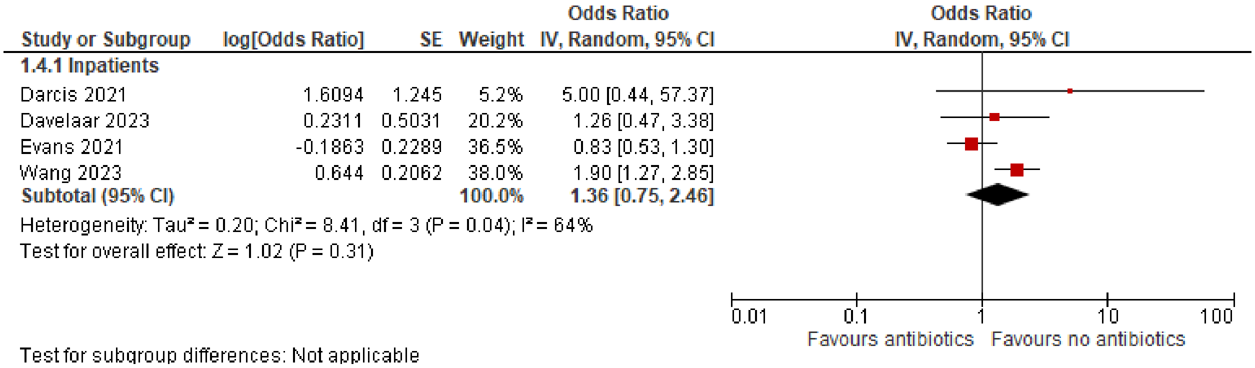

Figure S1. PCC by antibiotic use

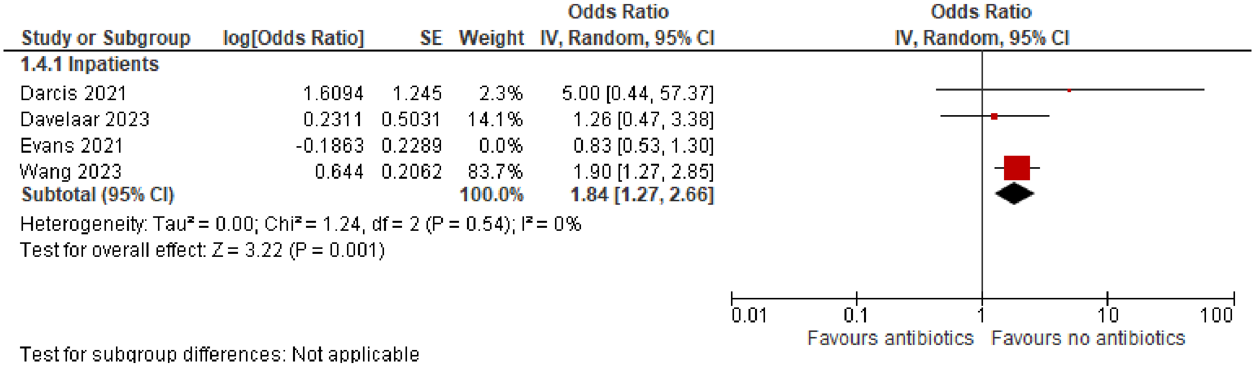

Figure S2. PCC by antibiotic use (sensitivity analysis, excluding non-recovery study [Evans 2021])

| Study or Subgroup | Antivirals Events | Antivirals Total | No antivirals Events | No antivirals Total | Weight | Risk Ratio M-H, Random, 95% CI |
|---|---|---|---|---|---|---|
| **2.1.1 Outpatients** | | | | | | |
| Chuang 2023 | 266 | 12245 | 295 | 12245 | 78.4% | 0.90 [0.77, 1.06] |
| Congdon 2023 | 58 | 250 | 62 | 250 | 21.6% | 0.94 [0.68, 1.28] |
| **Subtotal (95% CI)** | | **12495** | | **12495** | **100.0%** | **0.91 [0.79, 1.05]** |
| Total events | 324 | | 357 | | | |

Heterogeneity: Tau² = 0.00; Chi² = 0.04, df = 1 (P = 0.84); I² = 0%
Test for overall effect: Z = 1.29 (P = 0.20)

Test for subgroup differences: Not applicable

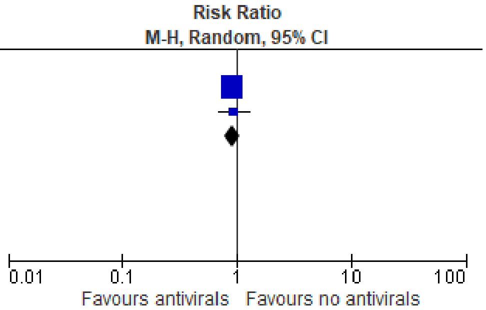

Figure S3. Fatigue by antiviral use

| Study or Subgroup | log[Odds Ratio] | SE | Weight | Odds Ratio IV, Random, 95% CI |
|---|---|---|---|---|
| **2.4.1 Inpatients** | | | | |
| Frontera 2021 | 0.7885 | 0.2684 | 52.6% | 2.20 [1.30, 3.72] |
| Milne 2021 | -0.7765 | 0.4459 | 47.4% | 0.46 [0.19, 1.10] |
| **Subtotal (95% CI)** | | | **100.0%** | **1.05 [0.23, 4.85]** |

Heterogeneity: Tau² = 1.09; Chi² = 9.04, df = 1 (P = 0.003); I² = 89%
Test for overall effect: Z = 0.06 (P = 0.95)

Test for subgroup differences: Not applicable

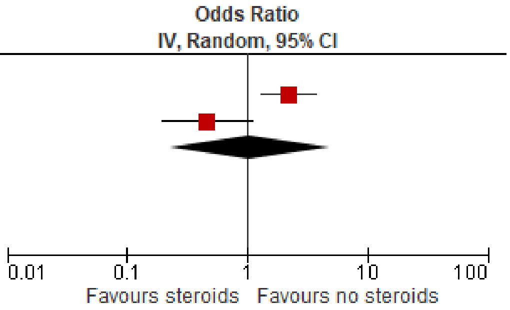

Figure S4. Fatigue by steroid use

| Study or Subgroup | Steroids Mean | Steroids SD | Steroids Total | No steroids Mean | No steroids SD | No steroids Total | Weight | Std. Mean Difference IV, Random, 95% CI |
|---|---|---|---|---|---|---|---|---|
| **2.3.1 Inpatients** | | | | | | | | |
| Berentschot 2023 | 23.1 | 8.9 | 248 | 23.7 | 9.1 | 119 | 21.5% | -0.07 [-0.29, 0.15] |
| Leavy 2023 | 36.79 | 12.23 | 738 | 36.31 | 12.87 | 489 | 78.5% | 0.04 [-0.08, 0.15] |
| **Subtotal (95% CI)** | | | **986** | | | **608** | **100.0%** | **0.02 [-0.09, 0.12]** |

Heterogeneity: Tau² = 0.00; Chi² = 0.70, df = 1 (P = 0.40); I² = 0%
Test for overall effect: Z = 0.31 (P = 0.76)

Test for subgroup differences: Not applicable

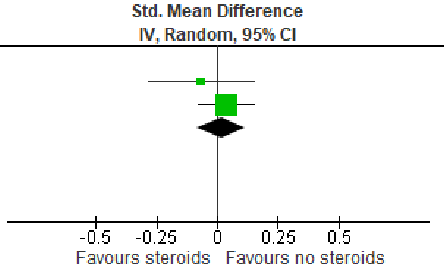

Figure S5. Fatigue by steroid use (continuous)

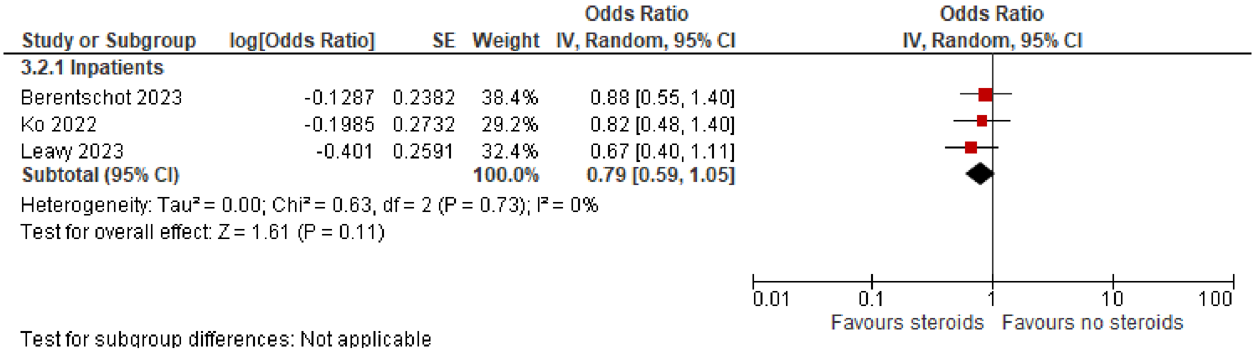

Figure S6. Breathlessness/Dyspnea by steroid use

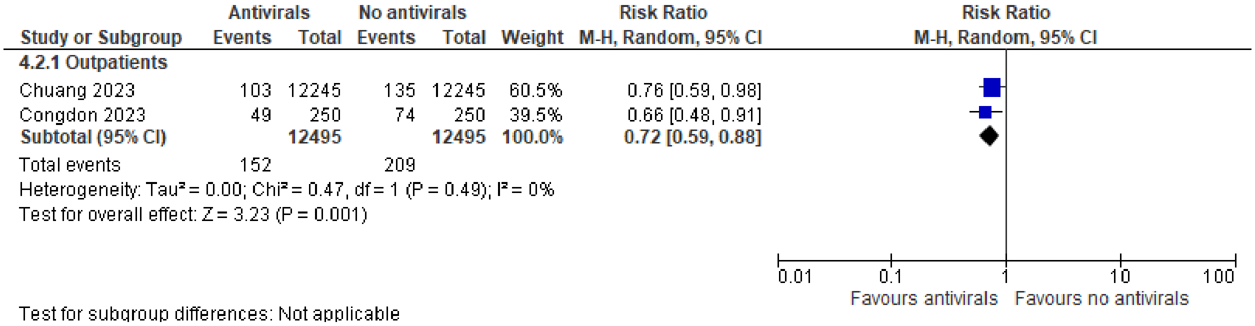

Figure S7. Cognitive impairment by antiviral use

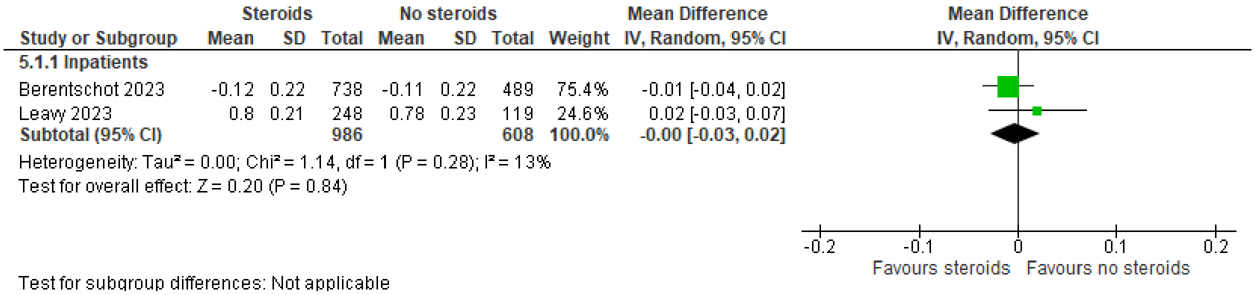

Figure S8. Health-related quality of life by steroid use

Searches

Table S7. Search for Medline [Ovid]

Ovid MEDLINE(R) ALL <1946 to February 09, 2024>

| | | |
|---|---|---|
| 1 | Post-Acute COVID-19 Syndrome/ and (pc or th).fs. | 413 |
| 2 | ((post covid* or post coronavirus* or long covid* or long coronavirus*) adj2 (prevent* or prophyla* or treatment* or management*)).ab. | 322 |
| 3 | 1 or 2 | 709 |
| 4 | Post-Acute COVID-19 Syndrome/ | 2985 |
| 5 | (long COVID* or long coronavirus* or longCOVID* or longcoronavirus*).ti,ab,kf. | 4733 |
| 6 | (sequela* adj5 (COVID* or coronavirus* or corona virus* or SARS-COV-2 or SARS-COV2 or SARSCOV-2 or SARSCOV2)).ti,ab,kf. | 2682 |
| 7 | ((post or chronic or long term or longterm or persistent*) adj3 (COVID* or coronavirus* or corona virus* or SARS-COV-2 or SARS-COV2 or SARSCOV-2 or SARSCOV2) adj4 (condition* or sequela* or syndrome* or subsyndrome* or clinical syndrome* or disorder* or symptom* or outcome* or clinical outcome* or function* or followup or follow-up or subtyp* or sub-typ* or phenotyp* or complication* or survivor*)).ti,ab,kf. | 5003 |
| 8 | ((post acute or postacute or late complication*) adj3 (COVID* or coronavirus* or corona virus* or SARS-COV-2 or SARS-COV2 or SARSCOV-2 or SARSCOV2)).ti,ab,kf. | 1860 |
| 9 | PASC.ti,kf. | 369 |
| 10 | post-COVID*.kf. | 1769 |
| 11 | or/4-10 | 10222 |
| 12 | COVID-19/ or SARS-CoV-2/ | 260259 |
| 13 | (COVID* or coronavirus* or corona virus* or 2019nCoV or 19nCoV or COVID19* or COVID or SARS-COV-2 or SARSCOV-2 or SARS-COV2 or SARSCOV2 or SARS coronavirus 2 or Severe Acute Respiratory Syndrome Coronavirus 2 or Severe Acute Respiratory Syndrome Corona Virus 2).ti,ab,kf,nm,ot,ox,rx,px. | 419256 |
| 14 | 12 or 13 | 424813 |
| 15 | (((post acute or postacute or sub-acute or subacute or chronic or long or longterm or late) adj sequela*) or PASC).ti,ab,kf. | 4057 |
| 16 | (long haul* or longhaul*).ti,ab,kf. | 1360 |
| 17 | ((persist* or long* or residual or prolonged) adj8 ((olfactory or chemosensor*) adj (disorder* or dysfunction*))).ti,ab,kf. | 237 |
| 18 | (post* or chronic* or long or longterm or sequela*).ti,ab,kf. | 5904171 |
| 19 | (PFS or (pulmonary adj3 fibro*) or (lung adj3 fibro*) or fatigue syndrome? or myalgic encephalomyelitis or ME-CFS or ME?CFS or (postural adj3 tachycardia*) or POTS or MIS-C or MIS-A or PIMS or PIMSTS or PIMS-TS).ti,ab,kf. | 106191 |
| 20 | ((multisystem* or multi-system*) adj3 (inflamm* or hyperinflamm*)).ti,ab,kf. | 4302 |

| | | |
|---|---|---|
| 21 | 18 and (19 or 20) | 33771 |
| 22 | or/15-17,21 | 39152 |
| 23 | 14 and 22 [adapted from CADTH Literature Search Strategy - Post-COVID-19 Condition Treatment and Management Rapid Scoping Review] | 3470 |
| 24 | exp vitamins/ | 357185 |
| 25 | exp vitamin b 6/ | 16870 |
| 26 | exp vitamin b 12/ | 24019 |
| 27 | exp riboflavin/ | 15829 |
| 28 | exp niacinamide/ | 17447 |
| 29 | exp thiamine/ | 12590 |
| 30 | exp folic acid/ | 42459 |
| 31 | exp ascorbic acid/ | 45940 |
| 32 | exp Dietary Supplements/ | 103759 |
| 33 | vitamin?.tw,kf. | 261733 |
| 34 | riboflavin.tw,kf. | 12151 |
| 35 | (niacinamide or enduramide or nicobion or nicotinamide or papulex).tw,kf. | 27099 |
| 36 | (thiamine or aneurin or thiamin).tw,kf. | 14892 |
| 37 | (folic acid or folacin or folate or folvite or pteroylglutamic acid).tw,kf. | 49508 |
| 38 | (ascorbic acid or ferrous ascorbate or hybrin or "l-ascorbic acid" or magnesium ascorbate or magnesium ascorbicum or "magnesium di-l-ascorbate" or magnorbin or sodium ascorbate).tw,kf. | 39929 |
| 39 | ((diet* or nutrition* or herbal* or food?) adj3 supplement*).tw,kf. | 80615 |
| 40 | exp Probiotics/ or probiotic*.tw,kf. | 45818 |
| 41 | Zinc/ | 68000 |
| 42 | Magnesium/ | 70431 |
| 43 | Selenium/ or selenium*.tw,kf. | 40905 |
| 44 | (mineral* or zinc or magnesium).tw,kf. | 421275 |
| 45 | Ubiquinone/ or "coenzyme q10".tw,kf. | 12120 |
| 46 | exp Diet Therapy/ or ((diet* or nutrition*) adj2 (advice* or therap* or counsel* or change* or intervention* or program*)).tw,kf. or (dietitian* or nutritionist*).tw,kf. | 137402 |
| 47 | exp anti-inflammatory agents/ | 576037 |
| 48 | (((antiinflammator* or anti inflammator*) adj2 (d rug? or pharmaceutical? or agent? or substance? or medicin* or prescription?)) or NSAID or NSAIDs).tw,kf. | 47269 |

| | | |
|---|---|---|
| 49 | exp adrenal cortex hormones/ | 428303 |
| 50 | (adrenal cortex hormone? or corticoid? or cortical steroid? or cortico steroid? or corticosteroid? or dermocorticosteroid?).tw,kf. | 140188 |
| 51 | (dexamethasone? or decaject? or decameth? or decaspray? of dexasone? or dexpak? or hexadecadrol? or hexadrol? or maxidex? or methylfluorprednisolone? or millicorten? or oradexon?).tw,kf. | 66806 |
| 52 | (prednisone or cortan or cortancyl or cutason or dacortin or decortin or decortisyl or dehydrocortisone or deltasone or encorton or encortone or enkortolon or kortancyl or liquid pred or meticorten or orasone or panafcort or panasol or predni tablinen or prednidib or predniment or prednison acsis or prednison galen or prednison hexal or pronisone or rectodelt or sone or sterapred or ultracorten or winpred or delta cortisone).tw,kf. | 33245 |
| 53 | (methylprednisolone or methylprednisolone or medrol or metipred or urbason).tw,kf. | 20354 |
| 54 | (hydrocortisone or "acticort" or "aeroseb hc" or "ala-cort" or "ala-scalp" or "alfacort" or "algicortis" or "alkindi" or "alpha derm" or "alphaderm" or "anucort-hc" or "anumed-hc" or "anutone-hc" or "aquanil hc" or "balneol-hc" or "barseb hc" or "beta-hc" or "biacort" or "cetacort" or "cobadex" or "colocort" or "compound f" or "cordicare lotion" or "coripen" or "cort dome" or "cortef" or "cortef cream" or "cortenema" or "cortibel" or "corticorenol" or "cortifan" or "cortiphate" or "cortisol" or "cortisole" or "cortispray" or "cortoderm" or "cortril" or "cotacort" or "covocort" or "cremicort-h" or "cutaderm" or "dermacrin hc lotion" or "dermaid" or "derm-aid cream" or "dermaid soft cream" or "dermocare" or "dermocortal" or "dermolate" or "dioderm" or "eczacort" or "ef cortelan" or "efcortelan" or "egocort" or "egocort cream" or "eksalb" or "eldecort" or "emo-cort" or "epicort" or "ficortril" or "filocot" or "flexicort" or "glycort" or "gly-cort" or "hc no. 1" or "hc no. 4" or "h-cort" or "hebcort" or "hebcort v" or "hemorrhoidal hc" or "hemril-30" or "hemril-hc uniserts" or "hi-cor" or "hidrotisona" or "hycor" or "hycort" or "hydracort" or "hydrasson" or "hydro ricortex" or "hydrocort" or "hydrocorticosteroid" or "hydrocortisate" or "hydrocortison" or "hydrocortisonum" or "hydrocortisyl" or "hydrocortone" or "hydrogalen" or "hydrokort" or "hydrokortison" or "hydro-rx" or "hydrotopic" or "hysone" or "hytisone" or "hytone" or "hytone lotion" or "incortin h" or "instacort 10" or "kyypakkaus" or "lacticare hc" or "lemnis fatty cream hc" or "lenirit" or "medihaler cort" or "medihaler duo" or "medrocil" or "mildison" or "mitocortyl demangeaisons" or "munitren" or "nogenic hc" or "novohydrocort" or "nsc 10483" or "nsc 741" or "nsc10483" or "nutracort" or "optef" or "otosone f" or "penecort" or "plenadren" or "prepcort" or "prevex hc" or "pro cort" or "procort" or "proctocort" or "proctosert hc" or "proctosol-hc" or "proctosone" or "proctozone hc" or "procutan" or "rectasol-hc" or "rectocort" or "rederm" or "sanatison" or "scalp-aid" or "schericur" or "schericur 0.25%" or "scherosone f" or "sistral hydrocort" or "skincalm" or "stie-cort" or "substance m" or "synacort" or "texacort" or "triburon-hc" or "unicort" or "vasocort").tw,kf. | 89634 |
| 55 | nitric oxide/ | 97093 |
| 56 | (nitric oxide or endogenous nitrate vasodilator or mononitrogen monoxide or nitrogen monoxide or genosyl or inomax or noxivent).tw,kf. | 169020 |
| 57 | exp prednisolone/ | 54193 |
| 58 | (prednisolone or "adelcort" or "antisolon" or "antisolone" or "aprednislon" or "aprednislone" or "benisolon" or "benisolone" or "berisolon" or "berisolone" or "caberdelta" or "capsoid" or "codelcortone" or "co-hydeltra" or "compresolon" or "cortadeltona" or "cortadeltone" or "cortalone" or "cortelinter" or "cortisolone" or "cotolone" or "dacortin" or "dacortin h" or "dacrotin" or "decaprednil" or "decortin h" or "decortril" or "dehydro cortex" or "dehydro hydrocortison" or "dehydro hydrocortisone" or "dehydrocortex" or "dehydrocortisol" or "dehydrocortisole" or "dehydrohydrocortison" or "dehydrohydrocortisone" or "delcortol" or "delta 1 hydrocortisone" or "delta cortef" or "delta cortril" or "delta ef cortelan" or "delta f" or "delta hycortol" or "delta hydrocortison" or "delta hydrocortisone" or "delta ophticor" or "delta stab" or "delta1 dehydrocortisol" or "delta1 dehydrohydrocortisone" or "delta1 hydrocortisone" or "deltacortef" or "deltacortenolo" or "deltacortil" or "deltacortoil" or "deltacortril" or "deltaderm" or "deltaglycortril" or "deltahycortol" or "deltahydrocortison" or "deltahydrocortisone" or "deltaophticor" or "deltasolone" or "deltastab" or "deltidrosol" or "deltisilone" or "deltisolon" or "deltisolone" or "deltolasson" or "deltolassone" or "deltosona" or "deltosone" or "depo-predate" or "dermosolon" or "dhasolone" or "di adreson f" or "di adresone f" or "diadreson f" or "diadresone f" or "dicortol" or "domucortone" or "encortelon" or "encortelone" or | 33244 |

"encortolon" or "equisolon" or "fernisolone-p" or "glistelone" or "hefasolon" or "hostacortin h" or "hostacortin h vet" or "hydeltra" or "hydeltrone" or "hydrelta" or "hydrocortancyl" or "hydrocortidelt" or "hydrodeltalone" or "hydrodeltisone" or "hydroretrocortin" or "hydroretrocortine" or "inflanefran" or "insolone" or "keteocort h" or "key-pred" or "lenisolone" or "leocortol" or "liquipred" or "lygal kopftinktur n" or "mediasolone" or "meprisolon" or "meprisolone" or "metacortalon" or "metacortalone" or "metacortandralon" or "metacortandralone" or "metacortelone" or "meti derm" or "meticortelone" or "metiderm" or "morlone" or "mydrapred" or "neo delta" or "nisolon" or "nisolone" or "nsc 9120" or "nsc9120" or "opredsone" or "panafcortelone" or "panafcortolone" or "panafort" or "paracortol" or "phlogex" or "pre cortisyl" or "preconin" or "precortalon" or "precortancyl" or "precortisyl" or "predacort 50" or "predaject-50" or "predalone 50" or "predartrina" or "predartrine" or "predate-50" or "predeltilone" or "predisole" or "predisyr" or "pred-ject-50" or "predne dome" or "prednecort" or "prednedome" or "prednelan" or "predni coelin" or "predni h tablinen" or "prednicoelin" or "prednicort" or "prednicortelone" or "prednifor drops" or "predni-helvacort" or "predniment" or "predniretard" or "prednis" or "prednisil" or "prednisolon" or "prednisolona" or "prednisolone alcohol" or "prednisolone h" or "prednisolone oleosae sr 82" or "prednivet" or "prednorsolon" or "prednorsolone" or "predonine" or "predorgasolona" or "predorgasolone" or "prelon" or "prelone" or "prenilone" or "prenin" or "prenolone" or "preventan" or "prezolon" or "rubycort" or "scherisolon" or "scherisolona" or "serilone" or "solondo" or "solone" or "solupren" or "soluprene" or "spiricort" or "spolotane" or "sterane" or "sterolone" or "supercortisol" or "supercortizol" or "taracortelone" or "walesolone" or "wysolone").tw,kf.

59 exp aspirin/ 49059

60 (aspirin or acetylsalicylic acid or "8-hour bayer" or "acenterine" or "acesal" or "acetan" or "acetard" or "aceticil" or "aceticyl" or "acetilum" or "acetonyl" or "acetophen" or "acetosal" or "acetosalicylic acid" or "acetosalin" or "acetosalum" or "acetyl salicylate" or "acetyl salicylic acid" or "acetylic salicylic acid" or "acetylin" or "acetylo" or "acetylo salicylic acid" or "acetylon" or "acetylosalicylic acid" or "acetylsal" or "acetylsalicyclic acid" or "acetylsalicyl" or "acetylsalicylate" or "acetylsalicylate strontium" or "acetylsalicylic acid plus glycine" or "acetylsalicylic acid sodium salt" or "acetylsalicylic acid strontium salt" or "acetylsalycic acid" or "acetylsalycylic acid" or "acetysal" or "acidulatum" or "acidum acetyl salicylicum" or "acidum acetylosalicylicum" or "acidum acetylsalicylicum" or "actorin" or "acylpyrin" or "acylpyrine" or "acytosal" or "adiro" or "alabukun" or "alasil" or "albyl e" or "albyl minor" or "alka seltzer" or "alkaspirin" or "anasprin" or "andol" or "anopyrin" or "ansin" or "anthrom" or "aptor" or "arthralgyl" or "arthritis strength bufferin" or "asacard" or "asaetta" or "asaflow" or "asaphen" or "asapor" or "asatard" or "asawin" or "aspec" or "aspent" or "aspergum" or "aspex" or "aspilets" or "aspirem" or "aspirgran" or "aspiricor" or "aspirina" or "aspirine" or "aspirinine" or "aspirisucre" or "aspisol" or "aspo cid" or "aspro" or "aspro cardio" or "aspro clear" or "asproflash" or "asrina" or "asrivo" or "asta" or "asteric" or "asteric acid" or "astrix" or "bamyl" or "bayaspirina" or "bebesan" or "biprin" or "bokey" or "boxazin" or "breoprin" or "bufferin" or "cafenol" or "cardioasa" or "cardioasae" or "cardioaspirina" or "cartia" or "caspirin" or "catalgine" or "catalgix" or "cemerit" or "cemirit" or "claradin" or "claragine" or "colfarit" or "comoprin" or "contrheuma" or "contrheuma retard" or "darosal" or "depot aspirin" or "dispirin" or "dolean" or "durlaza" or "dusil" or "easprin" or "ecasil" or "ecosprin" or "ecotrin" or "egalgic" or "emocin" or "empirin" or "encaprin" or "encine em" or "endosprin" or "entaprin" or "entericin" or "enteroprin" or "enterosarine" or "enterospirine" or "entrophen" or "eskotrin" or "euthermine" or "extren" or "flamasacard" or "genasprin" or "globentyl" or "godamed" or "gotosan" or "helicon" or "herz ass" or "hjertemagnyl" or "idotyl" or "infatabs a" or "istopirin" or "istopyrine" or "ivepirine" or "juvepirine" or "keypo" or "kilios" or "kinderaspirin" or "magnecyl brus" or "magnyl dak" or "mcn r 358" or "measurin" or "mejoral" or "melabon" or "micristin" or "micropyrin" or "migrasaa" or "mikristin" or "miniasal" or "mycristin" or "naspro" or "novasen" or "nu seal" or "nuseals" or "nu-seals" or "nu-seals asa" or "ortho acetoxybenzoate" or "ortho acetoxybenzoic acid" or "ortho acetyloxybenzoate" or "ortho acetyloxybenzoic acid" or "ostoprin" or "pancemol" or "para acetylsalicylic acid" or "paracin" or "paynocil" or "pengo" or "platet 300 cleartab" or "plewin" or "polopiryna" or "premaspin" or "primaspan" or "proprin" or "pyronoval" or "reumyl" or "rhodine" or "rhonal" or "ronal" or "salacetin" or "salacetogen" or "saletin" or "salisalido" or "salospir" or "sargepirine" or "sedergine" or "sedergine forte" or "sodium acetylsalicylate" or "sodium bicarbonate acetyl salicylate" or "sodium bicarbonate acetylsalicylate" or "soldral" or "solpyron" or "solucetyl" or "solupsa" or "spren" or "super tru" or "tapal" or "temagin" or "tevapirin" or "th 2152" or "thrombo-aspilets" or "toldex retard" or "treupahlin" or "treuphalin" or "tromalyt" or "tromcor" or "turivital" or "vazalore" or "verin" or "vitalink" or "xaxa" or "zorprin").tw,kf. 66766

61 (adalimumab or "abp 501" or "abp501" or "abrilada" or "abt d2e7" or "abtd2e7" or "adaly" or "amgevita" or "amjevita" or "amsparity" or "avt 02" or "avt02" or "bat 1406" or "bat1406" or "bax 2923" or "bax 923" or "bax2923" or "bax923" or "bi 695501" or "bi695501" or "chs 1420" or "chs1420" or "cinnora" or "ct p17" or "ctp17" or "cyltezo" or "da 3113" 10030

or "da3113" or "dmb 3113" or "dmb3113" or "exemptia" or "fkb 327" or "fkb327" or "fyzoclad" or "gp 2017" or "gp2017" or "hadlima" or "halimatoz" or "hefiya" or "hlx 03" or "hlx03" or "hulio" or "humira" or "hyrimoz" or "ibi 303" or "ibi303" or "idacio" or "imraldi" or "kromeya" or "lu 200134" or "lu200134" or "m 923" or "m923" or "mabura" or "monoclonal antibody d2e7" or "msb 11022" or "msb11022" or "ons 3010" or "ons3010" or "pf 06410293" or "pf 6410293" or "pf06410293" or "pf6410293" or "raheara" or "sb 5" or "sb5" or "solymbic" or "trudexa" or "zrc 3197" or "zrc3197").tw,kf.

62 (celecoxib or "aclarex" or "artilog" or "artroxil" or "caditar" or "celcox" or "celebra" or "celebreks" or "celebrex" or "celecox" or "celib" or "celora" or "coxel" or "coxid" or "dilox" or "eliflam" or "elyxyb" or "lexfin" or "onsenal" or "sc 58635" or "sc58635" or "solexa" or "ym 177" or "ym177" or "zycel").tw,kf. 8420

63 (diclofenac or "abdiflam" or "abitren" or "acuflam" or "akis" or "algipatch" or "algistick" or "algopain eze" or "algoplast" or "allvoran" or "almiral" or "alonpin" or "antalcalm" or "apo-diclofenac ec" or "arcanafenac" or "arthrifen" or "artren" or "artrenac" or "artrites" or "assaren" or "athrofen" or "ba 47210" or "ba47210" or "berafen gel" or "berifen" or "betaren" or "bolabomin" or "calozan" or "catanac" or "catas" or "cencenag" or "clo-far" or "clofec" or "clofen" or "clonac" or "clonaren" or "clonodifen" or "cordralan" or "curinflam" or "ddl plaster" or "declophen" or "decrol" or "deflamat" or "deflam-k" or "delphinac" or "denaclof" or "depain" or "diceus" or "dicipan" or "diclac" or "diclax" or "diclo" or "diclobasan" or "diclobene" or "diclod" or "diclodent" or "diclodoc" or "diclodolor" or "diclofen" or "diclofen cremogel" or "diclofenac rekur" or "diclofenac resin" or "diclofenac resinate" or "diclofenac sodium" or "dicloflam" or "diclohexal" or "dicloin" or "diclomax" or "diclomol" or "diclon" or "diclopax" or "diclophenac sodium" or "diclopuren" or "dicloral" or "dicloran gel" or "diclorecip" or "dicloren" or "dicloreum" or "diclosan sr" or "diclosian" or "diclotec" or "diclowal" or "dicsnal" or "difen" or "difena" or "difenac" or "difenol gel" or "difnal k" or "dioxaflex" or "dioxaflex retard" or "divoltar" or "dixol" or "doflastad" or "doflex" or "dolaren" or "dolflam-retard" or "dolo voltaren" or "doloflam" or "dolotren" or "doragon" or "dosanac" or "duravolten" or "dycon sr" or "dyloject" or "ecofenac" or "econac" or "effekton" or "eflagen" or "epifenac" or "eslofen" or "evadol" or "evinopon" or "feloran" or "fenac" or "fenadium" or "fenaspec" or "flameril" or "flexagen" or "flogofenac" or "flogosin d" or "flogozan" or "fortfen sr" or "freejex" or "gp 45840" or "grofenac" or "hizemin" or "imflac" or "inac gel" or "indicam" or "inflamac" or "inflanac" or "isv 205" or "isv205" or "jonac gel" or "kadiflam" or "kinespir" or "klofen l" or "klotaren" or "kriplex" or "lesflam" or "leviogel" or "lifenac" or "lofenac" or "lotirac" or "magluphen" or "merflam" or "modifenac" or "monoflam" or "motifene" or "naboal" or "nac gel" or "naclof" or "nacoflar" or "nadifen" or "novapirina" or "novo-difenac" or "novolten" or "ofenac" or "olfen" or "optanac" or "orthophen" or "osteoflam" or "painstop" or "panamor" or "pennsaid" or "profenac" or "relaxyl gel" or "remethan" or "renvol emulgel" or "rewodina" or "rheufenac" or "rheumafen" or "rheumatac" or "rheumatac retard" or "rhewlin" or "rhewlin sr" or "rhumalgan" or "rolactin" or "sailib" or "savismin" or "sefnac" or "slofenac" or "sodium diclofenac" or "solaraze" or "sophenoderm" or "soproxen" or "spraymik" or "sr 318t" or "staren" or "sting gel" or "tabiflex" or "tds 943" or "tds943" or "tigen plaster" or "toraren" or "traulen" or "tsudohmin" or "uniclonax" or "uniren" or "valentac" or "vartelon" or "veral" or "voldal" or "voldic" or "volero" or "volfenac" or "volna-k" or "volsaid" or "volta" or "voltadex emulgel" or "voltadvance" or "voltalen" or "voltalen emulgel" or "voltalgan" or "voltaren" or "voltarene" or "voltarenspe" or "voltarol" or "voltine" or "voltral" or "voltrix" or "voren emulgel" or "votalen" or "voveran" or "vurdon" or "wergyl" or "xenid" or "yuren" or "zolterol" or "zorvolex").tw,kf. 15849

64 dimethyl sulfoxide/ 15968

65 (dimethyl sulfoxide or "damul" or "demasorb" or "demavet" or "demeso" or "demexide" or "dimethyl sulphoxide" or "dimethylsulfoxide" or "dimethylsulphoxide" or "dimexide" or "dms 70" or "dms 90" or "dms70" or "dms90" or "dmso" or "dolicur" or "domoso" or "dromisol" or "gamasal 90" or "hyadur" or "infiltrina" or "methyl sulfoxide" or "methylsulfoxide" or "nsc 763" or "nsc763" or "rimso 100" or "rimso 50" or "somipront" or "sq 9453" or "sq9453" or "syntexan").tw,kf. 35198

66 (indomethacin or " algiflam" or "algometacin" or "amuno" or "antalgin dialicels" or "apo-indomethacin" or "areumatin" or "argilex" or "arthrexin" or "articulen" or "artracin" or "artrilona s" or "artrinovo" or "artrocid" or "asimet" or "benocid" or "betacin" or "bonidon" or "boutycin" or "catlep" or "chrono indocid" or "chronoindocid" or "confortid" or "docin" or "dolazal" or "dolazol" or "dolcidium" or "dometin" or "durametacin" or "elmego spray" or "elmetacin" or "endometacin" or "flamaret" or "flexin continus" or "grindocin" or "helvecin" or "idicin" or "im-75" or "imbrilon" or "imet" or "inacid" or "indacin" or "indalgin" or "inderapollon" or "indicin" or "indo phlogont" or "indocap" or "indocid" 37942

or "indocin" or "indocolir" or "indocollyre" or "indogesic" or "indolag" or "indolar sr" or "indolemmon" or "indo-lemmon" or "indomecin" or "indomed" or "indomee" or "indomelan" or "indomelol" or "indomet retard" or "indometacin sodium" or "indometacine" or "indomethacin" or "indomethacine" or "indomethacinum" or "indomethegan" or "indometicina mckesson" or "indometin" or "indometin depot" or "indomexum" or "indomin" or "indono" or "indoptic" or "indoptol" or "indorektal" or "indorem" or "indos" or "indosan" or "indosima" or "indosmos" or "indo-tablinen" or "indotard" or "indovis" or "indoxen" or "indoy" or "indren" or "indrenin" or "indylon" or "inflazon" or "inmetsin" or "inteban" or "lauzit" or "luiflex" or "lyo indometacin trihydrate" or "malival" or "mcn r 1166" or "mcn r1166" or "metacen" or "methacin" or "methindol" or "methindole" or "methocaps" or "metindol" or "mezolin" or "miometacen" or "mk 615" or "mk615" or "mobilan" or "novomethacin" or "osmogit" or "osmosin" or "reumacid" or "reusin" or "rheumacid" or "rheumacin" or "salinac" or "servimeta" or "sidocin" or "tannex" or "taye" or "tivorbex" or "vi-gel" or "vonum").tw,kf.

| | | |
|---|---|---|
| 67 | methotrexate/ | 41832 |
| 68 | (methotrexate or "abitrexate" or "amethopterin" or "amethopterine" or "ametopterine" or "antifolan" or "biotrexate" or "canceren" or "cl 14377" or "cl14377" or "emtexate" or "emthexat" or "emthexate" or "emtrexate" or "enthexate" or "farmitrexat" or "farmitrexate" or "farmotrex" or "folex" or "ifamet" or "imeth" or "intradose mtx" or "jylamvo" or "lantarel" or "ledertrexate" or "maxtrex" or "metex" or "methoblastin" or "methohexate" or "methotrate" or "methotrexat" or "methotrexato" or "methoxtrexate" or "methrotrexate" or "methylaminopterin" or "methylaminopterine" or "meticil" or "metoject" or "metothrexate" or "metotrexat" or "metotrexate" or "metotrexin" or "metrex" or "mexate" or "mpi 5004" or "mpi5004" or "neotrexate" or "nordimet" or "novatrex" or "nsc 740" or "nsc740" or "otrexup" or "otrexup pfs" or "rasuvo" or "reditrex" or "reumatrex" or "rheumatrex" or "texate" or "texorate" or "trexall" or "xaken" or "xatmep" or "zexate").tw,kf. | 49125 |
| 69 | (naproxen or " acusprain" or "aflamax" or "aflaxen" or "agilex" or "agilxen" or "aleve" or "alpoxen" or "alpron" or "anaprox" or "anexopen" or "apo-naproxen" or "apranax" or "apraxin" or "apronax" or "artagen" or "artron" or "artroxen" or "axer alfa" or "babel" or "bipronyl" or "bonyl" or "congex" or "crysanal" or "dafloxen" or "daprox" or "daprox entero" or "deflamox" or "dextro naproxen" or "diferbest" or "diocodal" or "dolormin fuer frauen" or "dolormin fur frauen" or "dolormin gs" or "dysmenalgit" or "dysmenalgit n" or "ec naprosyn" or "equiproxen" or "femex" or "feminax ultra" or "flanax" or "flanax forte" or "floginax" or "flonax" or "floxene" or "fuxen" or "galpharm period pain relief" or "gibixen" or "headlon" or "iraxen" or "laraflex" or "lasonil antinfiammatorio e antireumatico" or "lefaine" or "leniartil" or "levo naproxen" or "licorax" or "methoxypropiocin" or "miranax" or "momendol" or "nafasol" or "naixan" or "napolon" or "naposin" or "napreben" or "naprelan" or "napren" or "naprium" or "naprius" or "naproflam" or "naprogesic" or "naprong" or "naprontag" or "naprorex" or "naprossene" or "naprostad" or "naprosyn" or "naprosyne" or "naprovite" or "naproxeno" or "naproxi 250" or "naproxi 500" or "naproxyn" or "naprozyne" or "naprux" or "napsyn" or "napxen" or "narma" or "narocin" or "naxen" or "naxopren" or "naxyn" or "neprossin" or "norswel" or "novonaprox" or "novo-naprox" or "novuran" or "nuprafem" or "nycopren" or "pactens" or "prexan" or "priaxen" or "prodilor" or "pronaxen" or "proxen" or "proxidol" or "rahsen" or "rs 3540" or "rs 3650" or "rs3540" or "rs3650" or "sanomed" or "saritilron" or "seladin" or "shiprosyn" or "sutolin" or "synaprosyn" or "synflex" or "tohexen" or "uniflam" or "u-ritis" or "velsay" or "veradol" or "vinsen" or "wintrex" or "xenar" or "xenobid").tw,kf. | 7618 |
| 70 | Hydroxychloroquine/ | 5984 |
| 71 | (hydroxychloroquine or chloroquinol or ercoquin or hydrochloroquine or hydrocloroquine or oxychloroquine or quensyl or sn-8137 or sn8137 or tlc-19 or tlc19 or win-1258 or win1258).tw,kf. | 9295 |
| 72 | exp histamine h1 antagonists/ or exp histamine h2 antagonists/ or antihistamine*.tw,kf. | 62316 |
| 73 | exp antiviral agents/ | 404812 |
| 74 | ((antivir* or anti vir*) adj2 (drug? or pharmaceutical? or agent? or substance? or medicin* or prescription?)).tw,kf. | 29233 |
| 75 | (tocilizumab or actemra or atlizumab or lusinex or "r 1569" or r1569 or roactemra).tw,kf. | 6541 |
| 76 | (baricitinib or "incb 028050" or "incb 28050" or incb028050 or incb28050 or "ly 3009104" or ly300910 or olumiant).tw,kf. | 1458 |

| | | |
|---|---|---|
| 77 | (remdesivir or redyx or ro-7286260 or ro7286260 or veklury or gs-5734 or gs5734).tw,kf. | 3809 |
| 78 | (nirmatrelvir* or pf-07321332 or pf-7321332 or pf07321332 or pf7321332 or paxlovid or ritonavir* or a-84538 or a84538 or abt-538 or abt-84538 or abt538 or abt84538 or norvir or orb-102 or orb102 or ritovir).tw,kf. | 9125 |
| 79 | (favipiravir or avicod or avigan or favicovir or t-705 or t705).tw,kf. | 1408 |
| 80 | (oseltamivir or ebilfumin or en-241104 or en241104 or enzamir or fluvir or gs-4104 or gs4104 or gs4104002 or hgp-0919 or hgp0919 or hip-1403 or hip1403 or oseltamavir or ro-64-0796 or ro640796 or segosana or tamiflu or tamivil).tw,kf. | 4595 |
| 81 | (ganciclovir or gancyclovir or citovirax or cymevan or cymeven* or cytovene or denocin or denosine or dihydroxypropoxymethylguanine or virgan or vitrasert or zirgan).tw,kf. | 7781 |
| 82 | ((anticoagulant* or anti coagulant* or antithrombo* or anti-thrombo*) adj2 (drug? or pharmaceutical? or agent? or substance? or medicin* or prescription?)).tw,kf. | 11115 |
| 83 | exp heparin, low-molecular-weight/ | 14448 |
| 84 | (low adj3 heparin).tw,kf. | 15526 |
| 85 | (choay or depolymerized heparin or traxyparine).tw,kf. | 101 |
| 86 | (enoxaparin or clexan or clexane or decipar or inhixa or klexane or ledraxen or lovenox or neoparin or qualiop klinik or thorinane).tw,kf. | 5596 |
| 87 | Azithromycin/ | 6976 |
| 88 | (azithromycin or aratro or aruzilina or atizor or azacid or azadose or azasite or azatril or azenil or azi-sandoz or azi-teva or azibactron or azibiot or azimed or azimin or azimycin or aziprome or azirox or azirutec or azithral or azithran or azitredil or azitrocin or azitromax or azitromicin* or azitrox or azivirus or aziwill or aziwok or azomyne or azromax or aztrin or azycyna or azydrop or azylung or azytact or azyter or azythromycin or bactrazol or batif or bazyt or bezanin or ciroz or clamelle or goldamycin or inedol or infectoazit or kromicin or macromax or macrozit or makromicin or mezatrin or nobaxin or novozithron or octavax or ordipha or razimax or rezan or ribotrex or sumamed or sunamed or tobyl or tromix or trozamil or trozocina or ultreon or vinzam or xithrone or zaraxin or zaret or zarom or zedbac or zetamax or zeto or zibramax or zifin or zimacrol or zimericina or zinfect or zistic or zithrobest or Zithromax* or zithroplus or zithrotel or zithrox or zithroxyn or zitinn or zitrim or zitrobifan or zitrobiotic or zitrocin* or zitrogram or zitronova or zitrozin or zmax).tw,kf. | 12609 |
| 89 | Doxycycline/ | 11014 |
| 90 | (Doxycycline or adoxa or amermycin or apprilon or atrax or azudoxat or bactidox or banndoclin or basedillin or bassado or biocolyn or biodoxi or bronmycin or calcium-doxycycline or cloran or cyclidox or dentistar or deoxycycline or deoxymycin dispersal or deoxymykoin or deoxyoxytetracycline or desoxy oxytetracycline or desoxycycline or doinmycin or dosil or dotur or doxaciclindoxat or doxatet or doxi-sergo or doxibiotic or doxicycline or doxilin or doximed or doximycin or doxin or doxine or doxirobe or doxocycline or doxsig or doxy or doxybiocin or doxycen or doxycen retard or doxychel or doxycin or doxycyclin or doxylag or doxylin or doxymycin or doxypuren or doxytec or doxytrim or dumoxin or duracycline or efracea or esdoxin or etidoxina or gewacyclin or ibralene or idocyclin or idocyklin or interdoxin or investin or longamycin or lydox or magdrin or medomycin or mespafin or mildox or miraclin or monodox or nanodox or nordox or nsc-56228 or oracea or oraycea or paldomycin or radox or remycin or respidox or roximycin or serodoxy or servidoxine or servidoxyne or siadocin or siclidon or sigadoxin or spanor or supracyclin or supramycina or tenutan or tolexine or torymycin or tsurupioxin or unidox or veemycin or viadoxin or vibra-s or vibrabiotic or vibracina or vibradox or vibramicina or vibramycin* or vibraveineuse or vibravenos or vibravet or viradoxyl-n or wanmycin or xyrosa or zadorin or zenavod).tw,kf. | 17639 |
| 91 | exp Amoxicillin/ | 12857 |

| | | |
|---|---|---|
| 92 | (actimoxi* or amoxicillin* or amoxil or amoxycillin* or clamoxyl or hydroxyampicillin or penamox or polymox or trimox or wymox or amox-clav or amoxi-clavulanate or augmentin or clavulin or co-amoxiclav or spektramox or synulox).tw,kf. | 23075 |
| 93 | Trimethoprim, Sulfamethoxazole Drug Combination/ | 7647 |
| 94 | Vancomycin/ | 16646 |
| 95 | (abactrim or bactifor or bactrim or biseptol or centran or centrin or co-trimoxazole or drylin or eslectin or eusaprim or insozalin or kepinol or lescot or metomide or oriprim or septra or septrin or (sulfamethoxazole adj2 trimethoprim) or sulprim or sumetrolim or trimedin or trimethoprimsulfa or trimezole or trimosulfa or (diatracin or vanco-cell or vanco azupharma or vanco-saar or vancocin* or vancomicina* or vancomycin*)).tw,kf. | 50047 |
| 96 | Ceftriaxone/ or exp Fluoroquinolones/ or exp Cephalosporins/ | 82384 |
| 97 | (acantex or axone or benaxona or biotrakson or biotriax or bioxon or broadced or brospec or cef-3 or cefaflox or cefalogen or cefatriaxone or cefaxona or cefaxone or cefin or cefotal or cefotriaxon or cefotriaxone or cefriex or ceftrex or ceftrian or ceftriaxone or ceftrilem or cefxon or cephin or cephtriaxone or cerixon or cikedrix or ecotrixon or elpicef or eurocef or ferfacef or forgram or glicocef or gomcephin or grifotriaxona or incephin or keftriaxon or keptrix or lopratin or lyceft or medoxonum or megion or mesporin or monocef or nakaxone or novosef or oframax or pantrixon or retrokor or rinxofay or rocefalin or rocefin or rocephalin or Rocephin* or rocidar or rowecef or roxcef or roxon or samixon or sintrex or socef or sunflow or tacex or torocef-1 or trexofin or triaken or triax or triaxone or tricefin or tricephin or trijec or xtenda or zefaxone or zefone 250).tw,kf. | 14042 |
| 98 | (Fluoroquinolone* or Ciprofloxacin* or Fleroxacin* or Enoxacin* or Enrofloxacin* or Gatifloxacin* or Gemifloxacin* or Moxifloxacin* or Norfloxacin* or Ofloxacin* or Levofloxacin* or Pefloxacin* or (Cephalosporin* or Cefalosporin* or Cefamandoleor* or Cefoperazone* or Cefazolin* or Cefdinir* or Cefepime* or Cefonicid* or Cefsulodin* or Ceftibuten* or Cefuroxime* or Cephacetrile* or Cefotaxime* or Cephalothin* or Cephapirin* or Cephalexin* or Cefaclor* or Cefadroxil* or Cefatrizine* or Cephaloglycin* or Cephradine* or Cephaloridine* or Ceftazidime* or Cephamycin* or Cefmetazole* or Cefotetan* or Cefoxitin*)).tw,kf. | 117908 |
| 99 | Immunomodulating Agents/ or Immunosuppressive Agents/ | 105945 |
| 100 | ((immunomodula* adj2 (agent* or drug* or medication? or medicine? or compound? or therap*)) or immunomodulator? or immunotherap*).tw,kf. | 215369 |
| 101 | ((immunosuppress* or immuno-suppress* or immunodepress*) adj2 (agent* or drug* or medication? or medicine? or compound? or substance? or therap*)).tw,kf. | 55599 |
| 102 | Rivaroxaban/ | 4904 |
| 103 | (rivaroxaban or assubex or ast-8294 or ast8294 or bay-59-7939 or bay-597939 or bay59-7939 or bay59-7939 or bay597939 or bs-112 or bs112 or dst-8294 or dst8294 or jnj-39039039 or jnj39039039 or kriva or naxat or rivaro or rivarolto or rivaxa or throsaben or xanirva or xarelto or xerdoxo or xindus).tw,kf. | 7785 |
| 104 | Interleukin Inhibitors/ | 74 |
| 105 | (interleukin inhibitor? or cytokine antagonist or cytokine receptor block* or interleukin receptor antagonist?).tw,kf. | 219 |
| 106 | ((cilgavimab adj2 tixagevimab) or evusheld or "azd 1061 plus azd 8895" or "azd 1061/azd 8895" or azd-7442 or "azd 8895 plus azd 1061" or "azd 8895/azd 1061" or "azd1061 plus azd8895" or "azd1061/azd8895" or azd7442 or "azd8895 plus azd1061" or "azd8895/azd1061").tw,kf. | 265 |
| 107 | (itolizumab or alzumab or bmab-600 or bmab600 or eq-001 or eq001).tw,kf. | 62 |
| 108 | (dupilumab or bat-2406 or bat2406 or dupixent or regn-668 or regn668 or sar-231893 or sar231893).tw,kf. | 2845 |
| 109 | (pamrevlumab or fg-3019 or fg3019).tw,kf. | 52 |

| | | |
|---|---|---|
| 110 | exp antibodies, monoclonal/ | 281491 |
| 111 | ((monoclonal or clonal or hybridoma) adj3 antibod*).tw,kf. | 219461 |
| 112 | (bamlanivimab or etesevimab or "bamlanivimab/etesevimab" or "2423943-37-5" or "LY-3819253" or "LY-COV555" or "LY3819253" or "UNII-45I6OFJ8QH" or "WHO 11876" or "2423948-94-9" or "anti-Sars-cov-2 antibody JS016" or "CB6" or "JS016" or "LY COV016" or "LY-3832479" or "LY-COV016" or "LY3832479" or "NP005" or "UNII-N7Q9NLF11I" or "WHO 11873").tw,kf. | 554 |
| 113 | (casirivimab or imdevimab or "casirivimab/imdevimab" or regen-cov or "REGN-COV2" or regn10933 or regn10987 or "anti-sars-cov-2 regn-cov2" or "2415933-42-3" or "REGN-10933" or "REGN10933" or "UNII-J0FI6WE1QN" or "WHO 11861" or "2415933-40-1" or "REGN-10987" or "REGN10987" or "UNII-2Z3DQD2JHM" or "WHO 11863").tw,kf. | 426 |
| 114 | ("Sotrovimab" or "2423014-07-5" or "GSK-4182136" or "GSK4182136" or "UNII-1MTK0BPN8V" or "VIR-7831" or "VIR7831").tw,kf. | 333 |
| 115 | (amubarvimab or "BRII-196" or "BRII-198" or "DZIF-10c" or "BI 767551" or "SCTA01" or "Ty027" or "HLX70").tw,kf. | 24 |
| 116 | ("C144-LS" or "C-135-LS" or "C144-LS/C-135-LS").tw,kf. | 3 |
| 117 | ("COVI-GUARD" or "STI-1499").tw,kf. | 1 |
| 118 | ("COVI-AMG" or "sti-2020").tw,kf. | 4 |
| 119 | (enuzovimab or plutavimab or lomtegovimab or "HFB30132A").tw,kf. | 1 |
| 120 | ("ABBV-47D11" or "ABBV-2B04" or "AZD7442").tw,kf. | 36 |
| 121 | ("BI 767551" or "DZIF-10c" or "COR-101").tw,kf. | 4 |
| 122 | (baricitinib or olumiant or incb28050 or ly3009104).tw,kf. | 1456 |
| 123 | ((immune adj3 (sera or serum? or plasma)) or antisera or antiserum or immun#serum).tw,kf. | 79297 |
| 124 | exp immunoglobulins/ | 994947 |
| 125 | (immune* globulin? or immuneglobulin? or immunoglobulin?).tw,kf. | 185483 |
| 126 | ((convalescent adj2 (plasma or serum or sera)) or (plasma adj3 therap*)).tw,kf. | 14991 |
| 127 | exp ibuprofen/ | 10216 |
| 128 | (ibuprofen or abfen or "aches-n-pain" or "act-3" or actiprofen or "adex 200" or adex liqui-gels or advil or afebril or aktren or aktren spezial or algiasdin or algifor or algofen or algoflex or allipen or alvofen express or "am-fam 400" or anadin or anadvil or analgyl or anbifen or anco or andran or anflagen or antalgil or antarene or antiflam or apo-ibuprofen or aragel or "atril 300" or attritin or balkaprofen or berlistar or bestafen or betaprofen or bifen or bluton or brufanic or brufedol or brufen or brufort or brugesic or brumare or brumed or brupro or buburone or bufect or bufohexal or bupogesic or burana or butacortelone or butifen or caldolor or calprofen or cap-profen or cenbufen or codral period pain or combiflam or contraneural or cuprofen or dalsy or "dc 7034" or "dc7034" or "dg 7034" or "dg7034" or dibufen or "diffutab sr 600" or dimidon or dolan fp or dolgit or dolobene ibu or dolocyl or dolodolgit or dolofen-f or dolomax or dolormin or dolval or donjust b or dorival or druisel or easifon or ecoprofen or emflam or epobron or ergix douleur et fievre or eudorlin extra or exidol or expanfen or febratic or febryn or femapirin or fenalgic or fenbid or flamicon or flarin or froben dolore or galprofen or gelufene or gyno-neuralgin or halprin or haltran or hemagene tailleur or h-loniten or ib-100 or ibalgin or ibofen or ibosure or ibu or ibuberl or ibucalm or ibudak or ibudol or ibudolor or ibufarmalid or ibufen or ibuflam or ibufug or ibugel or ibugesic or ibukern or ibuleve or ibulgan or ibuloid or ibumetin or ibumousse or ibunin or ibupen or ibupirac or ibuprin or ibuprocin or ibuprofene or ibuprohm or ibuprom or iburon or ibusal or ibuspray or ibustar or ibusynth or ibutop or ibux or ibuxin or idyl sr or ifenin or infant's motrin or infibu or inflanor or inflanor forte or ipren or irfen or junifen or junipro or kenfen or kontraneural or lamidon or | 18252 |

| | | |
|---|---|---|
| | librofem or lidifen or liptan or lopane or malafene or maxagesic or "mcn r 1451" or medicol or medipren or mediprin or mensoton or midol or momentact or motrin or mynosedin or nagifen-d or napacetin or neobrufen or neobrufen retard or nerofen or neutropain or nobfelon or nobgen or norflam-t or noritis or norton or novogent or novoprofen or nugin or nuprin or nureflex or nurofen or optifen or opturem or ostarin or ostofen or ozonol or paduden or panafen or pedea or pediacare fever or pediprofen or perdophen pediatrie or perofen or phorpain or phorpain gel or proartinal or profen or profeno or proff or proflex or proris or provin or provon or quadrax or rafen or ranofen or rapidophen or rapidophen forte or ratiodolor or rebugen or renidon or reuvol or rhelafen or roidenin or rufen or rupan or saridon n or schufen or seclodin or solufen lidose or solvium or spalt or syntofene or tabalon or tab-profen or taskine or tatanal or tofen or trendar or umafen or unipro or upfen or uprofen or urem or viamal febbre e dolore or zafen or zofen).tw,kf. | |
| 129 | exp Selective Serotonin Reuptake Inhibitors/ | 46656 |
| 130 | (((serotonin or 5-Hydroxytryptamine or 5-HT) adj3 (reuptake-inhibitor? or uptake-inhibitor?)) or SSRI? or fluvoxamine or du-23000 or du23000 or fluoxamin? or fluroxamine?).tw,kf. | 25157 |
| 131 | exp "Serotonin and Noradrenaline Reuptake Inhibitors"/ | 5586 |
| 132 | (SNRI* or SSNRI* or ((((noradrenalin or norepinephrine) adj2 serotonin) or dual) adj3 (reuptake-inhibitor* or uptake-inhibitor*))).tw,kf. | 3297 |
| 133 | citalopram/ or escitalopram/ or fluoxetine/ or Sertraline/ | 17844 |
| 134 | Paroxetine/ or Desvenlafaxine Succinate/ or Duloxetine Hydrochloride/ or milnacipran/ or levomilnacipran/ or Venlafaxine Hydrochloride/ | 9158 |
| 135 | (celexa or citalopram* or cytalopram or seropram or escitalopram* or lexapro or fluoxetin* or prozac or sarafem or altruline or sertraline or aremis or besitran or gladem or lustral or sealdin or zoloft or aropax or paroxetin* or paxil or seroxat or (desvenlafaxine* or ellefore or khedezla or o-desmethylvenlafaxine or o-norvenlafaxine or pristiq* or cymbalta or duloxetin* or dalcipran or fetzima or impulsor or ixel or joncia or levomilnacipran* or midalcipran or milnaneurax or savella* or toledomin* or dobupal or efexor or effexor or grz5rcb1qg or venlafaxin* or trevilor or vandral)).tw,kf. | 33713 |
| 136 | stem cell transplantation/ or hematopoietic stem cell transplantation/ or mesenchymal stem cell transplantation/ or stem-cell transplant*.tw,kf. | 118748 |
| 137 | Metformin/ | 18418 |
| 138 | (metformin or anj-900 or anj900 or apophage or aron or benofomin or dabex or denkaform or deson or dextin or diabetase or diabetformin or diabetmin or diabetosan or diabex or diafat or diaformin or diaformina or diametin or diamin or dianben or diformin or diformin retard or dimefor or dimethylbiguanide or dimethyldiguanide or dmgg or dybis or efb-0027 or efb0027 or eraphage or espa-formin or euform retard or fluamine or flumamine or fornidd or fortamet or glafornil or glibudon or glifage or gliguanid or glucaminol or glucofage or glucofago or glucoform or glucoformin or glucohexal or glucoless or glucomet or glucomin or glucomine or gluconil or Glucophage* or glucostop or glucotika or gludepatic or glufor or gluformin or glukophage or glumeformin or glumet or glumetza or glupa or glustress or glyciphage or glycomet or glycon or glycoran or glyformin or glymet or haurymellin or hipoglucin or islotin or jesacrin or juformin or la-6023 or la6023 or maformin or meglucon or meguan or melbin or melformin or mellittin or merckformin or mescorit or metaformin or metfogamma or metfoliquid geriasan or metforal or metformax or metformin hydrochloride or metformina or metformine or methformin or metiguanide or metomin or metphormin or miformin or neoform or newmet or nndg or reglus-500 or riomet or risidon or rudimet or siamformet or siofor or thiabet or vimetrol or walaphage).tw,kf. | 29509 |
| 139 | Ivermectin/ | 7703 |
| 140 | (ivermectin or bibovel or cardomec or cevamec or diapec or driponin or efacti or epimek or equimec or eqvalan or eqvalen or iveraxiro or ivercare or ivergalen or ivergelan or ivermectina or ivermectol or iverscab or ivertin or ivexterm or ivomec or k-237 or k237 or loutol or manburesa or mectizan or mk-933 or mk933 or oramec or posela or quanox | 7902 |

| | | |
|---|---|---|
| | gotas or revectina or romitu or scaball or scabioral or scatol or securo or sklice or soolantra or stromectol or vona or zulima).tw,kf. | |
| 141 | Resveratrol/ | 11157 |
| 142 | (resveratrol or srt-501 or srt501).tw,kf. | 17478 |
| 143 | Naltrexone/ | 8761 |
| 144 | (naltrexone* or abernil or addex-1000 or addex1000 or adepend or antaxon or antaxone or celupan or cyto-201 or cyto201 or depade or dependex or en-1639a or en1639a or ethylex or irt-103 or irt-104 or irt103 or irt104 or lodonal or n-cyclopropylmethylnoroxymorphone or nalerona or nalorex or naltex or naltrel or naltrexin or nemexin or nodict or nutrexon or opnt-002 or opnt002 or phaltrexia or pti-901 or pti901 or regental or revez or revia or stat-200 or stat-201 or stat-205 or stat200 or stat201 or stat205 or tranalex or trexan or um-792 or um792 or vivitrex or vivitrol).tw,kf. | 8018 |
| 145 | Colchicine/ | 15228 |
| 146 | (colchicin* or colchicum-dispert or colchily or colchimedio or colchiquim or colchisol or colchysat or colcine or colcrys or colctab or colgout or colrefuz or gloperba or goutichine or goutnil or kolkicin or kolkisin or mitigare or mpc-004 or mpc004 or myinfla or tolchicine).tw,kf. | 18374 |
| 147 | or/24-146 [Drugs] | 4512809 |
| 148 | stem cell transplantation/ or hematopoietic stem cell transplantation/ or mesenchymal stem cell transplantation/ or stem-cell transplant*.tw,kf. | 118748 |
| 149 | Low-Level Light Therapy/ or (photobiomodulation or photo-biomodulation or photo-bio-modulation or ((low-power or low-level or low-energy or low-intensity or soft or cold) adj2 (light or laser) adj2 (therap* or irradiat*)) or (laser adj2 biostimulat*) or lllt or ((laser adj2 phototherap*) or biostimulat*)).tw,kf. | 12707 |
| 150 | rehabilitation/ or "activities of daily living"/ or exercise therapy/ or neurological rehabilitation/ or rehabilitation.tw,kf. | 324600 |
| 151 | ((cognitive or neurologic*) adj4 (rehab* or motor-therap*)).tw,kf. | 6192 |
| 152 | self-management/ or self-care/ | 41534 |
| 153 | (((patient or self) adj (manag* or treat*)) or self care).tw,kf. | 98075 |
| 154 | patient education as topic/ or patient medication knowledge/ or health literacy/ or (patient* adj3 (educat* or knowledg* or literacy or literate or behavior* or behaviour* or attitud* or belief* or believ*)).tw,kf. | 189906 |
| 155 | self-help groups/ | 9652 |
| 156 | (((self help or support*) adj3 (group? or club? or servic* or social*)) or (therap* adj3 (club? or group* or social*))).tw,kf. | 152649 |
| 157 | disease management/ or pain management/ | 84625 |
| 158 | ((manag* or cope or coping) adj3 (symptom* or disease* or condition* or pain or discomfort*)).tw,kf. | 168604 |
| 159 | exercise/ or exp running/ or swimming/ or exp walking/ or exp *physical fitness/ or yoga/ or "physical education and training"/ or physical therapy specialty/ | 278094 |
| 160 | ("physical activit*" or walk* or pedestrian* or bicycl* or cycling or cyclist* or biking or bike* or "active lifestyle*" or "aerobic fitness" or "aerobic exercise*" or (running not "running water") or runner* or jog* or swim* or yoga or (physical* adj2 (activit* or active or exercise*)) or ((exercise* or fitness or aerobic*) adj2 (regimen* or training or intervention* or program* or class* or course* or train* or rehab*))).tw,kf. | 603608 |
| 161 | (physiotherap* or ((physical or physio) adj1 (therap* or treat*))).tw,kf. | 69817 |

| | | |
|---|---|---|
| 162 | health services, indigenous/ | 4229 |
| 163 | ((indigenous or metis or aboriginal or inuit or (native adj (america* or alaska* or hawaii* or canad*)) or first nation* or first people*) and (healthcare or (health adj2 (care* or service* or program*)) or healing* or medic* or therap* or remed*)).tw,kf. | 19801 |
| 164 | exp complementary therapies/ or exp medicine, traditional/ or herbal medicine/ or plants, medicinal/ | 300049 |
| 165 | ((alternat* or complementary or folk or traditional or holistic or chinese or african or tribal) adj2 (medic* or therap* or healing* or treat* or remed*)).tw,kf. | 209928 |
| 166 | (hypnotism or hypnosis or hypnotherapy).tw,kf. | 9901 |
| 167 | (ayurved* or kampo or kanpo or acupunctur* or homeopath*).tw,kf. | 41312 |
| 168 | ((botanical* or herb* or plant or plants or plantlet* or root or roots or natural) adj2 (drug? or extract? or healing* or ingredient? or medic* or preparation* or product or products or remedies or remedy* or supplement* or treat* or therap*)).tw,kf. | 218151 |
| 169 | Rest/ or (rest or (avoid* adj2 (exercise* or exert*))).tw,kf. | 187955 |
| 170 | exp mental health services/ or exp social work/ or psychiatric rehabilitation/ or psychiatric nursing/ | 137662 |
| 171 | ((mental health or emotion* or psych* or wellbeing or well-being or stress* or wellness* or anxiet* or depress* or obsess* or ocd or mood* or posttrauma* or post-trauma* or ptsd or schizo* or personality disorder* or bipolar* or adhd or attention deficit* or addict*) adj3 (care or service? or support? or treat* or therap* or psychotherap* or counsel* or hotline*)).tw,kf. | 377613 |
| 172 | (lifestyle adj3 (change* or intervention* or advice*)).tw,kf. | 28194 |
| 173 | exp *aftercare/ or *outpatients/ | 145603 |
| 174 | (aftercare or after care or follow-up care or referral?).tw,kf. | 160551 |
| 175 | *models, organizational/ or exp *delivery of health care/ | 742029 |
| 176 | ((healthcare or health care or clinical* or public health or community) adj4 (approach* or model? or service? or deliver* or system* or distribut* or framework? or guideline?)).tw,kf. | 515060 |
| 177 | (recovery adj2 (team* or clinic? or centre? or center? or model* or program*)).ti,ab,kf. | 4419 |
| 178 | ((interdisciplin* or inter-disciplin* or cross-disciplin* or multidiscipline* or multi-disciplin*) adj3 (team* or clinic? or centre? or center? or model* or program*)).ti,ab,kf. | 14507 |
| 179 | ((care or treatment or healthcare) adj2 (model* or path* or plan*)).ti,ab,kf. | 194653 |
| 180 | (care adj2 (coordinat* or co-ordinat*)).ti,ab,kf. | 13318 |
| 181 | (patient-centred care or patient-centered care or case manag*).ti,ab,kf. | 27737 |
| 182 | exp "Referral and Consultation"/ | 87810 |
| 183 | (refer* adj4 (specialist? or clinic? or therapy or therapist or counsel*)).tw,kf. | 30032 |
| 184 | or/148-183 [other interventions] | 4078061 |
| 185 | (11 or 23) and (147 or 184) | 4288 |
| 186 | 3 or 185 | 4644 |

| | | |
|---|---|---|
| 187 | Epidemiologic studies/ | 9486 |
| 188 | exp case control studies/ | 1480736 |
| 189 | exp cohort studies/ | 2571431 |
| 190 | Case control.tw. | 159674 |
| 191 | (cohort adj (study or studies)).tw. | 340085 |
| 192 | Cohort analy$.tw. | 12663 |
| 193 | (Follow up adj (study or studies)).tw. | 57598 |
| 194 | (observational adj (study or studies)).tw. | 172857 |
| 195 | (prospect* adj (study or studies)).tw. | 213782 |
| 196 | Longitudinal.tw. | 338108 |
| 197 | Retrospective.tw. | 790016 |
| 198 | or/187-197 [Adapted from OVID observational studies filter] | 3471467 |
| 199 | (Randomized Controlled Trial or Controlled Clinical Trial or Pragmatic Clinical Trial or Equivalence Trial or Clinical Trial, Phase III).pt. | 704265 |
| 200 | exp Randomized Controlled Trial/ | 610088 |
| 201 | exp Randomized Controlled Trials as Topic/ | 170776 |
| 202 | Controlled Clinical Trial/ | 95551 |
| 203 | exp Controlled Clinical Trials as Topic/ | 176494 |
| 204 | Random Allocation/ | 107070 |
| 205 | Double-Blind Method/ | 177493 |
| 206 | Single-Blind Method/ | 33241 |
| 207 | Placebos/ | 35934 |
| 208 | Control Groups/ | 2087 |
| 209 | (random* or sham or placebo*).ti,ab,hw,kf,kw. | 1878649 |
| 210 | ((singl* or doubl*) adj (blind* or dumm* or mask*)).ti,ab,hw,kf. | 273138 |
| 211 | ((tripl* or trebl*) adj (blind* or dumm* or mask*)).ti,ab,hw,kf. | 1766 |
| 212 | (control* adj3 (study or studies or trial* or group*)).ti,ab,kf. | 1275703 |
| 213 | (Nonrandom* or non random* or non-random* or quasi-random* or quasirandom*).ti,ab,hw,kf. | 56985 |
| 214 | allocated.ti,ab,hw. | 87490 |
| 215 | ((open label or open-label) adj5 (study or studies or trial*)).ti,ab,hw,kf. | 46907 |
| 216 | ((equivalence or superiority or non-inferiority or noninferiority) adj3 (study or studies or trial*)).ti,ab,hw,kf. | 12991 |

| | | |
|---|---|---|
| 217 | (pragmatic study or pragmatic studies).ti,ab,hw,kf,kw. | 638 |
| 218 | ((pragmatic or practical) adj3 trial*).ti,ab,hw,kf. | 8171 |
| 219 | ((quasiexperimental or quasi-experimental) adj3 (study or studies or trial*)).ti,ab,hw,kf. | 13277 |
| 220 | (phase adj3 (III or "3") adj3 (study or studies or trial*)).ti,hw,kf. | 36306 |
| 221 | or/199-220 [Adapted from "Strings attached: CADTH database search filters"] | 2691325 |
| 222 | 186 and (198 or 221) | 1569 |
| 223 | case reports/ or (case-stud* or case-report*).jw. or (case* and report*).ti. | 2511796 |
| 224 | (address or autobiography or bibliography or biography or case reports or classical article or clinical trial, veterinary or clinical trials, veterinary as topic or editorial or historical article or interview or news or newspaper article or observational study, veterinary).pt. | 3706107 |
| 225 | 222 not (223 or 224) | 1524 |
| 226 | limit 225 to yr="2022 - 2024" | 1239 |
| 227 | (202205* or 202206* or 202207* or 202208* or 202209* or 20221* or 2023* or 2024*).ez,dt,ed. | 3064509 |
| 228 | 226 and 227 | 1103 |

### Table S8. Search from Embase [Ovid]

Embase <1974 to 2024 February 09>

| | | |
|---|---|---|
| 1 | long COVID/dm, dt, th, pc | 556 |
| 2 | ((post covid* or post coronavirus* or long covid* or long coronavirus*) adj2 (prevent* or prophyla* or treatment* or management)).ab. | 413 |
| 3 | 1 or 2 | 904 |
| 4 | Long covid/ | 6847 |
| 5 | (long COVID* or long coronavirus* or longCOVID* or longcoronavirus*).ti,ab,kf. | 5869 |
| 6 | (sequela* adj5 (COVID* or coronavirus* or corona virus* or SARS-COV-2 or SARS-COV2 or SARSCOV-2 or SARSCOV2)).ti,ab,kf. | 3508 |
| 7 | ((post or chronic or long term or longterm) adj3 (COVID* or coronavirus* or corona virus* or SARS-COV-2 or SARS-COV2 or SARSCOV-2 or SARSCOV2) adj4 (condition* or sequela* or syndrome* or subsyndrome* or clinical syndrome* or disorder* or symptom* or outcome* or clinical outcome* or function* or followup or follow-up or subtyp* or sub-typ* or phenotyp* or complication* or survivor*)).ti,ab,kf. | 6440 |
| 8 | ((post acute or postacute or late complication*) adj3 (COVID* or coronavirus* or corona virus* or SARS-COV-2 or SARS-COV2 or SARSCOV-2 or SARSCOV2)).ti,ab,kf. | 2357 |
| 9 | PASC.ti,kf. | 430 |
| 10 | post-COVID*.kf. | 1855 |

| | | |
|---|---|---|
| 11 | (post-COVID* adj fatigue).ti,ab,kf. | 45 |
| 12 | or/4-11 | 14166 |
| 13 | exp Coronavirus disease 2019/ | 383874 |
| 14 | exp Severe acute respiratory syndrome coronavirus 2/ | 109044 |
| 15 | (COVID* or coronavirus* or corona virus* or 2019nCoV or 19nCoV or COVID19* or COVID or SARS-COV-2 or SARSCOV-2 or SARS-COV2 or SARSCOV2 or SARS coronavirus 2 or Severe Acute Respiratory Syndrome Coronavirus 2 or Severe Acute Respiratory Syndrome Corona Virus 2).ti,ab,kf,ot. | 477967 |
| 16 | or/13-15 | 511459 |
| 17 | (((post acute or postacute or sub-acute or subacute or chronic or long or longterm or late) adj sequela*) or PASC).ti,ab,kf. | 5046 |
| 18 | (long haul* or longhaul*).ti,ab,kf. | 1567 |
| 19 | ((persist* or long* or residual or prolonged) adj8 ((olfactory or chemosensor*) adj (disorder* or dysfunction*))).ti,ab,kf. | 274 |
| 20 | (post* or chronic* or long or longterm or sequela*).ti,ab,kf. | 7967312 |
| 21 | (PFS or (pulmonary adj3 fibro*) or (lung adj3 fibro*) or fatigue syndrome? or myalgic encephalomyelitis or ME-CFS or ME?CFS or (postural adj3 tachycardia*) or POTS or MIS-C or MIS-A or PIMS or PIMSTS or PIMS-TS).ti,ab,kf. | 203855 |
| 22 | ((multisystem* or multi-system*) adj3 (inflamm* or hyperinflamm*)).ti,ab,kf. | 6458 |
| 23 | 20 and (21 or 22) | 67969 |
| 24 | or/17-19,23 | 74504 |
| 25 | 16 and 24 | 5051 |
| 26 | exp *vitamin/ | 301224 |
| 27 | exp *folic acid/ | 22253 |
| 28 | *dietary supplement/ | 7882 |
| 29 | vitamin?.tw,kf. | 326137 |
| 30 | riboflavin.tw,kf. | 12729 |
| 31 | (niacinamide or enduramide or nicobion or nicotinamide or papulex).tw,kf. | 29463 |
| 32 | (thiamine or aneurin or thiamin).tw,kf. | 14859 |
| 33 | (folic acid or folacin or folate or folvite or pteroylglutamic acid).tw,kf. | 62402 |
| 34 | (ascorbic acid or ferrous ascorbate or hybrin or "l-ascorbic acid" or magnesium ascorbate or magnesium ascorbicum or "magnesium di-l-ascorbate" or magnorbin or sodium ascorbate).tw,kf. | 43636 |
| 35 | ((diet* or nutrition* or herbal* or food?) adj3 supplement*).tw,kf. | 100263 |
| 36 | exp probiotic agent/ | 55741 |
| 37 | exp probiotic agent/ or probiotic*.tw,kf. | 66494 |

| | | |
|---|---|---|
| 38 | Zinc/ or Magnesium/ or Selenium/ or selenium*.tw,kf. | 262159 |
| 39 | (mineral* or zinc or magnesium).tw,kf. | 503713 |
| 40 | Ubiquinone/ or "coenzyme q10".tw,kf. | 14205 |
| 41 | exp *antiinflammatory agent/ | 861548 |
| 42 | (((antiinflammator* or anti inflammator*) adj2 (drug? or pharmaceutical? or agent? or substance? or medicin* or prescription?)) or NSAID or NSAIDs).tw,kf. | 119071 |
| 43 | exp *corticosteroid/ | 318495 |
| 44 | (adrenal cortex hormone? or corticoid? or cortical steroid? or cortico steroid? or corticosteroid? or dermocorticosteroid?).tw,kf. | 201481 |
| 45 | (dexamethasone? or decaject? or decameth? or decaspray? of dexasone? or dexpak? or hexadecadrol? or hexadrol? or maxidex? or methylfluorprednisolone? or millicorten? or oradexon?).tw,kf. | 96721 |
| 46 | (prednisone or cortan or cortancyl or cutason or dacortin or decortin or decortisyl or dehydrocortisone or deltasone or encorton or encortone or enkortolon or kortancyl or liquid pred or meticorten or orasone or panafcort or panasol or predni tablinen or prednidib or predniment or prednison acsis or prednison galen or prednison hexal or pronisone or rectodelt or sone or sterapred or ultracorten or winpred or delta cortisone).tw,kf. | 63181 |
| 47 | (methylprednisolone or methylprednisolone or medrol or metipred or urbason).tw,kf. | 37613 |
| 48 | (hydrocortisone or "acticort" or "aeroseb hc" or "ala-cort" or "ala-scalp" or "alfacort" or "algicortis" or "alkindi" or "alpha derm" or "alphaderm" or "anucort-hc" or "anumed-hc" or "anutone-hc" or "aquanil hc" or "balneol-hc" or "barseb hc" or "beta-hc" or "biacort" or "cetacort" or "cobadex" or "colocort" or "compound f" or "cordicare lotion" or "coripen" or "cort dome" or "cortef" or "cortef cream" or "cortenema" or "cortibel" or "corticorenol" or "cortifan" or "cortiphate" or "cortisol" or "cortisole" or "cortispray" or "cortoderm" or "cortril" or "cotacort" or "covocort" or "cremicort-h" or "cutaderm" or "dermacrin hc lotion" or "dermaid" or "derm-aid cream" or "dermaid soft cream" or "dermocare" or "dermocortal" or "dermolate" or "dioderm" or "eczacort" or "ef cortelan" or "efcortelan" or "egocort" or "egocort cream" or "eksalb" or "eldecort" or "emo-cort" or "epicort" or "ficortril" or "filocot" or "flexicort" or "glycort" or "gly-cort" or "hc no. 1" or "hc no. 4" or "h-cort" or "hebcort" or "hebcort v" or "hemorrhoidal hc" or "hemril-30" or "hemril-hc uniserts" or "hi-cor" or "hidrotisona" or "hycor" or "hycort" or "hydracort" or "hydrasson" or "hydro ricortex" or "hydrocort" or "hydrocorticosteroid" or "hydrocortisate" or "hydrocortison" or "hydrocortisonum" or "hydrocortisyl" or "hydrocortone" or "hydrogalen" or "hydrokort" or "hydrokortison" or "hydro-rx" or "hydrotopic" or "hysone" or "hytisone" or "hytone" or "hytone lotion" or "incortin h" or "instacort 10" or "kyypakkaus" or "lacticare hc" or "lemnis fatty cream hc" or "lenirit" or "medihaler cort" or "medihaler duo" or "medrocil" or "mildison" or "mitocortyl demangeaisons" or "munitren" or "nogenic hc" or "novohydrocort" or "nsc 10483" or "nsc 741" or "nsc10483" or "nutracort" or "optef" or "otosone f" or "penecort" or "plenadren" or "prepcort" or "prevex hc" or "pro cort" or "procort" or "proctocort" or "proctosert hc" or "proctosol-hc" or "proctosone" or "proctozone hc" or "procutan" or "rectasol-hc" or "rectocort" or "rederm" or "sanatison" or "scalp-aid" or "schericur" or "schericur 0.25%" or "scherosone f" or "sistral hydrocort" or "skincalm" or "stie-cort" or "substance m" or "synacort" or "texacort" or "triburon-hc" or "unicort" or "vasocort").tw,kf. | 112716 |
| 49 | (nitric oxide or endogenous nitrate vasodilator or mononitrogen monoxide or nitrogen monoxide or genosyl or inomax or noxivent).tw,kf. | 215893 |
| 50 | (prednisolone or "adelcort" or "antisolon" or "antisolone" or "aprednislon" or "aprednislone" or "benisolon" or "benisolone" or "berisolon" or "berisolone" or "caberdelta" or "capsoid" or "codelcortone" or "co-hydeltra" or "compresolon" or "cortadeltona" or "cortadeltone" or "cortalone" or "cortelinter" or "cortisolone" or "cotolone" or "dacortin" or "dacortin h" or "dacrotin" or "decaprednil" or "decortin h" or "decortril" or "dehydro cortex" or "dehydro hydrocortison" or "dehydro hydrocortisone" or "dehydrocortex" or "dehydrocortisol" or "dehydrocortisole" or "dehydrohydrocortison" or "dehydrohydrocortisone" or "delcortol" or "delta 1 hydrocortisone" or "delta cortef" or "delta cortril" or "delta ef cortelan" or "delta f" or "delta hycortol" or "delta hydrocortison" or "delta hydrocortisone" or | 49769 |

"delta ophticor" or "delta stab" or "delta1 dehydrocortisol" or "delta1 dehydrohydrocortisone" or "delta1 hydrocortisone" or "deltacortef" or "deltacortenolo" or "deltacortil" or "deltacortoil" or "deltacortril" or "deltaderm" or "deltaglycortril" or "deltahycortol" or "deltahydrocortison" or "deltahydrocortisone" or "deltaophticor" or "deltasolone" or "deltastab" or "deltidrosol" or "deltisilone" or "deltisolon" or "deltisolone" or "deltolasson" or "deltolassone" or "deltosona" or "deltosone" or "depo-predate" or "dermosolon" or "dhasolone" or "di adreson f" or "di adresone f" or "diadreson f" or "diadresone f" or "dicortol" or "domucortone" or "encortelon" or "encortelone" or "encortolon" or "equisolon" or "fernisolone-p" or "glistelone" or "hefasolon" or "hostacortin h" or "hostacortin h vet" or "hydeltra" or "hydeltrone" or "hydrelta" or "hydrocortancyl" or "hydrocortidelt" or "hydrodeltalone" or "hydrodeltisone" or "hydroretrocortin" or "hydroretrocortine" or "inflanefran" or "insolone" or "keteocort h" or "key-pred" or "lenisolone" or "leocortol" or "liquipred" or "lygal kopftinktur n" or "mediasolone" or "meprisolon" or "meprisolone" or "metacortalon" or "metacortalone" or "metacortandralon" or "metacortandralone" or "metacortelone" or "meti derm" or "meticortelone" or "metiderm" or "morlone" or "mydrapred" or "neo delta" or "nisolon" or "nisolone" or "nsc 9120" or "nsc9120" or "opredsone" or "panafcortelone" or "panafcortolone" or "panafort" or "paracortol" or "phlogex" or "pre cortisyl" or "preconin" or "precortalon" or "precortancyl" or "precortisyl" or "predacort 50" or "predaject-50" or "predalone 50" or "predartrina" or "predartrine" or "predate-50" or "predeltilone" or "predisole" or "predisyr" or "pred-ject-50" or "predne dome" or "prednecort" or "prednedome" or "prednelan" or "predni coelin" or "predni h tablinen" or "prednicoelin" or "prednicort" or "prednicortelone" or "prednifor drops" or "predni-helvacort" or "predniment" or "predniretard" or "prednis" or "prednisil" or "prednisolon" or "prednisolona" or "prednisolone alcohol" or "prednisolone h" or "prednisolone oleosae sr 82" or "prednivet" or "prednorsolon" or "prednorsolone" or "predonine" or "predorgasolona" or "predorgasolone" or "prelon" or "prelone" or "prenilone" or "prenin" or "prenolone" or "preventan" or "prezolon" or "rubycort" or "scherisolon" or "scherisolona" or "serilone" or "solondo" or "solone" or "solupren" or "soluprene" or "spiricort" or "spolotane" or "sterane" or "sterolone" or "supercortisol" or "supercortizol" or "taracortelone" or "walesolone" or "wysolone").tw,kf.

51 (aspirin or acetylsalicylic acid or "8-hour bayer" or "acenterine" or "acesal" or "acetan" or "acetard" or "aceticil" or "aceticyl" or "acetilum" or "acetonyl" or "acetophen" or "acetosal" or "acetosalicylic acid" or "acetosalin" or "acetosalum" or "acetyl salicylate" or "acetyl salicylic acid" or "acetylic salicylic acid" or "acetylin" or "acetylo" or "acetylo salicylic acid" or "acetylon" or "acetylosalicylic acid" or "acetylsal" or "acetylsalicyclic acid" or "acetylsalicyl" or "acetylsalicylate" or "acetylsalicylate strontium" or "acetylsalicylic acid plus glycine" or "acetylsalicylic acid sodium salt" or "acetylsalicylic acid strontium salt" or "acetylsalycic acid" or "acetylsalycylic acid" or "acetysal" or "acidulatum" or "acidum acetyl salicylicum" or "acidum acetylosalicylicum" or "acidum acetylsalicylicum" or "actorin" or "acylpyrin" or "acylpyrine" or "acytosal" or "adiro" or "alabukun" or "alasil" or "albyl e" or "albyl minor" or "alka seltzer" or "alkaspirin" or "anasprin" or "andol" or "anopyrin" or "ansin" or "anthrom" or "aptor" or "arthralgyl" or "arthritis strength bufferin" or "asacard" or "asaetta" or "asaflow" or "asaphen" or "asapor" or "asatard" or "asawin" or "aspec" or "aspent" or "aspergum" or "aspex" or "aspilets" or "aspirem" or "aspirgran" or "aspiricor" or "aspirina" or "aspirine" or "aspirinine" or "aspirisucre" or "aspisol" or "aspo cid" or "aspro" or "aspro cardio" or "aspro clear" or "asproflash" or "asrina" or "asrivo" or "asta" or "asteric" or "asteric acid" or "astrix" or "bamyl" or "bayaspirina" or "bebesan" or "biprin" or "bokey" or "boxazin" or "breoprin" or "bufferin" or "cafenol" or "cardioasa" or "cardioasae" or "cardioaspirina" or "cartia" or "caspirin" or "catalgine" or "catalgix" or "cemerit" or "cemirit" or "claradin" or "claragine" or "colfarit" or "comoprin" or "contrheuma" or "contrheuma retard" or "darosal" or "depot aspirin" or "dispirin" or "dolean" or "durlaza" or "dusil" or "easprin" or "ecasil" or "ecosprin" or "ecotrin" or "egalgic" or "emocin" or "empirin" or "encaprin" or "encine em" or "endosprin" or "entaprin" or "entericin" or "enteroprin" or "enterosarine" or "enterospirine" or "entrophen" or "eskotrin" or "euthermine" or "extren" or "flamasacard" or "genasprin" or "globentyl" or "godamed" or "gotosan" or "helicon" or "herz ass" or "hjertemagnyl" or "idotyl" or "infatabs a" or "istopirin" or "istopyrine" or "ivepirine" or "juvepirine" or "keypo" or "kilios" or "kinderaspirin" or "magnecyl brus" or "magnyl dak" or "mcn r 358" or "measurin" or "mejoral" or "melabon" or "micristin" or "micropyrin" or "migrasaa" or "mikristin" or "miniasal" or "mycristin" or "naspro" or "novasen" or "nu seal" or "nuseals" or "nu-seals" or "nu-seals asa" or "ortho acetoxybenzoate" or "ortho acetoxybenzoic acid" or "ortho acetyloxybenzoate" or "ortho acetyloxybenzoic acid" or "ostoprin" or "pancemol" or "para acetylsalicylic acid" or "paracin" or "paynocil" or "pengo" or "platet 300 cleartab" or "plewin" or "polopiryna" or "premaspin" or "primaspan" or "proprin" or "pyronoval" or "reumyl" or "rhodine" or "rhonal" or "ronal" or "salacetin" or "salacetogen" or "saletin" or "salisalido" or "salospir" or "sargepirine" or "sedergine" or "sedergine forte" or "sodium acetylsalicylate" or "sodium bicarbonate acetyl salicylate" or "sodium bicarbonate acetylsalicylate" or "soldral" or "solpyron" or "solucetyl" or "solupsa" or "spren" or "super tru" 147604

or "tapal" or "temagin" or "tevapirin" or "th 2152" or "thrombo-aspilets" or "toldex retard" or "treupahlin" or "treuphalin" or "tromalyt" or "tromcor" or "turivital" or "vazalore" or "verin" or "vitalink" or "xaxa" or "zorprin").tw,kf.

52 (adalimumab or "abp 501" or "abp501" or "abrilada" or "abt d2e7" or "abtd2e7" or "adaly" or "amgevita" or "amjevita" or "amsparity" or "avt 02" or "avt02" or "bat 1406" or "bat1406" or "bax 2923" or "bax 923" or "bax2923" or "bax923" or "bi 695501" or "bi695501" or "chs 1420" or "chs1420" or "cinnora" or "ct p17" or "ctp17" or "cyltezo" or "da 3113" or "da3113" or "dmb 3113" or "dmb3113" or "exemptia" or "fkb 327" or "fkb327" or "fyzoclad" or "gp 2017" or "gp2017" or "hadlima" or "halimatoz" or "hefiya" or "hlx 03" or "hlx03" or "hulio" or "humira" or "hyrimoz" or "ibi 303" or "ibi303" or "idacio" or "imraldi" or "kromeya" or "lu 200134" or "lu200134" or "m 923" or "m923" or "mabura" or "monoclonal antibody d2e7" or "msb 11022" or "msb11022" or "ons 3010" or "ons3010" or "pf 06410293" or "pf 6410293" or "pf06410293" or "pf6410293" or "raheara" or "sb 5" or "sb5" or "solymbic" or "trudexa" or "zrc 3197" or "zrc3197").tw,kf. 28009

53 (celecoxib or "aclarex" or "artilog" or "artroxil" or "caditar" or "celcox" or "celebra" or "celebreks" or "celebrex" or "celecox" or "celib" or "celora" or "coxel" or "coxid" or "dilox" or "eliflam" or "elyxyb" or "lexfin" or "onsenal" or "sc 58635" or "sc58635" or "solexa" or "ym 177" or "ym177" or "zycel").tw,kf. 14009

54 (diclofenac or "abdiflam" or "abitren" or "acuflam" or "akis" or "algipatch" or "algistick" or "algopain eze" or "algoplast" or "allvoran" or "almiral" or "alonpin" or "antalcalm" or "apo-diclofenac ec" or "arcanafenac" or "arthrifen" or "artren" or "artrenac" or "artrites" or "assaren" or "athrofen" or "ba 47210" or "ba47210" or "berafen gel" or "berifen" or "betaren" or "bolabomin" or "calozan" or "catanac" or "catas" or "cencenag" or "clo-far" or "clofec" or "clofen" or "clonac" or "clonaren" or "clonodifen" or "cordralan" or "curinflam" or "ddl plaster" or "declophen" or "decrol" or "deflamat" or "deflam-k" or "delphinac" or "denaclof" or "depain" or "diceus" or "dicipan" or "diclac" or "diclax" or "diclo" or "diclobasan" or "diclobene" or "diclod" or "diclodent" or "diclodoc" or "diclodolor" or "diclofen" or "diclofen cremogel" or "diclofenac rekur" or "diclofenac resin" or "diclofenac resinate" or "diclofenac sodium" or "dicloflam" or "diclohexal" or "dicloin" or "diclomax" or "diclomol" or "diclon" or "diclopax" or "diclophenac sodium" or "diclopuren" or "dicloral" or "dicloran gel" or "diclorecip" or "dicloren" or "dicloreum" or "diclosan sr" or "diclosian" or "diclotec" or "diclowal" or "dicsnal" or "difen" or "difena" or "difenac" or "difenol gel" or "difnal k" or "dioxaflex" or "dioxaflex retard" or "divoltar" or "dixol" or "doflastad" or "doflex" or "dolaren" or "dolflam-retard" or "dolo voltaren" or "doloflam" or "dolotren" or "doragon" or "dosanac" or "duravolten" or "dycon sr" or "dyloject" or "ecofenac" or "econac" or "effekton" or "eflagen" or "epifenac" or "eslofen" or "evadol" or "evinopon" or "feloran" or "fenac" or "fenadium" or "fenaspec" or "flameril" or "flexagen" or "flogofenac" or "flogosin d" or "flogozan" or "fortfen sr" or "freejex" or "gp 45840" or "grofenac" or "hizemin" or "imflac" or "inac gel" or "indicam" or "inflamac" or "inflanac" or "isv 205" or "isv205" or "jonac gel" or "kadiflam" or "kinespir" or "klofen l" or "klotaren" or "kriplex" or "lesflam" or "leviogel" or "lifenac" or "lofenac" or "lotirac" or "magluphen" or "merflam" or "modifenac" or "monoflam" or "motifene" or "naboal" or "nac gel" or "naclof" or "nacoflar" or "nadifen" or "novapirina" or "novo-difenac" or "novolten" or "ofenac" or "olfen" or "optanac" or "orthophen" or "osteoflam" or "painstop" or "panamor" or "pennsaid" or "profenac" or "relaxyl gel" or "remethan" or "renvol emulgel" or "rewodina" or "rheufenac" or "rheumafen" or "rheumatac" or "rheumatac retard" or "rhewlin" or "rhewlin sr" or "rhumalgan" or "rolactin" or "sailib" or "savismin" or "sefnac" or "slofenac" or "sodium diclofenac" or "solaraze" or "sophenoderm" or "soproxen" or "spraymik" or "sr 318t" or "staren" or "sting gel" or "tabiflex" or "tds 943" or "tds943" or "tigen plaster" or "toraren" or "traulen" or "tsudohmin" or "uniclonax" or "uniren" or "valentac" or "vartelon" or "veral" or "voldal" or "voldic" or "volero" or "volfenac" or "volna-k" or "volsaid" or "volta" or "voltadex emulgel" or "voltadvance" or "voltalen" or "voltalen emulgel" or "voltalgan" or "voltaren" or "voltarene" or "voltarenspe" or "voltarol" or "voltine" or "voltral" or "voltrix" or "voren emulgel" or "votalen" or "voveran" or "vurdon" or "wergyl" or "xenid" or "yuren" or "zolterol" or "zorvolex").tw,kf. 24901

55 (dimethyl sulfoxide or "damul" or "demasorb" or "demavet" or "demeso" or "demexide" or "dimethyl sulphoxide" or "dimethylsulfoxide" or "dimethylsulphoxide" or "dimexide" or "dms 70" or "dms 90" or "dms70" or "dms90" or "dmso" or "dolicur" or "domoso" or "dromisol" or "gamasal 90" or "hyadur" or "infiltrina" or "methyl sulfoxide" or "methylsulfoxide" or "nsc 763" or "nsc763" or "rimso 100" or "rimso 50" or "somipront" or "sq 9453" or "sq9453" or "syntexan").tw,kf. 46043

56 (indomethacin or " algiflam" or "algometacin" or "amuno" or "antalgin dialicels" or "apo-indomethacin" or "areumatin" or "argilex" or "arthrexin" or "articulen" or "artracin" or "artrilona s" or "artrinovo" or "artrocid" or "asimet" or 47351

| | | |
|---|---|---|
| | "benocid" or "betacin" or "bonidon" or "boutycin" or "catlep" or "chrono indocid" or "chronoindocid" or "confortid" or "docin" or "dolazal" or "dolazol" or "dolcidium" or "dometin" or "durametacin" or "elmego spray" or "elmetacin" or "endometacin" or "flamaret" or "flexin continus" or "grindocin" or "helvecin" or "idicin" or "im-75" or "imbrilon" or "imet" or "inacid" or "indacin" or "indalgin" or "inderapollon" or "indicin" or "indo phlogont" or "indocap" or "indocid" or "indocin" or "indocolir" or "indocollyre" or "indogesic" or "indolag" or "indolar sr" or "indolemmon" or "indo-lemmon" or "indomecin" or "indomed" or "indomee" or "indomelan" or "indomelol" or "indomet retard" or "indometacin sodium" or "indometacine" or "indomethacin" or "indomethacine" or "indomethacinum" or "indomethegan" or "indometicina mckesson" or "indometin" or "indometin depot" or "indomexum" or "indomin" or "indono" or "indoptic" or "indoptol" or "indorektal" or "indorem" or "indos" or "indosan" or "indosima" or "indosmos" or "indo-tablinen" or "indotard" or "indovis" or "indoxen" or "indoy" or "indren" or "indrenin" or "indylon" or "inflazon" or "inmetsin" or "inteban" or "lauzit" or "luiflex" or "lyo indometacin trihydrate" or "malival" or "mcn r 1166" or "mcn r1166" or "metacen" or "methacin" or "methindol" or "methindole" or "methocaps" or "metindol" or "mezolin" or "miometacen" or "mk 615" or "mk615" or "mobilan" or "novomethacin" or "osmogit" or "osmosin" or "reumacid" or "reusin" or "rheumacid" or "rheumacin" or "salinac" or "servimeta" or "sidocin" or "tannex" or "taye" or "tivorbex" or "vi-gel" or "vonum").tw,kf. | |
| 57 | (methotrexate or "abitrexate" or "amethopterin" or "amethopterine" or "ametopterine" or "antifolan" or "biotrexate" or "canceren" or "cl 14377" or "cl14377" or "emtexate" or "emthexat" or "emthexate" or "emtrexate" or "enthexate" or "farmitrexat" or "farmitrexate" or "farmotrex" or "folex" or "ifamet" or "imeth" or "intradose mtx" or "jylamvo" or "lantarel" or "ledertrexate" or "maxtrex" or "metex" or "methoblastin" or "methohexate" or "methotrate" or "methotrexat" or "methotrexato" or "methoxtrexate" or "methrotrexate" or "methylaminopterin" or "methylaminopterine" or "meticil" or "metoject" or "metothrexate" or "metotrexat" or "metotrexate" or "metotrexin" or "metrex" or "mexate" or "mpi 5004" or "mpi5004" or "neotrexate" or "nordimet" or "novatrex" or "nsc 740" or "nsc740" or "otrexup" or "otrexup pfs" or "rasuvo" or "reditrex" or "reumatrex" or "rheumatrex" or "texate" or "texorate" or "trexall" or "xaken" or "xatmep" or "zexate").tw,kf. | 84549 |
| 58 | (naproxen or " acusprain" or "aflamax" or "aflaxen" or "agilex" or "agilxen" or "aleve" or "alpoxen" or "alpron" or "anaprox" or "anexopen" or "apo-naproxen" or "apranax" or "apraxin" or "apronax" or "artagen" or "artron" or "artroxen" or "axer alfa" or "babel" or "bipronyl" or "bonyl" or "congex" or "crysanal" or "dafloxen" or "daprox" or "daprox entero" or "deflamox" or "dextro naproxen" or "diferbest" or "diocodal" or "dolormin fuer frauen" or "dolormin fur frauen" or "dolormin gs" or "dysmenalgit" or "dysmenalgit n" or "ec naprosyn" or "equiproxen" or "femex" or "feminax ultra" or "flanax" or "flanax forte" or "floginax" or "flonax" or "floxene" or "fuxen" or "galpharm period pain relief" or "gibixen" or "headlon" or "iraxen" or "laraflex" or "lasonil antinfiammatorio e antireumatico" or "lefaine" or "leniartil" or "levo naproxen" or "licorax" or "methoxypropiocin" or "miranax" or "momendol" or "nafasol" or "naixan" or "napolon" or "naposin" or "napreben" or "naprelan" or "napren" or "naprium" or "naprius" or "naproflam" or "naprogesic" or "naprong" or "naprontag" or "naprorex" or "naprossene" or "naprostad" or "naprosyn" or "naprosyne" or "naprovite" or "naproxeno" or "naproxi 250" or "naproxi 500" or "naproxyn" or "naprozyne" or "naprux" or "napsyn" or "napxen" or "narma" or "narocin" or "naxen" or "naxopren" or "naxyn" or "neprossin" or "norswel" or "novonaprox" or "novo-naprox" or "novuran" or "nuprafem" or "nycopren" or "pactens" or "prexan" or "priaxen" or "prodilor" or "pronaxen" or "proxen" or "proxidol" or "rahsen" or "rs 3540" or "rs 3650" or "rs3540" or "rs3650" or "sanomed" or "saritilron" or "seladin" or "shiprosyn" or "sutolin" or "synaprosyn" or "synflex" or "tohexen" or "uniflam" or "u-ritis" or "velsay" or "veradol" or "vinsen" or "wintrex" or "xenar" or "xenobid").tw,kf. | 12424 |
| 59 | *hydroxychloroquine/ | 6890 |
| 60 | (hydroxychloroquine or chloroquinol or ercoquin or hydrochloroquine or hydrocloroquine or oxychloroquine or quensyl or sn-8137 or sn8137 or tlc-19 or tlc19 or win-1258 or win1258).tw,kf. | 16569 |
| 61 | exp *histamine H1 receptor antagonist/ or exp *histamine H2 receptor antagonist/ or antihistamine*.tw,kf. | 127161 |
| 62 | exp *antivirus agent/ | 493849 |
| 63 | ((antivir* or anti vir*) adj2 (drug? or pharmaceutical? or agent? or substance? or medicin* or prescription?)).tw,kf. | 37696 |
| 64 | *tocilizumab/ | 5743 |

| | | |
|---|---|---|
| 65 | (tocilizumab or actemra or atlizumab or lusinex or "r 1569" or r1569 or roactemra).tw,kf. | 15060 |
| 66 | *baricitinib/ | 1473 |
| 67 | (baricitinib or "incb 028050" or "incb 28050" or incb028050 or incb28050 or "ly 3009104" or ly300910 or olumiant).tw,kf. | 2931 |
| 68 | *remdesivir/ | 1598 |
| 69 | (remdesivir or redyx or ro-7286260 or ro7286260 or veklury or gs-5734 or gs5734).tw,kf. | 5444 |
| 70 | *nirmatrelvir plus ritonavir/ | 592 |
| 71 | (nirmatrelvir* or pf-07321332 or pf-7321332 or pf07321332 or pf7321332 or paxlovid or ritonavir* or a-84538 or a84538 or abt-538 or abt-84538 or abt538 or abt84538 or norvir or orb-102 or orb102 or ritovir).tw,kf. | 14643 |
| 72 | *favipiravir/ | 700 |
| 73 | (favipiravir or avicod or avigan or favicovir or t-705 or t705).tw,kf. | 1955 |
| 74 | *oseltamivir/ | 2733 |
| 75 | (oseltamivir or ebilfumin or en-241104 or en241104 or enzamir or fluvir or gs-4104 or gs4104 or gs4104002 or hgp-0919 or hgp0919 or hip-1403 or hip1403 or oseltamavir or ro-64-0796 or ro640796 or segosana or tamiflu or tamivil).tw,kf. | 7838 |
| 76 | *ganciclovir/ | 5489 |
| 77 | (ganciclovir or gancyclovir or citovirax or cymevan or cymeven* or cytovene or denocin or denosine or dihydroxypropoxymethylguanine or virgan or vitrasert or zirgan).tw,kf. | 12021 |
| 78 | ((anticoagulant* or anti coagulant* or antithrombo* or anti-thrombo*) adj2 (drug? or pharmaceutical? or agent? or substance? or medicin* or prescription?)).tw,kf. | 17118 |
| 79 | exp *low molecular weight heparin/ | 17585 |
| 80 | (low adj3 heparin).tw,kf. | 24892 |
| 81 | (choay or depolymerized heparin or traxyparine).tw,kf. | 371 |
| 82 | (enoxaparin or clexan or clexane or decipar or inhixa or klexane or ledraxen or lovenox or neoparin or qualiop klinik or thorinane).tw,kf. | 13954 |
| 83 | *azithromycin/ | 7976 |
| 84 | (azithromycin or aratro or aruzilina or atizor or azacid or azadose or azasite or azatril or azenil or azi-sandoz or azi-teva or azibactron or azibiot or azimed or azimin or azimycin or aziprome or azirox or azirutec or azithral or azithran or azitredil or azitrocin or azitromax or azitromicin* or azitrox or azivirus or aziwill or aziwok or azomyne or azromax or aztrin or azycyna or azydrop or azylung or azytact or azyter or azythromycin or bactrazol or batif or bazyt or bezanin or ciroz or clamelle or goldamycin or inedol or infectoazit or kromicin or macromax or macrozit or makromicin or mezatrin or nobaxin or novozithron or octavax or ordipha or razimax or rezan or ribotrex or sumamed or sunamed or tobyl or tromix or trozamil or trozocina or ultreon or vinzam or xithrone or zaraxin or zaret or zarom or zedbac or zetamax or zeto or zibramax or zifin or zimacrol or zimericina or zinfect or zistic or zithrobest or Zithromax* or zithroplus or zithrotel or zithrox or zithroxyn or zitinn or zitrim or zitrobifan or zitrobiotic or zitrocin* or zitrogram or zitronova or zitrozin or zmax).tw,kf. | 20775 |
| 85 | *doxycycline/ | 12381 |

| | | |
|---|---|---|
| 86 | (Doxycycline or adoxa or amermycin or apprilon or atrax or azudoxat or bactidox or banndoclin or basedillin or bassado or biocolyn or biodoxi or bronmycin or calcium-doxycycline or cloran or cyclidox or dentistar or deoxycycline or deoxymycin dispersal or deoxymykoin or deoxyoxytetracycline or desoxy oxytetracycline or desoxycycline or doinmycin or dosil or dotur or doxaciclindoxat or doxatet or doxi-sergo or doxibiotic or doxicycline or doxilin or doximed or doximycin or doxin or doxine or doxirobe or doxocycline or doxsig or doxy or doxybiocin or doxycen or doxycen retard or doxychel or doxycin or doxycyclin or doxylag or doxylin or doxymycin or doxypuren or doxytec or doxytrim or dumoxin or duracycline or efracea or esdoxin or etidoxina or gewacyclin or ibralene or idocyclin or idocyklin or interdoxin or investin or longamycin or lydox or magdrin or medomycin or mespafin or mildox or miraclin or monodox or nanodox or nordox or nsc-56228 or oracea or oraycea or paldomycin or radox or remycin or respidox or roximycin or serodoxy or servidoxine or servidoxyne or siadocin or siclidon or sigadoxin or spanor or supracyclin or supramycina or tenutan or tolexine or torymycin or tsurupioxin or unidox or veemycin or viadoxin or vibra-s or vibrabiotic or vibracina or vibradox or vibramicina or vibramycin* or vibraveineuse or vibravenos or vibravet or viradoxyl-n or wanmycin or xyrosa or zadorin or zenavod).tw,kf. | 28088 |
| 87 | exp *amoxicillin/ | 16305 |
| 88 | (actimoxi* or amoxicillin* or amoxil or amoxycillin* or clamoxyl or hydroxyampicillin or penamox or polymox or trimox or wymox or amox-clav or amoxi-clavulanate or augmentin or clavulin or co-amoxiclav or spektramox or synulox).tw,kf. | 39349 |
| 89 | *cotrimoxazole/ | 17216 |
| 90 | *vancomycin/ | 21867 |
| 91 | (abactrim or bactifor or bactrim or biseptol or centran or centrin or co-trimoxazole or drylin or eslectin or eusaprim or insozalin or kepinol or lescot or metomide or oriprim or septra or septrin or (sulfamethoxazole adj2 trimethoprim) or sulprim or sumetrolim or trimedin or trimethoprimsulfa or trimezole or trimosulfa or (diatracin or vanco-cell or vanco azupharma or vanco-saar or vancocin* or vancomicina* or vancomycin*)).tw,kf. | 76390 |
| 92 | *ceftriaxone/ or exp *cephalosporin derivative/ or exp *quinolone derivative/ | 119906 |
| 93 | (acantex or axone or benaxona or biotrakson or biotriax or bioxon or broadced or brospec or cef-3 or cefaflox or cefalogen or cefatriaxone or cefaxona or cefaxone or cefin or cefotal or cefotriaxon or cefotriaxone or cefriex or ceftrex or ceftrian or ceftriaxone or ceftrilem or cefxon or cephin or cephtriaxone or cerixon or cikedrix or ecotrixon or elpicef or eurocef or ferfacef or forgram or glicocef or gomcephin or grifotriaxona or incephin or keftriaxon or keptrix or lopratin or lyceft or medoxonum or megion or mesporin or monocef or nakaxone or novosef or oframax or pantrixon or retrokor or rinxofay or rocefalin or rocefin or rocephalin or Rocephin* or rocidar or rowecef or roxcef or roxon or samixon or sintrex or socef or sunflow or tacex or torocef-1 or trexofin or triaken or triax or triaxone or tricefin or tricephin or trijec or xtenda or zefaxone or zefone 250).tw,kf. | 24945 |
| 94 | (Fluoroquinolone* or Ciprofloxacin* or Fleroxacin* or Enoxacin* or Enrofloxacin* or Gatifloxacin* or Gemifloxacin* or Moxifloxacin* or Norfloxacin* or Ofloxacin* or Levofloxacin* or Pefloxacin* or (Cephalosporin* or Cefalosporin* or Cefamandoleor* or Cefoperazone* or Cefazolin* or Cefdinir* or Cefepime* or Cefonicid* or Cefsulodin* or Ceftibuten* or Cefuroxime* or Cephacetrile* or Cefotaxime* or Cephalothin* or Cephapirin* or Cephalexin* or Cefaclor* or Cefadroxil* or Cefatrizine* or Cephaloglycin* or Cephradine* or Cephaloridine* or Ceftazidime* or Cephamycin* or Cefmetazole* or Cefotetan* or Cefoxitin*)).tw,kf. | 165066 |
| 95 | *immunomodulating agent/ | 9770 |
| 96 | *immunosuppressive agent/ | 20739 |
| 97 | ((immunomodula* adj2 (agent* or drug* or medication? or medicine? or compound? or therap*)) or immunomodulator? or immunotherap*).tw,kf. | 328210 |
| 98 | ((immunosuppress* or immuno-suppress* or immunodepress*) adj2 (agent* or drug* or medication? or medicine? or compound? or substance? or therap*)).tw,kf. | 88981 |

| | | |
|---|---|---|
| 99 | *rivaroxaban/ | 7425 |
| 100 | (rivaroxaban or assubex or ast-8294 or ast8294 or bay-59-7939 or bay-597939 or bay59-7939 or bay59-7939 or bay597939 or bs-112 or bs112 or dst-8294 or dst8294 or jnj-39039039 or jnj39039039 or kriva or naxat or rivaro or rivarolto or rivaxa or throsaben or xanirva or xarelto or xerdoxo or xindus).tw,kf. | 16680 |
| 101 | *cytokine receptor antagonist/ | 744 |
| 102 | (interleukin inhibitor? or cytokine antagonist or cytokine receptor block* or interleukin receptor antagonist?).tw,kf. | 310 |
| 103 | ((cilgavimab adj2 tixagevimab) or evusheld or "azd 1061 plus azd 8895" or "azd 1061/azd 8895" or azd-7442 or "azd 8895 plus azd 1061" or "azd 8895/azd 1061" or "azd1061 plus azd8895" or "azd1061/azd8895" or azd7442 or "azd8895 plus azd1061" or "azd8895/azd1061").tw,kf. | 506 |
| 104 | (itolizumab or alzumab or bmab-600 or bmab600 or eq-001 or eq001).tw,kf. | 144 |
| 105 | (dupilumab or bat-2406 or bat2406 or dupixent or regn-668 or regn668 or sar-231893 or sar231893).tw,kf. | 5031 |
| 106 | (pamrevlumab or fg-3019 or fg3019).tw,kf. | 265 |
| 107 | exp *monoclonal antibody/ | 291442 |
| 108 | ((monoclonal or clonal or hybridoma) adj3 antibod*).tw,kf. | 282883 |
| 109 | (bamlanivimab or etesevimab or "bamlanivimab/etesevimab" or "2423943-37-5" or "LY-3819253" or "LY-COV555" or "LY3819253" or "UNII-45I6OFJ8QH" or "WHO 11876" or "2423948-94-9" or "anti-Sars-cov-2 antibody JS016" or "CB6" or "JS016" or "LY COV016" or "LY-3832479" or "LY-COV016" or "LY3832479" or "NP005" or "UNII-N7Q9NLF11I" or "WHO 11873").tw,kf. | 720 |
| 110 | (casirivimab or imdevimab or "casirivimab/imdevimab" or regen-cov or "REGN-COV2" or regn10933 or regn10987 or "anti-sars-cov-2 regn-cov2" or "2415933-42-3" or "REGN-10933" or "REGN10933" or "UNII-J0FI6WE1QN" or "WHO 11861" or "2415933-40-1" or "REGN-10987" or "REGN10987" or "UNII-2Z3DQD2JHM" or "WHO 11863").tw,kf. | 696 |
| 111 | *bamlanivimab plus etesevimab/ or *bamlanivimab/ | 233 |
| 112 | *casirivimab plus imdevimab/ or *casirivimab/ | 316 |
| 113 | *sotrovimab/ | 234 |
| 114 | ("Sotrovimab" or "2423014-07-5" or "GSK-4182136" or "GSK4182136" or "UNII-1MTK0BPN8V" or "VIR-7831" or "VIR7831").tw,kf. | 527 |
| 115 | *amubarvimab/ | 11 |
| 116 | (amubarvimab or "BRII-196" or "BRII-198" or "DZIF-10c" or "BI 767551" or "SCTA01" or "Ty027" or "HLX70").tw,kf. | 49 |
| 117 | ("C144-LS" or "C-135-LS" or "C144-LS/C-135-LS").tw,kf. | 4 |
| 118 | ("COVI-GUARD" or "STI-1499").tw,kf. | 5 |
| 119 | ("COVI-AMG" or "sti-2020").tw,kf. | 3 |
| 120 | *enuzovimab/ | 1 |
| 121 | *plutavimab/ | 0 |
| 122 | (enuzovimab or plutavimab).tw,kf. | 0 |
| 123 | *cilgavimab plus tixagevimab/ | 244 |

| | | |
|---|---|---|
| 124 | lomtegovimab/ | 7 |
| 125 | *baricitinib/ | 1473 |
| 126 | HFB30132A.tw,kf. | 1 |
| 127 | ("ABBV-47D11" or "ABBV-2B04" or "AZD7442").tw,kf. | 52 |
| 128 | ("BI 767551" or "DZIF-10c" or "COR-101").tw,kf. | 8 |
| 129 | (baricitinib or olumiant or incb28050 or ly3009104).tw,kf. | 2834 |
| 130 | ((immune adj3 (sera or serum? or plasma)) or antisera or antiserum or immun#serum).tw,kf. | 77757 |
| 131 | exp *antiserum/ | 9535 |
| 132 | exp *immunoglobulin/ | 164377 |
| 133 | (immune* globulin? or immuneglobulin? or immunoglobulin?).tw,kf. | 235781 |
| 134 | exp *convalescent blood product/ | 1265 |
| 135 | ((convalescent adj2 (plasma or serum or sera)) or (plasma adj3 therap*)).tw,kf. | 20928 |
| 136 | *ibuprofen/ | 14745 |
| 137 | (ibuprofen or abfen or "aches-n-pain" or "act-3" or actiprofen or "adex 200" or adex liqui-gels or advil or afebril or aktren or aktren spezial or algiasdin or algifor or algofen or algoflex or allipen or alvofen express or "am-fam 400" or anadin or anadvil or analgyl or anbifen or anco or andran or anflagen or antalgil or antarene or antiflam or apo-ibuprofen or aragel or "atril 300" or attritin or balkaprofen or berlistar or bestafen or betaprofen or bifen or bluton or brufanic or brufedol or brufen or brufort or brugesic or brumare or brumed or brupro or buburone or bufect or bufohexal or bupogesic or burana or butacortelone or butifen or caldolor or calprofen or cap-profen or cenbufen or codral period pain or combiflam or contraneural or cuprofen or dalsy or "dc 7034" or "dc7034" or "dg 7034" or "dg7034" or dibufen or "diffutab sr 600" or dimidon or dolan fp or dolgit or dolobene ibu or dolocyl or dolodolgit or dolofen-f or dolomax or dolormin or dolval or donjust b or dorival or druisel or easifon or ecoprofen or emflam or epobron or ergix douleur et fievre or eudorlin extra or exidol or expanfen or febratic or febryn or femapirin or fenalgic or fenbid or flamicon or flarin or froben dolore or galprofen or gelufene or gyno-neuralgin or halprin or haltran or hemagene tailleur or h-loniten or ib-100 or ibalgin or ibofen or ibosure or ibu or ibuberl or ibucalm or ibudak or ibudol or ibudolor or ibufarmalid or ibufen or ibuflam or ibufug or ibugel or ibugesic or ibukern or ibuleve or ibulgan or ibuloid or ibumetin or ibumousse or ibunin or ibupen or ibupirac or ibuprin or ibuprocin or ibuprofene or ibuprohm or ibuprom or iburon or ibusal or ibuspray or ibustar or ibusynth or ibutop or ibux or ibuxin or idyl sr or ifenin or infant's motrin or infibu or inflanor or inflanor forte or ipren or irfen or junifen or junipro or kenfen or kontraneural or lamidon or librofem or lidifen or liptan or lopane or malafene or maxagesic or "mcn r 1451" or medicol or medipren or mediprin or mensoton or midol or momentact or motrin or mynosedin or nagifen-d or napacetin or neobrufen or neobrufen retard or nerofen or neutropain or nobfelon or nobgen or norflam-t or noritis or norton or novogent or novoprofen or nugin or nuprin or nureflex or nurofen or optifen or opturem or ostarin or ostofen or ozonol or paduden or panafen or pedea or pediacare fever or pediprofen or perdophen pediatrie or perofen or phorpain or phorpain gel or proartinal or profen or profeno or proff or proflex or proris or provin or provon or quadrax or rafen or ranofen or rapidophen or rapidophen forte or ratiodolor or rebugen or renidon or reuvol or rhelafen or roidenin or rufen or rupan or saridon n or schufen or seclodin or solufen lidose or solvium or spalt or syntofene or tabalon or tab-profen or taskine or tatanal or tofen or trendar or umafen or unipro or upfen or uprofen or urem or viamal febbre e dolore or zafen or zofen).tw,kf. | 28379 |
| 138 | exp *serotonin uptake inhibitor/ | 123212 |
| 139 | (((serotonin or 5-Hydroxytryptamine or 5-HT) adj3 (reuptake-inhibitor? or uptake-inhibitor?)) or SSRI? or fluvoxamine or du-23000 or du23000 or fluoxamin? or fluroxamine?).tw,kf. | 38100 |

| | | |
|---|---|---|
| 140 | exp *serotonin noradrenalin reuptake inhibitor/ | 81816 |
| 141 | (SNRI* or SSNRI* or ((((noradrenalin or norepinephrine) adj2 serotonin) or dual) adj3 (reuptake-inhibitor* or uptake-inhibitor*))).tw,kf. | 5512 |
| 142 | *citalopram/ or *escitalopram/ or *fluoxetine/ or *sertraline/ or *paroxetine/ or *desvenlafaxine/ or *duloxetine/ or *milnacipran/ or *milnacipran/ or *venlafaxine/ | 30876 |
| 143 | (celexa or citalopram* or cytalopram or seropram or escitalopram* or lexapro or fluoxetin* or prozac or sarafem or altruline or sertraline or aremis or besitran or gladem or lustral or sealdin or zoloft or aropax or paroxetin* or paxil or seroxat or (desvenlafaxine* or ellefore or khedezla or o-desmethylvenlafaxine or o-norvenlafaxine or pristiq* or cymbalta or duloxetin* or dalcipran or fetzima or impulsor or ixel or joncia or levomilnacipran* or midalcipran or milnaneurax or savella* or toledomin* or dobupal or efexor or effexor or grz5rcb1qg or venlafaxin* or trevilor or vandral)).tw,kf. | 55618 |
| 144 | *metformin/ | 25732 |
| 145 | (metformin or anj-900 or anj900 or apophage or aron or benofomin or dabex or denkaform or deson or dextin or diabetase or diabetformin or diabetmin or diabetosan or diabex or diafat or diaformin or diaformina or diametin or diamin or dianben or diformin or diformin retard or dimefor or dimethylbiguanide or dimethyldiguanide or dmgg or dybis or efb-0027 or efb0027 or eraphage or espa-formin or euform retard or fluamine or flumamine or fornidd or fortamet or glafornil or glibudon or glifage or gliguanid or glucaminol or glucofage or glucofago or glucoform or glucoformin or glucohexal or glucoless or glucomet or glucomin or glucomine or gluconil or Glucophage* or glucostop or glucotika or gludepatic or glufor or gluformin or glukophage or glumeformin or glumet or glumetza or glupa or glustress or glyciphage or glycomet or glycon or glycoran or glyformin or glymet or haurymellin or hipoglucin or islotin or jesacrin or juformin or la-6023 or la6023 or maformin or meglucon or meguan or melbin or melformin or mellittin or merckformin or mescorit or metaformin or metfogamma or metfoliquid geriasan or metforal or metformax or metformin hydrochloride or metformina or metformine or methformin or metiguanide or metomin or metphormin or miformin or neoform or newmet or nndg or reglus-500 or riomet or risidon or rudimet or siamformet or siofor or thiabet or vimetrol or walaphage).tw,kf. | 51563 |
| 146 | *ivermectin/ | 5843 |
| 147 | (ivermectin or bibovel or cardomec or cevamec or diapec or driponin or efacti or epimek or equimec or eqvalan or eqvalen or iveraxiro or ivercare or ivergalen or ivergelan or ivermectina or ivermectol or iverscab or ivertin or ivexterm or ivomec or k-237 or k237 or loutol or manburesa or mectizan or mk-933 or mk933 or oramec or posela or quanox gotas or revectina or romitu or scaball or scabioral or scatol or securo or sklice or soolantra or stromectol or vona or zulima).tw,kf. | 10651 |
| 148 | *resveratrol/ | 12069 |
| 149 | (resveratrol or srt-501 or srt501).tw,kf. | 22025 |
| 150 | *naltrexone/ | 6658 |
| 151 | (naltrexone* or abernil or addex-1000 or addex1000 or adepend or antaxon or antaxone or celupan or cyto-201 or cyto201 or depade or dependex or en-1639a or en1639a or ethylex or irt-103 or irt-104 or irt103 or irt104 or lodonal or n-cyclopropylmethylnoroxymorphone or nalerona or nalorex or naltex or naltrel or naltrexin or nemexin or nodict or nutrexon or opnt-002 or opnt002 or phaltrexia or pti-901 or pti901 or regental or revez or revia or stat-200 or stat-201 or stat-205 or stat200 or stat201 or stat205 or tranalex or trexan or um-792 or um792 or vivitrex or vivitrol).tw,kf. | 11184 |
| 152 | *colchicine/ | 11853 |
| 153 | (colchicin* or colchicum-dispert or colchily or colchimedio or colchiquim or colchisol or colchysat or colcine or colcrys or colctab or colgout or colrefuz or gloperba or goutichine or goutnil or kolkicin or kolkisin or mitigare or mpc-004 or mpc004 or myinfla or tolchicine).tw,kf. | 22957 |

| | | |
|---|---|---|
| 154 | or/26-153 [Drugs] | 4995887 |
| 155 | exp diet therapy/ or ((diet* or nutrition*) adj2 (advice* or therap* or counsel* or change* or intervention* or program*)).tw,kf. or (dietitian* or nutritionist*).tw,kf. | 503853 |
| 156 | exp *stem cell transplantation/ or stem-cell transplant*.tw,kf. | 164451 |
| 157 | low level laser therapy/ or (photobiomodulation or photo-biomodulation or photo-bio-modulation or ((low-power or low-level or low-energy or low-intensity or soft or cold) adj2 (light or laser) adj2 (therap* or irradiat*)) or (laser adj2 biostimulat*) or lllt or ((laser adj2 phototherap*) or biostimulat*)).tw,kf. | 32822 |
| 158 | *rehabilitation/ or muscle training/ or exp *neurorehabilitation/ or daily life activity/ or exp *kinesiotherapy/ or rehabilitation.tw,kf. | 466815 |
| 159 | ((cognitive or neurologic*) adj4 (rehab* or motor-therap*)).tw,kf. | 9756 |
| 160 | exp *self care/ | 35204 |
| 161 | (((patient or self) adj (manag* or treat*)) or self care).tw,kf. | 145602 |
| 162 | exp *patient attitude/ or *health literacy/ or *patient education/ or (patient* adj3 (educat* or knowledg* or literacy or literate or behavior* or behaviour* or attitud* or belief* or believ*)).tw,kf. | 307723 |
| 163 | *self help/ | 6383 |
| 164 | (((self help or support*) adj3 (group? or club? or servic* or social*)) or (therap* adj3 (club? or group* or social*))).tw,kf. | 210902 |
| 165 | *disease management/ or ((manag* or cope or coping) adj3 (symptom* or disease* or condition* or pain or discomfort*)).tw,kf. | 245783 |
| 166 | exp *exercise/ or exp *sport/ or exp *physical activity/ or *fitness/ or exp *physical medicine/ | 730625 |
| 167 | ("physical activit*" or walk* or pedestrian* or bicycl* or cycling or cyclist* or biking or bike* or "active lifestyle*" or "aerobic fitness" or "aerobic exercise*" or (running not "running water") or runner* or jog* or swim* or yoga or (physical* adj2 (activit* or active or exercise*)) or ((exercise* or fitness or aerobic*) adj2 (regimen* or training or intervention* or program* or class* or course* or train* or rehab*))).tw,kf. | 781166 |
| 168 | (physiotherap* or ((physical or physio) adj1 (therap* or treat*))).tw,kf. | 109656 |
| 169 | *indigenous health care/ | 713 |
| 170 | ((indigenous or metis or aboriginal or inuit or (native adj (america* or alaska* or hawaii* or canad*)) or first nation* or first people*) and (healthcare or (health adj2 (care* or service* or program*)) or healing* or medic* or therap* or remed*)).tw,kf. | 28151 |
| 171 | exp *alternative medicine/ or *traditional healer/ or exp *traditional medicine/ or exp *medicinal plant/ or *herbaceous agent/ or exp *plant medicinal product/ | 1075207 |
| 172 | ((alternat* or complementary or folk or traditional or holistic or chinese or african or tribal) adj2 (medic* or therap* or healing* or treat* or remed*)).tw,kf. | 304448 |
| 173 | *hypnosis/ or (hypnotism or hypnosis or hypnotherapy).tw,kf. | 12792 |
| 174 | exp *acupuncture/ or *ayurvedic drug/ | 40680 |
| 175 | (ayurved* or kampo or kanpo or acupunctur* or homeopath*).tw,kf. | 61829 |

| | | |
|---|---|---|
| 176 | ((botanical* or herb* or plant or plants or plantlet* or root or roots or natural) adj2 (drug? or extract? or healing* or ingredient? or medic* or preparation* or product or products or remedies or remedy* or supplement* or treat* or therap*)).tw,kf. | 289666 |
| 177 | Rest/ or (rest or (avoid* adj2 (exercise* or exert*))).tw,kf. | 278148 |
| 178 | exp *mental health care/ or *social work/ or *psychosocial rehabilitation/ | 80958 |
| 179 | ((mental health or emotion* or psych* or wellbeing or well-being or stress* or wellness* or anxiet* or depress* or obsess* or ocd or mood* or posttrauma* or post-trauma* or ptsd or schizo* or personality disorder* or bipolar* or adhd or attention deficit* or addict*) adj3 (care or service? or support? or treat* or therap* or psychotherap* or counsel* or hotline*)).tw,kf. | 495624 |
| 180 | (lifestyle adj3 (change* or intervention* or advice*)).tw,kf. | 41042 |
| 181 | exp *aftercare/ or *outpatient/ or *outpatient care/ | 90571 |
| 182 | (aftercare or after care or follow-up care or referral?).tw,kf. | 263437 |
| 183 | *health care delivery/ or *nonbiological model/ | 70709 |
| 184 | ((healthcare or health care or clinical* or public health or community) adj4 (approach* or model? or service? or deliver* or system* or distribut* or framework? or guideline?)).tw,kf. | 696265 |
| 185 | (recovery adj2 (team* or clinic? or centre? or center? or model* or program*)).ti,ab,kf. | 6596 |
| 186 | ((interdisciplin* or inter-disciplin* or cross-disciplin* or multidiscipline* or multi-disciplin*) adj3 (team* or clinic? or centre? or center? or model* or program*)).ti,ab,kf. | 26991 |
| 187 | ((care or treatment or healthcare) adj2 (model* or path* or plan*)).ti,ab,kf. | 291765 |
| 188 | (care adj2 (coordinat* or co-ordinat*)).ti,ab,kf. | 19983 |
| 189 | (patient-centred care or patient-centered care or case manag*).ti,ab,kf. | 36423 |
| 190 | exp *patient referral/ | 23069 |
| 191 | (refer* adj4 (specialist? or clinic? or therapy or therapist or counsel*)).tw,kf. | 55507 |
| 192 | or/155-191 [other interventions] | 6096735 |
| 193 | (12 or 25) and (154 or 192) | 7071 |
| 194 | 3 or 193 | 7464 |
| 195 | Clinical study/ | 165677 |
| 196 | Case control study/ | 213259 |
| 197 | Family study/ | 25787 |
| 198 | Longitudinal study/ | 206445 |
| 199 | Retrospective study/ | 1567259 |
| 200 | Prospective study/ | 905333 |
| 201 | Randomized controlled trials/ | 269111 |

| | | |
|---|---|---|
| 202 | 200 not 201 | 894314 |
| 203 | Cohort analysis/ | 1115562 |
| 204 | (Cohort adj (study or studies)).mp. | 500125 |
| 205 | (Case control adj (study or studies)).tw. | 173110 |
| 206 | (follow up adj (study or studies)).tw. | 75278 |
| 207 | (observational adj (study or studies)).tw. | 268223 |
| 208 | (epidemiologic$ adj (study or studies)).tw. | 124520 |
| 209 | (prospect* adj (study or studies)).tw. | 329476 |
| 210 | Longitudinal.tw. | 457359 |
| 211 | Retrospective.tw. | 1311189 |
| 212 | 195 or 196 or 197 or 198 or 199 or 202 or 203 or 204 or 205 or 206 or 207 or 208 or 209 or 210 or 211 | 4396623 |
| 213 | Randomized Controlled Trial/ | 806896 |
| 214 | Randomized Controlled Trial (topic)/ | 269111 |
| 215 | Controlled Clinical Trial/ | 472383 |
| 216 | Controlled Clinical Trial (topic)/ | 13446 |
| 217 | Randomization/ | 99045 |
| 218 | Double Blind Procedure/ | 215796 |
| 219 | Single Blind Procedure/ | 53564 |
| 220 | placebo/ | 408881 |
| 221 | Control Group/ | 110751 |
| 222 | (random* or sham or placebo*).ti,ab,hw,kf. | 2626827 |
| 223 | ((singl* or doubl*) adj (blind* or dumm* or mask*)).ti,ab,hw,kf. | 370375 |
| 224 | ((tripl* or trebl*) adj (blind* or dumm* or mask*)).ti,ab,hw,kf. | 2294 |
| 225 | (control* adj3 (study or studies or trial* or group*)).ti,ab,kf. | 1777893 |
| 226 | (Nonrandom* or non random* or non-random* or quasi-random* or quasirandom*).ti,ab,hw,kf. | 72484 |
| 227 | allocated.ti,ab,hw. | 112944 |
| 228 | ((open label or open-label) adj5 (study or studies or trial*)).ti,ab,hw,kf. | 90016 |
| 229 | ((equivalence or superiority or non-inferiority or noninferiority) adj3 (study or studies or trial*)).ti,ab,hw,kf. | 19297 |
| 230 | (pragmatic study or pragmatic studies).ti,ab,hw,kf. | 989 |
| 231 | ((pragmatic or practical) adj3 trial*).ti,ab,hw,kf. | 9200 |

| | | |
|---|---|---|
| 232 | ((quasiexperimental or quasi-experimental) adj3 (study or studies or trial*)).ti,ab,hw,kf. | 20839 |
| 233 | (phase adj3 (III or "3") adj3 (study or studies or trial*)).ti,hw,kf. | 133292 |
| 234 | or/213-233 | 3879787 |
| 235 | 194 and (212 or 234) | 3098 |
| 236 | *case report/ or (case-stud* or case-report*).jw. or (case* and report*).ti. | 616526 |
| 237 | (editorial or conference abstract or conference paper or conference review).pt. | 6624884 |
| 238 | 235 not (236 or 237) | 2123 |
| 239 | limit 238 to yr="2022 - 2024" | 1777 |
| 240 | (202205* or 202206* or 202207* or 202208* or 202209* or 20221* or 2023* or 2024*).dc,dd,dp. | 3943805 |
| 241 | 239 and 240 | 1673 |
| 242 | PMIDS FROM MEDLINE | 761 |
| 243 | 241 not 242 | 1017 |

Table S9. Legend for database searches

| | |
|---|---|
| **/** | At the end of a term or phrase, searches for term or phrase as a subject heading (index term). |
| ***** | "unlimited" truncation symbol. It substitutes for any number of characters at the end of a term or the root of a term, to retrieve plurals and variant spellings<br><br>Before a subject heading, indicates that the subject heading is a main topic of the article |
| **?** | "wildcard" symbol. Substitutes for one or no characters |
| **.ab** | Abstract field code |
| **ab. /freq=2** | Searches for terms that are present at least twice in the abstract |
| **.kf** | Keywords assigned by authors |
| **.mp** | Multipurpose field code – searches in title, abstract, drug trade name (tn) and additional fields |
| **.ti** | Title field code |
| **.tw** | Text Word field code. In Embase includes title, abstract, and drug trade name (tn) |
| **adj#** | Operator that searches for terms adjacent within # number of words, in any order |
| **exp** | "Explodes" a subject heading by searching for the main subject heading and for all Narrower Terms |
